# The SENTINEL study of differentiated service delivery models for HIV treatment in Malawi, South Africa, and Zambia: research protocol for a prospective cohort study

**DOI:** 10.1101/2023.05.22.23290361

**Authors:** Sophie Pascoe, Amy Huber, Idah Mokhele, Nkgomeleng Lekodeba, Vinolia Ntjikelane, Linda Sande, Timothy Tcherini, Prudence Haimbe, Sydney Rosen, the AMBIT SENTINEL study team

**Author notes:** Author for correspondence: Sydney Rosen, Boston University School of Public Health, 801 Massachusetts Ave, Room 390, Boston, MA 02118, USA. Deceased. Funding: Bill & Melinda Gates Foundation.

## Abstract

**Background:** Many countries in sub-Saharan Africa are rapidly scaling up “differentiated service delivery” (DSD) models for HIV treatment to improve the quality of care, increase access, reduce costs, and support the continued expansion and sustainability of antiretroviral therapy (ART) programs. Although there is some published evidence about the health outcomes of patients in DSD models, little is known about their impacts on healthcare providers’ job satisfaction, patients’ quality of life, costs to providers or patients, or how DSD models affect resource allocation at the facility level.

**Methods:** SENTINEL is a multi-year observational study that will collect detailed data about DSD models for ART delivery and related services from 12 healthcare facilities in Malawi, 24 in South Africa, and 12 in Zambia. The first round of SENTINEL included a patient survey, provider survey, provider time-and-motion observations, and facility resource use inventory. A survey of clients testing for HIV and a supplement to the facility resource use component to describe service delivery integration will be added for the second round. The patient survey will ask up to 10 patients enrolled in each DSD model at each study site about their experiences in HIV care and in DSD models, costs incurred seeking treatment, and preferences for HIV service delivery. The provider survey will ask up to 10 providers per site about the impact of DSD models on their positions and clinics. The time-and-motion component will directly observe the time use of a sample of providers implementing DSD models. Finally, the resource utilization component will collect facility-level data about DSD model availability and enrollment and the human and other resources needed to implement them. SENTINEL is planned to include four or more approximately annual rounds of data collection between 2021 and 2026.

**Discussion:** As national DSD programs for HIV treatment mature, it is important to understand how individual healthcare facilities are interpreting and implementing national guidelines and how healthcare workers and clients are adapting to new models of service delivery. SENTINEL will help policy makers and program managers understand the benefits and costs of differentiated service delivery and improve resource allocation going forward.

## Introduction

To achieve global goals for the treatment of HIV, many countries in sub-Saharan Africa are experimenting with and scaling up differentiated service delivery (DSD) models. These models alter key characteristics of service delivery, such as location, provider cadre, or visit frequency. Common DSD models include multi-month dispensing of medications, facility-based “fast track” services, community- or home-based drug distribution, provider-led adherence clubs, and patient-led community adherence groups.

DSD models are assumed to generate a wide range of potential benefits. These include improved access and greater satisfaction with healthcare for clients, increased clinic efficiency and quality, lower costs to providers and patients, and better health outcomes(1). Evidence to support these assumptions is accumulating but remains scarce. A few DSD programs have been formally described and evaluated in the literature(2–11); many others are being implemented officially or as pilot projects under routine care, without formal evaluation. Models that have been rigorously evaluated have often been implemented in the course of randomized trials, rather than routine practice(12). While there is an increasing body of published evaluations of routine practice, for example in Malawi(13), Uganda(14), Zambia(15), and South Africa(16,17), these have been limited in scope and rarely compared to DSD models to conventional care.

One reason for the dearth of evidence about the impacts of DSD models is that most existing medical record and health facility reporting systems do not capture the data needed for DSD evaluation. Routinely generated electronic medical records, which were designed prior to the advent of DSD programs, typically do not document the details of individual patients’ participation in DSD models. While some countries have recently added such fields to their data capturing forms, completion of the fields remains inconsistent and inaccurate. Even in the instances where a specific model of care is reported or can be inferred from medical record data, the variables available do not allow evaluation of the costs and benefits of the model of care, separate from the general benefits of the HIV treatment program. Data about the impact of differentiated service delivery on healthcare providers’ responsibilities, use of time, and job satisfaction are not routinely collected at all. Without this information, the impact of DSD models on the health system, individual healthcare facilities, or patients themselves cannot be ascertained.

The multiyear SENTINEL study of differentiated service delivery aims to fill in some of these information gaps in three countries in sub-Saharan Africa—Malawi, South Africa, and Zambia—through multiple rounds of observational data collection. Here we describe the SENTINEL study protocol (Supplementary file 1), which specifies collection of survey data from patients and providers, observation of providers’ time use, and facility resource utilization at a selected set of healthcare facilities in the three focus countries.

## Methods/design

### Overview

SENTINEL is a multifaceted, multi-round, observational study intended to capture information about differentiated service delivery for HIV treatment and related topics, such as differentiated models of HIV testing and integration of care for other conditions into HIV treatment. SENTINEL is one activity of the AMBIT Project, an evaluation of the impact of differentiated service delivery supported by the Bill & Melinda Gates Foundation and implemented by the Health Economics and Epidemiology Research Office (HE^2^RO) in South Africa, the Clinton Health Access Initiative (CHAI) in Malawi and Zambia, and Boston University in the U.S.

The SENTINEL protocol (Supplementary File 1) includes a set of related objectives, each of which aims to explore a different aspect of differentiated HIV treatment models and relies on a different data set. The protocol specifies a number of research questions, including:

1. How has the introduction of DSD models for HIV treatment affected quality of care for patients, including those not enrolled in or eligible for DSD?
2. How has the introduction of DSD models affected individual providers’ perceived workloads and job satisfaction?
3. What is the allocation of clinicians’ and other staff time at the facility level, by staff cadre and type of patient, and how is it affected by the number and uptake of models at the site?
4. What do patients like and dislike about each model and what would their preferences be for different model characteristics in the future?
5. What are the direct and indirect time and financial costs to patients of participation in each model?
6. What are the costs to healthcare providers of DSD models?

The first round of SENTINEL consisted of four components: patient survey, provider survey, provider time-and-motion observations, and facility resource use. Two additional components, a survey of clients testing for HIV and a supplement to the facility resource use component to describe service delivery integration, were added for SENTINEL 2. Additional components may be added to later rounds, as new questions and relevant topics arise and the scope of differentiated service delivery evolves. Each of the components of SENTINEL enrolls a different study population and has a different target sample size, as described below. As SENTINEL is an entirely observational study that aims to describe service delivery, rather than comparing outcomes, sample sizes were chosen to optimize the use of study resources and time availability.

SENTINEL is planned to include at least four approximately annual rounds of data collection. The first round (SENTINEL 1.0) was conducted between June 2021 and February 2022. Data collection for the second round (SENTINEL 2.0) began in September 2022 and was completed in May 2023. Round 3 is expected to begin late in 2023 and Round 4 in late 2024. Additional rounds will continue annually thereafter as information needs and resources allow. Although there is a separate protocol for SENTINEL in each AMBIT focus country (South Africa, Malawi and Zambia) in order to meet each local ethics board’s formatting requirements, the survey instruments and study procedures are identical for all three countries. In this paper, we describe the SENTINEL 2.0 protocol, which contains several amendments to the original (SENTINEL 1.0) protocol and is the current version of the study design.

Below, we first describe the study sites. For each component of the study, we then describe the population and sample size, enrollment and data collection, and data analysis approaches. All data collection instruments are included as Supplementary Files.

### Study sites

The study’s name, SENTINEL, refers to the inclusion of a selected set of healthcare facilities in each study country. Each facility with its associated differentiated treatment models is referred to as a sentinel site. Based on available resources, we chose 12 sites each in Malawi and Zambia and 24 in South Africa for the first round of data collection. Initially, the local study teams identified provinces and districts that were accessible, had a high burden of HIV, utilized the national electronic medical record system, and jointly provided diversity in setting (rural, urban), patient volume, DSD model offerings, and nongovernmental support partners.

Site assessment visits and a review of facility-level and DSD model-specific aggregate indicators were also conducted to determine which models each site was offering. We simultaneously conducted a survey of nongovernmental support partners and other DSD stakeholders to learn more about the models that were in use countrywide(18). Within each district, we also engaged with the relevant Departments and Ministries of Health, including national and district government health officials with responsibility for the sites, to select a set of facilities that represented the desired diversity. The sites are described in Table 1.

**Table 1.** AMBIT SENTINEL study sites.

| Facility | Setting | Number active ART patients at time of site selection | DSD models for ART patients reported by site during assessment visit |
| --- | --- | --- | --- |
| **MALAWI** |  |  |  |
| **Blantyre District (Southern Province)** |  |  |  |
| M Facility 1 | Urban | 2,925 | 6MMD, teen club, welcome back model, high viral load clinic |
| M Facility 2 | Urban | 8,400 | 6MMD, teen club, intensified care ART clinic |
| M Facility 3 | Urban | 6,178 | 6MMD, teen club, MIP |
| M Facility 4* | Rural | 6,592 | 6MMD |
| **Chiradzulu District (Southern Province)** |  |  |  |
| M Facility 5 | Rural | 5,288 | 6MMD, teen club, MIP |
| M Facility 6 | Rural | 2,633 | 6MMD, teen club |
| M Facility 7* | Rural | 6,520 | 6MMD, teen club |
| M Facility 8 | Rural | 3,944 | 6MMD, teen club |
| **Lilongwe District (Central Province)** |  |  |  |
| M Facility 9 | Rural | 1,025 | 6MMD, CAG, community outreach |
| M Facility 10* | Rural | 2,728 | 6MMD, teen club, MIP, high viral load clinic |
| M Facility 11* | Urban | 24,247 | 6MMD, nurse outreach community ART, FT, teen club, late shift |
| M Facility 12 | Urban | 4,200 | 6MMD, community outreach |
| **SOUTH AFRICA** |  |  |  |
| **West Rand District (Gauteng Province)** |  |  |  |
| SA Facility 1 | Urban | 1,783 | Ex- pup, fac-pup, AC |
| SA Facility 2 | Rural | 1,803 | Ex-pup, fac-pup |
| SA Facility 3 | Urban | 1,897 | Ex- pup, fac-pup, AC |
| SA Facility 4 | Urban | 2,116 | Ex- pup, fac-pup, AC |
| SA Facility 5 | Rural | 2,301 | Ex-pup, fac-pup |
| SA Facility 6* | Urban | 2,959 | Ex-pup, fac-pup |
| **Ekurhuleni District (Gauteng Province)\*\*** |  |  |  |
| SA Facility 7 | Urban | 1,482 | Ex-pup, fac-pup |
| SA Facility 8 | Urban | 2,386 | Ex-pup |
| SA Facility 9 | Urban | 2,658 | Ex- pup, fac-pup, AC |
| SA Facility 10 | Urban | 3,502 | Ex-pup |
| SA Facility 11 | Urban | 4,560 | Ex-pup, fac-pup |
| SA Facility 12 | Urban | 7,213 | Ex-pup, fac-pup |
| **Ehlanzeni District (Mpumalanga Province)** |  |  |  |
| SA Facility 13 | Urban | 6,622 | Ex pup, fac-pup, Pele Box |
| SA Facility 14 | Rural | 3,553 | Ex-pup, fac-pup |
| SA Facility 15 | Rural | 1,943 | Ex-pup, fac-pup |
| SA Facility 16 | Rural | 3,001 | Ex-pup, fac-pup |
| SA Facility 17* | Urban | 5,234 | Ex-pup, fac-pup, Pele Box |
| SA Facility 18* | Urban | 5,515 | Ex-pup, fac-pup |
| **King Cetshwayo District (KZN Province)** |  |  |  |
| SA Facility 19 | Rural | 1,182 | Ex- pup, fac-pup, AC |
| SA Facility 20 | Rural | 1,509 | Ex- pup, fac-pup, AC |
| SA Facility 21 | Rural | 2,231 | Ex- pup, fac-pup, AC |
| SA Facility 22 | Rural | 3,361 | Ex-pup, AC |
| SA Facility 23 | Rural | 5,190 | Ex- pup, fac-pup, AC |
| SA Facility 24 | Urban | 7,934 | Ex- pup, fac-pup, AC, youth model |
| **ZAMBIA** |  |  |  |
| **Chisamba District (Central Province)** |  |  |  |
| Z Facility 1 | Rural | 771 | CAGs, FT, MMD |
| **Kampiri Mposhi District (Central Province)** |  |  |  |
| Z Facility 2 | Urban | 6,752 | CAGs, FT, 3-5MMD, 6MMD |
| **Kabwe District (Central Province)** |  |  |  |
| Z Facility 3 | Urban | 2,696 | CAGs, MMSD 3-5, 6MMD, scholar model |
| Z Facility 4 | Urban | 3,625 | CAGs, family-based CAGs, FT, 3MMD, 6MMD, scholar model |
| **Mumbwa District (Central Province)** |  |  |  |
| Z Facility 5* | Rural | 2,742 | CAGs, FT, MMD |
| Z Facility 6* | Rural | 4,332 | CAGs, 3-5MMD, 6MMD |
| **Chilanga District (Lusaka Province)** |  |  |  |
| Z Facility 7 | Urban | 1,586 | MMD, FT |
| **Chongwe District (Lusaka Province)** |  |  |  |
| Z Facility 8 | Rural | 3542 | Fast track, MMD, weekend model |
| **Kafue District (Lusaka Province)** |  |  |  |
| Z Facility 9 | Urban | 3,545 | Fast track, 3-5MMD, 6MMD, weekend model |
| **Lusaka District (Lusaka Province)** |  |  |  |
| Z Facility 10* | Urban | 9,572 | Fast track, MMD, CAGs |
| Z Facility 11* | Urban | 11,400 | Fast track, MMD |
| Z Facility 12 | Urban | 5,519 | Fast track, MMD, urban adherence group, scholar model |

MMD, multi-month dispensing; MIP, mother-infant pairs model; CAG, community adherence group; FT, fast track; ex-pup, external pickup point; fac-pup, facility pickup point; AC, adherence club
\*Denotes that facility was a hospital in Malawi and Zambia or Community Health Clinic in South Africa, as opposed to a primary healthcare clinic.
\*\*Ekurhuleni District in Gauteng Province was included only in SENTINEL Round 1 but excluded from Round 2 due to challenges with data access. It has been replaced by Alfred Nzo District in the Eastern Cape Province, with specific sentinel sites still being identified for inclusion in SENTINEL Round 3.

### DSD client survey

The DSD client survey focuses on ART patient experiences in their model of care, as detailed in Panel 1. Survey questions will address participants’ past HIV care, preferences for service delivery, costs of seeking HIV treatment, and satisfaction with the care they are receiving. Because none of the three focus countries’ electronic medical records capture details of DSD model enrollment, participation, or dis-enrollment, the Sentinel survey will also ask for detailed information about DSD participation history.

At each site and in each round of Sentinel, we will enroll a target of 10 clients per DSD model of care, in addition to 10 clients who are eligible for DSD but are not enrolled in a DSD model and are receiving conventional care and 10 clients who are not eligible for DSD and remain in conventional care. The maximum sample size is thus based on the number of models found at each site in the course of the survey, with actual enrollment also determined by whether a sufficient number of clients (10) were participating in each model and available for interviewing during the data collection period. The study protocol allows enrollment of a maximum of the number of sites per country (12 or 24) x number of models per site (assumed to be up to 5) x 10 participants per model, or 600, 1,200, and 600 participants in Malawi, South Africa, and Zambia per round, respectively.

To be eligible for client survey enrollment, participants must be adults (≥18), have been on ART at the study site for at least 6 months, have received at least one medication refill within their current models of care (including conventional care), and be visiting the clinic for routine HIV-related care. Those who are unable to communicate in any of the languages into which the questionnaire has been translated, not physically, mentally, or emotionally able to participate, or unwilling or unable to provide written informed consent are excluded.

Potentially eligible clients will be referred to the study team by clinic staff sequentially as they present at the facility for routine HIV-related care. After confirmation of survey eligibility (Supplementary file 2) and provision of written informed consent (Supplementary file 3), participants will be interviewed using a structured questionnaire (Supplementary file 2). The informed consent form will ask for consent to access participants’ electronic and paper clinic records, so that survey responses can be matched to clinical inputs (e.g. number of clinic visits) and outcomes (e.g. retention in care).

Data analysis will start with simple frequencies of responses to each question on the patient survey, by site, model, and patient type. Questions on patient satisfaction, barriers, preferences, etc. will be reported as frequencies, stratified by model of care, patient type, age group, and gender as data allow. We will then look for associations between patient and facility characteristics and models of care. We will also estimate retention in care and VL suppression to examine whether outcomes differ by model of care.

For client-level costs of seeking care, we will estimate total cost/healthcare system interaction and then multiply by the number of interactions per client year to estimate a cost/patient/year. Monetary and time costs for patients will be estimated separately; time costs will also be converted to a monetary value using the local minimum wage or another appropriate metric.

Electronic medical records (EMR) for each site will be collected after survey completion and linked to survey responses, so that the full study record for each participant includes both survey responses and medical record data. The EMR data will be drawn from TIER.Net in South Africa, the national HIV EMR in Malawi, and SmartCare in Zambia. EMR data will be used to ascertain survey participants’ health outcomes (retention in care, viral suppression), to allow analysis of associations between patient characteristics, models of care, and health outcomes. EMR data will also provide estimates of the average numbers of healthcare system interactions each patient has per year, so that annual costs of seeking treatment and provider time use per patient served can be calculated.

**PANEL 1.** Summary of patient survey questions and topics.

|  |  |
| --- | --- |
| *Demographic and socioeconomic characteristics* | Age, gender, nationality, marital/relationship status, housing, education, employment, household composition, economic status |
| *Healthcare access and costs* | History of healthcare use, time on ART, missed ART visits, comorbidity healthcare received at this facility, patient costs to access clinic, changes in healthcare access as a result of COVID |
| *HIV treatment experience (conventional care)* | Current and previous DSD model enrollment, time in model, numbers of on- and off-site interactions with healthcare system and time required per interaction; satisfaction with healthcare provided and clinic experience; concerns; knowledge of HIV and ART, and sources of information; and suggestions for improvement; experience, costs, satisfaction, and concerns with care |
| *DSD model experience* | Choice in enrollment, experience with models, numbers of interactions, time required for DSD model interactions and cost, information shared about models, reasons for switching models (if applicable), satisfaction and concerns with models, recommendations for model, suggestions for improvement, best and worst characteristics |

### Provider survey

The provider survey (Supplementary file 4) focuses on providers’ experience with and attitudes toward the introduction of DSD models, as summarized in Panel 2. The provider survey addresses the attitudes, experiences, and preferences of a wide range of healthcare providers who have direct or indirect involvement in differentiated service delivery. At each site, study staff will interview up to 10 healthcare providers who are involved in DSD for HIV treatment, including clinical and non-clinical staff. The maximum sample size in the study protocol per round of data collection is 120 for Malawi, 240 for South Africa, and 120 for Zambia.

To be eligible for the survey, providers must have been working in their current role at the study site for at least 6 months and provide written informed consent (Supplementary file 5). We will ask each facility manager to introduce us to the individuals who are most involved in DSD model implementation at the facility and, if there are more than 10 staff involved in implementation, to advise on whom to invite to participate. We will attempt to include staff from all relevant cadres employed at the site who support implementation (e.g. nurses, counselors, pharmacy staff, community health workers, clerks). For facilities that have fewer than 10 providers in total, we will enroll as many as are involved in DSD model implementation and meet other eligibility criteria. We will seek written informed consent for survey participation; no individual identifiers will be collected for the provider survey.

Analysis of provider survey data will start with simple frequencies of responses to each closed question, by provider cadre. We will then summarize responses to open-ended questions. If data allow, we will stratify results by the models of care in use at the site and/or by the proportion of patients enrolled in non-conventional (differentiated) models of care and look for associations between provider characteristics (e.g. number of years in position) and views on DSD models.

**Panel 2.**
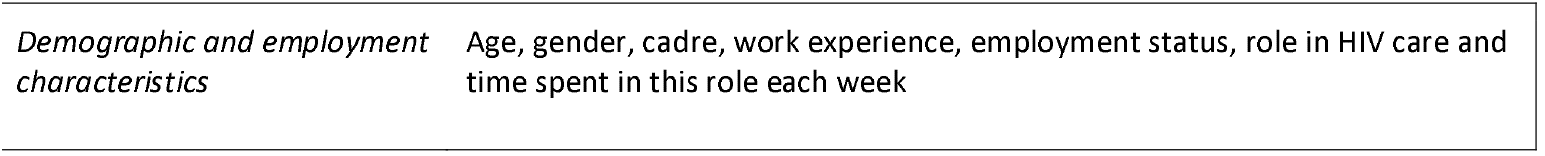
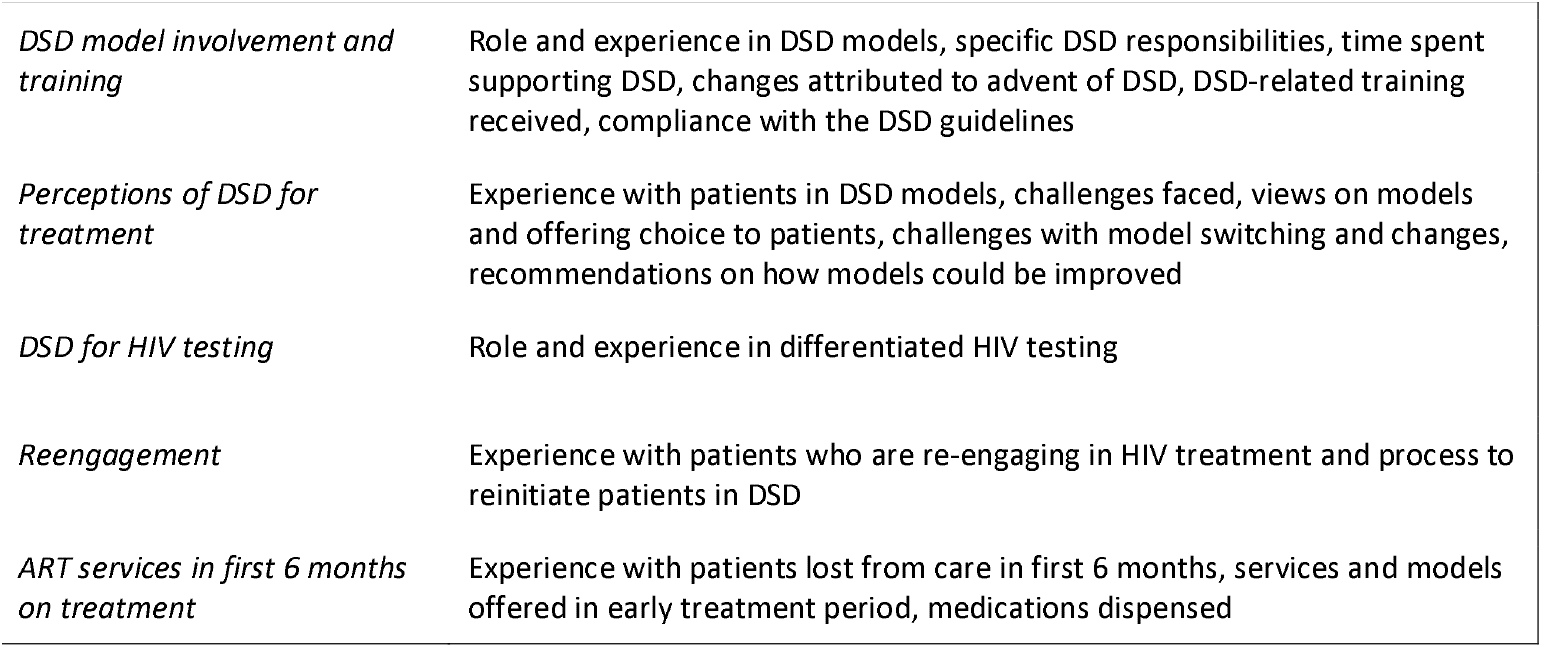
Summary of provider survey questions and topics.

### Time and motion observations

To assess how DSD models affect the use of providers’ time on a day-to-day basis, we will directly observe providers to record their time use. Each provider activity and time block, whether patient-facing or not, will be categorized and recorded by a study data collector. Providers will be asked for written informed consent for observation; no identifiers of any kind will be recorded, nor will data collectors observe actual patient interactions, only recording the start and end times of each consultation.

Up to 5 providers per site will be enrolled in the time and motion component, with each observed for up to two full working days. The total sample size will thus be 60 for Malawi, 120 for South Africa, and 60 for Zambia in each round of SENTINEL. Inclusion criteria for the time and motion component will be the same as for the provider survey, though individuals will not be required to participate in both. Potential participants will be identified by each facility’s operations manager and screened for eligibility by trained data collectors. After written informed consent is obtained, observation days will be scheduled. On each observation day, the SENTINEL data collector will arrive before the study participant and observe the participant’s time use from the start to the end of the working day. Each discrete activity and time block will be recorded by the data collector or the study participant; for patient interactions, study participants will complete a simple form indicating the primary purpose of the interaction and some patient characteristics (Supplementary File 6). These forms will be given to the data collector after each interaction and captured electronically.

We will analyze the time-and-motion data to generate mean time intervals, in minutes, for each provider cadre and time block use in the data set, including face-to-face interactions with patients, other time spent on patient care, and administrative time. Data will be pooled for all sites in each country to provide adequate sample sizes. Results will be used to estimate and compare staff time allocations per patient per year for each DSD model and explore whether DSD models are associated with differences in staff time use. Results will also be used to estimate the staff component of treatment costs, with provider fully loaded salaries (total cost to company) for each cadre multiplied by the time spent per patient.

### Resource utilization

This component of the study will not enroll human subjects but instead collect aggregate information about the study facilities. Data will be collected from each facility’s operations manager, ART staff, and/or other staff designated by the facility manager. Fields to be collected include a description of the HIV treatment program, DSD models for treatment and HIV testing offered, and DSD enrollment; staff numbers in each relevant cadre and proportions of staff time spent on DSD; ART patient volume; and details about each DSD model offered, including human and other resources needed to implement each model. We anticipate that not all sites will be able to provide data for all fields but that most can be completed. Resource utilization data will be collected in spreadsheets, as much of the data set is quantitative.

In SENTINEL 2.0, a section was added to the resource utilization instrument pertaining to how and to what extent HIV treatment is integrated with care for hypertension, diabetes, tuberculosis, any form of cancer, respiratory diseases, and/or mental health and with family planning services. This work aims to identify potential opportunities for simplifying schedules, so that HIV care visits overlap with scheduled clinic visits for comorbidities. Integration data will be reported as a taxonomy of approaches and descriptive statistics.

Resource utilization data will be used for descriptive reports on the status of DSD model (treatment and testing) implementation during each round of SENTINEL and to estimate the human and other resources required for each model, which will in turn serve as input to provider cost estimates. The data will also be used to estimate the number of patients on ART at each SENTINEL site and DSD uptake at the site, and to document the steps involved in linking patients to care who tested positive for HIV at the off-site facility linked testing locations.

### HIV testing models survey

Starting with Sentinel 2, the study will also enroll clients who are presenting at the sentinel clinics to have an HIV test and administer a survey (Supplementary file 7) to capture client experience with testing, understand the role of testing in engagement and re-engagement in care, and describe differentiated testing modalities. Survey questions will address the testing modality, reason for testing, location in the clinic tested, department referred from, other services provided, and HIV testing history. For those testing positive, questions will address prior ART exposure and readiness to initiate ART. For those who have previously been on ART, the questionnaire will ask about timing and reasons for disengagement, timing and reasons for re-engagement, and what services were provided at reengagement. Finally, for those testing HIV negative, questions will address the offer and uptake of PrEP and other preventive strategies after the negative test result.

Up to 50 adult (≥18) clients will be enrolled and consented (Supplementary file 8) per sentinel site, for a maximum sample size of 600 in Malawi, 1200 in South Africa, and 600 in Zambia. We will estimate and report simple frequencies of responses to each question on the patient survey, by site, model, and patient type. Questions on client satisfaction, barriers, preferences, etc. will be reported as frequencies, stratified by testing modality, client risk group, age group, and sex, as data allow.

### Limitations

We anticipate that the Sentinel study will have a number of limitations. First, the number of study sites is small, and generalizability to districts and provinces not included in the survey will require caution.

Second, in many cases survey participants are providing responses about their experiences with DSD models long after model enrollment, creating the possibility of recall bias. Third, as we can only enroll participants actively participating in ART care at health facilities, the study will not capture the experiences and perceptions of individuals who have already disengaged from care. Fourth, we will utilize routinely collected EMR data to ascertain treatment outcomes. While efficient in terms of resources, this approach is limited to data observed at the initiating facility and will not observe participants accessing care at other facilities. Silent transfers such as these may result in outcome misclassification among some classified as disengaged. Fifth, provider survey responses may reflect participants’ understanding of what is expected, rather than their actual practices or views. Although no identifiers are collected from providers, concerns about anonymity may influence provider responses. Sixth, the definition of a “model of service delivery” may be interpreted differently by different study sites and respondents, such that an approach to providing ART may be regarded by one site as a full DSD model and by another as services added to conventional care. The latter may not be reported to researchers as a DSD model in use at that site. Finally, as is often the case with research on HIV-related service delivery, secular changes to treatment delivery guidelines and/or other procedures are bound to occur over the course of SENTINEL, requiring adjustments to survey questions, eligibility criteria, and other study features as we go.

### Ethics

The SENTINEL study was approved with a separate protocol for each study country by the Boston University Institutional Review Board (Malawi H-41345, March 8, 2021; South Africa H-41402, March 11, 2021; Zambia H-41512, April 20, 2021) and by the University of the Witwatersrand Human Research Ethics Committee (Malawi M210270, June 9, 2021; South Africa M210241 June 4, 2021; Zambia M210342, September 30, 2021). In addition, the protocol for Malawi was approved by the Malawi National Health Science Research Committee (21/03/2672, March 12, 2021). The protocol for South Africa was approved by Provincial Health Research Committees through the National Health Research Database for each study district. The protocol for Zambia was approved by the ERES-Converge IRB (2021-Mar-012, April 7, 2021), the Zambia National Health Research Authority (NHRA00001/23/04/2021, April 23, 2021) and the Zambia Ministry of Health (MH/105/25/3, May 3, 2021).

### Study status

At the time of manuscript submission, SENTINEL has completed the first two rounds of data collection (labeled SENTINEL 1.0 and SENTINEL 2.0). Cleaning of SENTINEL 2.0 data is currently underway, with analysis expected during the remainder of 2023. The project has resources to implement two further rounds, with completion expected in 2026 or later, depending on whether additional rounds are conducted after the fourth.

## Discussion

The expansion of differentiated service delivery models for HIV treatment in sub-Saharan Africa has taken place swiftly, in response to international agency recommendations, donor agency pressure, encouragement from patient advocacy organizations, and generally positive findings from evaluations that have been conducted. Now that DSD programs are maturing, it is important to understand what is happening on the ground, as individual healthcare facilities interpret and implement national guidelines and healthcare workers and clients adapt to new models of service delivery. The SENTINEL study was designed to fill in gaps in routine data collection, with particular emphasis on the experiences of providers and clients. This information will help policy makers and program managers understand the benefits and costs of differentiated service delivery and improve resource allocation going forward.

We conclude by noting that implementation of SENTINEL is taking place concurrently with changes to national policies and procedures pertaining to the healthcare system, HIV testing and treatment, and DSD models in all three focus countries. Data about specific models of care, in particular, may become obsolete very soon after collection, as countries reconsider which models they wish to pursue for national implementation and/or alter guidelines in such a way that previous models of care are no longer relevant. We explicitly designed SENTINEL as a multi-year survey that would return to the same healthcare facilities each year but retain the flexibility to revise existing data collection instruments, introduce new areas of inquiry, and/or adjust enrollment criteria as suggested by secular changes. In this way, we hope that SENTINEL will allow us to keep up with the rapidly evolving healthcare systems we are studying and to continue to generate relevant evidence for improving those systems.

## Data Availability

The present work is a protocol paper and does not contain any data.

## List of abbreviations

AC: adherence club
ART: antiretroviral therapy
CAG: community adherence group
DSD: differentiated service delivery
Ex-pup: external pickup point
Fac-pup: facility pickup point
FT: fast track
MMD: multi-month dispensing
MIP: mother-infant pairs model

## Supplementary files

Supplementary file 1: Study protocol (example for South Africa, amended for SENTINEL 2.0)

Supplementary file 2: Patient survey screening form and survey instrument

Supplementary file 3: Patient survey consent information sheet and signature form

Supplementary file 4: Provider survey consent information sheet and signature form

Supplementary file 5: Provider survey instrument

Supplementary file 6: Time and motion data collection form

Supplementary file 7: Testing models survey instrument

Supplementary file 8: Testing models consent information sheet and signature form

## Declarations

### Ethics approval and consent to participate

Not applicable.

### Consent for publication

Not applicable.

### Availability of data and materials

Not applicable.

### Authors’ contributions

SP conceived of the study, drafted the initial instrument, drafted the initial manuscript, and reviewed and revised the final manuscript. AH conceived of the study, drafted the initial instrument, contributed to drafting the initial manuscript, and reviewed and revised the final manuscript. IM conceived of the study, drafted the initial instrument, contributed to drafting the initial manuscript, and reviewed and revised the final manuscript. NL contributed to drafting the initial instrument, contributed to drafting the initial manuscript, and reviewed and revised the final manuscript. VN contributed to drafting the initial instrument, contributed to drafting the initial manuscript, and reviewed and revised the final manuscript. LS contributed to drafting the initial instrument, contributed to drafting the initial manuscript, and reviewed and revised the final manuscript. TT conceived of the study, contributed to drafting the initial instrument, contributed to drafting the initial manuscript, and reviewed and revised the final manuscript. PH conceived of the study, contributed to drafting the initial instrument, contributed to drafting the initial manuscript, and reviewed and revised the final manuscript. SR conceived of the study, drafted the initial instrument, drafted the initial manuscript, and reviewed and revised the final manuscript. All individuals on the AMBIT SENTINEL study team contributed to conceiving of the study, drafting the initial instrument, and reviewing and revising the final manuscript. All authors read and approved the final manuscript.

## Acknowledgments

None.

