## Supplementary file 1 for "The SENTINEL study of differentiated service delivery models for HIV treatment in Malawi, South Africa, and Zambia: research protocol for a prospective cohort study"

**Research Protocol**

**Short title: SENTINEL 2.0 South Africa**

**August 28, 2022**
Version 2.1

BUMC IRB Protocol Number: H-41402 (United States)
Wits University HREC Protocol Number: M210241 (South Africa)

Federal-Wide Assurance Numbers:
FWA00000301 (Boston University Medical Campus)
FWA00000715 (University of the Witwatersrand)

**Contents**


1. SUMMARY OF SENTINEL-SOUTH AFRICA 4
2. INVESTIGATORS 4
3. BACKGROUND, RATIONALE AND OBJECTIVES 5
   a. *Background* 5
   b. *Rationale* 6
   c. *Objectives and research questions* 7
      (i) Domain 1: Provider time utilization 7
      (ii) Domain 2: Provider experiences 7
      (iii) Domain 3: Patient experiences with DSD models for ART 7
      (iv) Domain 4: Resource utilization (non-human subjects data) 8
      (v) Domain 5: Patient experiences with differentiated HIV testing 8
4. STUDY DESIGN AND PROCEDURES 8
   a. *Overview* 9
   b. *Study sites* 9
   c. *Domain 1 (Provider time)* 10
      (i) Sample selection 10
      (i) Inclusion and exclusion criteria 11
      (ii) Data collection 11
      (iii) Implementation of the study 12
      (iv) Data analysis 12
      (v) Informed consent 12
   d. *Domain 2 (Provider experiences)* 13
      (ii) Questionnaire 13
      (iii) Sample selection 13
      (iv) Inclusion and exclusion criteria 14
      (v) Implementation of survey 14
      (vi) Data analysis 14
      (vii) Informed consent 14
      (viii) Distress protocol 14
   e. *Domain 3 (Patient experiences with service delivery)* 15
      (i) Questionnaire 15
      (ii) Sample selection 15
      (iii) Inclusion and exclusion criteria 16
      (iv) Implementation of survey 17
      (v) Data analysis 17
      For each round of data collection, we will first estimate and report simple frequencies of responses to each question on the patient survey, by site, model, and patient type. 17
      (vi) Informed consent 17
      (vii) Distress protocol 18
   f. *Domain 4 (Resources)* 18
   g. *Domain 5 (Patient experience with differentiated HIV testing)* 18
      (i) Questionnaire 19
      (ii) Sample selection 19
      (iii) Inclusion and exclusion criteria 19
      (iv) Implementation of survey 20
      (v) Data analysis 20

(vi) Informed consent 20
(vii) Distress protocol 21
5. SAMPLE SIZE 21
6. DATA ENTRY AND STORAGE 22
7. DISSEMINATION OF FINDINGS 24
8. ETHICAL CONSIDERATIONS 24
*a. Potential risks and protections* 24
(i) Population 1: Providers to be observed for time-and-motion study 24
(ii) Population 2: Providers who participate in the provider survey 25
(iii) Populations 3 and 4: Patients who participate in the DSD of ART patient survey and patients who participate in the HIV testing survey 26
*b. Direct benefits* 27
*c. Indirect (societal) benefits* 27
*d. Informed consent* 28
*e. Subject confidentiality* 28
*f. Costs and payments* 28
*g. Access to data* 28
*h. COVID-19 considerations* 29
9. REFERENCES 29
10. APPENDICES 30

### 1. SUMMARY OF SENTINEL-SOUTH AFRICA

To achieve global goals for the treatment of HIV, many countries are piloting and scaling up differentiated service delivery models (DSD). A handful of efforts have been formally described and evaluated in the literature; many others are being implemented formally or informally under routine care, without a research or evaluation goal. For most countries however, we have little evidence on progress and challenges at the facility level—the number of patients actually participating in DSD models, health outcomes and non-health outcomes, effects on service delivery capacity and clinic efficiency and operations, and costs to providers and patients.

AMBIT is a set of data synthesis, data collection, and data analysis activities aimed at generating information for near- and long-term decision making and creating an approach and platform for ongoing evaluation of differentiated models of HIV treatment delivery. The first AMBIT protocol, "Gathering Records to Evaluate Antiretroviral Treatment" (GREAT, BU IRB H-38815, South Africa Wits HREC M190445), collects and analyzes comprehensive patient medical record data, allowing us to assess the effect of DSD models on patients' clinical outcomes and to evaluate uptake of DSD models at scale.

The **Sentinel-South Africa** study, the second AMBIT protocol, is examining the effect of DSD models on patient and provider satisfaction, service delivery capacity and quality, costs to patients, and other outcomes for which data are not routinely collected in patient-level medical records. The first round of Sentinel-SA was conducted in 2021. We are now amending the protocol to allow up to four additional annual rounds of data collection, in 2022-2025. We collected clinic aggregate data, conducted surveys of patients and providers, and observed operations at a selected set of 24 South African healthcare facilities and their affiliated DSD models in Round 1. Round 2 (2022) and later rounds will collect the same types of data and will expand the study's research questions to include differentiated models of HIV testing and linkage to care. Results are expected to inform South African policy makers and other local and international stakeholders on the actual implications of DSD models for patients, health system operations, and healthcare budgets.

### 2. INVESTIGATORS

This evaluation will be conducted by investigators from the Health Economics and Epidemiology Research Office (HE$^2$RO), part of Wits University in South Africa and Boston University in the U.S. Individual investigators are:


<u>Wits University (Wits Health Consortium), South Africa</u>

Sophie Pascoe (PI)
Principal Researcher
Health Economics and Epidemiology Research Office
Johannesburg, South Africa

Amy Huber (co-PI)
Senior Researcher
Health Economics and Epidemiology Research Office
Johannesburg, South Africa

Idah Mokhele

Senior Researcher
Health Economics and Epidemiology Research Office (HE$^2$RO)
Johannesburg, South Africa

Cheryl Hendrickson
Senior Researcher
Health Economics and Epidemiology Research Office
Johannesburg, South Africa

Linda Sande
Senior Researcher
Health Economics and Epidemiology Research Office
Johannesburg, South Africa

Boston University, United States

Sydney Rosen (PI)
Research Professor
Boston University School of Public Health
Boston, MA, USA

Nancy Scott
Assistant Professor
Boston University School of Public Health
Boston, MA, USA

### 3. BACKGROUND, RATIONALE AND OBJECTIVES

#### *a. Background*

To achieve global goals for the treatment of HIV, many countries are experimenting with and scaling up differentiated service delivery (DSD) models. These models alter key characteristics of service delivery, such as location, provider cadre, or visit frequency. Common DSD models include facility-based "fast track" services, community- or home-based drug distribution, multi-month dispensing of medications, and adherence clubs. In South Africa, where this study will take place, the Department of Health (DOH) and its implementing partners are currently supporting the scale-up of facility pick up points, external pick up points, and adherence club models of HIV treatment. A few other models are also being piloted at small numbers of sites in South Africa, at the initiative of NGO partners. In addition, DSD models for HIV testing are being explored to determine if the same advantages offered by differentiated treatment models—better retention in care, lower costs, reduced facility burden—can be extended to HIV testing.

In the published literature and in policy documents, DSD models are assumed to generate a wide range of potential benefits. These include increased clinic efficiency and capacity, lower costs to providers and patients, better health outcomes for HIV and non-HIV patients, and improved access and greater satisfaction with healthcare for patients. Despite a high level of confidence on the part of DSD advocates that at least some of these benefits will materialize, however, there is still relatively little evidence to support their assumptions. A few DSD programs have been formally described and evaluated in the literature[1–5]; many others are being implemented formally or informally under routine care, without a research or evaluation goal. Models that have been rigorously evaluated have often been implemented in the course of randomized trials, rather than routine practice[6]. Even evaluations of routine practice, as have been done in Malawi[7], Uganda[8], Zambia[9], and South Africa[10,11] have been limited in scope, have relied on assumptions due to missing data, or are already outdated. For most countries and models, we have little evidence of the impact of routinely-implemented DSD models on clinic efficiency and capacity; quality of care for HIV and non-HIV patients; or patient or provider satisfaction.

*b. Rationale*

The AMBIT project, supported by the Bill & Melinda Gates Foundation, is a set of data synthesis, data collection, and data analysis activities aimed at generating information for near- and long-term decision making and creating an approach and platform for ongoing evaluation of differentiated models of HIV treatment delivery. The first protocol developed for the AMBIT project, "Gathering Records to Evaluate Antiretroviral Treatment" (BU IRB H-38815, Wits HREC M190445), collects and analyzes patient medical record data, allowing us to assess the effect of DSD models on patients' clinical outcomes and to evaluate uptake of DSD models at scale. GREAT includes patient-level data collection from the national ART electronic medical record system (TIER.Net) and other paper and electronic sources, such as patient files and paper DSD model registers kept on site, CCMDD data from SyNCH, and data from the National Health Laboratory Service (NHLS).

While GREAT is answering some questions about DSD models in South Africa, one reason for the dearth of rigorous evaluations of DSD models in many African countries, including South Africa, is that medical records that we are relying on for the GREAT study do not capture the details of individual patients' participation in DSD models. Implementation of the models preceded efforts to adapt national paper and electronic record systems to include DSD-related data fields. While some countries have recently added such fields to their data capturing forms, as South Africa has done for adherence clubs and external pick up points, completion of the fields by clinic staff remains incomplete and inaccurate. Even in the instances where a specific model of care is reported or can be inferred from medical record data, the variables available tell us little beyond simple clinical outcomes, such as retention in care or viral suppression.

In addition to the limitations of existing medical records, patient- and facility-level data on the impact of DSD models on service delivery and non-clinical outcomes are not routinely collected in any country. Such data can therefore only be obtained by generation of new data through surveys and other methods and new analysis of aggregate data. Without this information, the impact of DSD models on the health system, individual healthcare facilities, or patients themselves cannot be ascertained. In combination with the GREAT-South Africa protocol, therefore, the Sentinel-South Africa study, the second AMBIT protocol, is examining the effect of DSD models of HIV treatment on patient and provider satisfaction, service delivery capacity and quality, costs to patients, and other outcomes for which data are not routinely collected in medical records. Starting with round 2 (Sentinel 2.0), data will be collected on the introduction of differentiated models of HIV testing.

In South Africa, we collected aggregate facility-level data, conducted surveys of patients and providers, and observed operations at a selected set of 24 sentinel healthcare facilities and their affiliated DSD models. The goal of the study, called Sentinel-South Africa, is to complement medical record data with primary information not currently available to policy makers through existing monitoring and evaluation procedures. Results are expected to inform South African policy makers and other local and international stakeholders on the actual implications of DSD models for patients, health system operations, and healthcare budgets.

*c. Objectives and research questions*

Sentinel-South Africa is a multi-faceted evaluation of the impact of differentiated service delivery for HIV testing and treatment on health facilities and patients in South Africa. Specific research questions for each model of care in use at each site, including standard or conventional care, and for the full cohort of ART patients, fall into five general domains. We note that Sentinel 1.0 included only the first four domains; Domain 5 has been added starting with Sentinel 2.0 to incorporate questions on HIV testing.

(i) Domain 1: Provider time utilization

DSD models are expected to reduce the average amount of time that clinicians spend per stable ART patient, which is in turn expected to increase the time they have available for non-stable ART patients and patients with other conditions. We ask the following questions:

1. What is the allocation of clinicians' and other staff time at the facility level, by staff cadre and type of patient, and how is it affected by the number and uptake of models at the site? This question will be answered using time-and-motion observations.

2. How has the introduction of DSD models affected quality of care for HIV patients and, if data allow, non-HIV patients, defined as provider (clinical and non-clinical) time spent per patient?

(ii) Domain 2: Provider experiences

DSD models are expected to improve healthcare providers' quality of professional life, by reducing the patient/provider ratio and allowing providers to spend more time with patients in need. It is likely that the scaleup of DSD models for testing and treatment has also changed facility procedures in other ways. We ask the following questions:

3. How has the introduction and scaleup of DSD models affected individual providers' perceived workloads?

4. How has the introduction and scaleup of DSD models affected individual providers' job satisfaction?

5. What have been the major facility-level procedural and other changes entailed in introducing, scaling up, and sustaining DSD models and how can implementation be improved in the future?

(iii) Domain 3: Patient experiences with DSD models for ART

A major goal of DSD models is to make ART delivery more "patient-centred," thus improving patients' experiences and reducing their costs of seeking ART and potentially increasing long-term adherence and retention in care. We ask the following general questions:

6. How much time do patients spend at facilities and at other DSD model venues for each model of care?

7. What are the direct and indirect costs to patients of participation in each model?

8. What do patients like and dislike about each model, how satisfied are they with their healthcare, and what would their preferences be for different model characteristics in the future?

(iv) Domain 4: Resource utilization (non-human subjects data)

A number of questions about the scale-up of DSD models for ART can be answered using aggregate, non-human subjects data that facilities typically collect for reporting purposes, such as number of patients and visits, staff availability, funding sources, etc. These data do not pertain to individual human subjects but are included here because they are integral to the overall goals of the study. We will ask the following questions:

9. Has the total number and/or mix of patients seen at the healthcare facility overall, including HIV and non-HIV patients, changed as the number of patients in DSD models has increased?

10. Has the total number or mix of providers at the healthcare facility overall, including for HIV and non-HIV services, changed as the number of patients in DSD models has increased?

11. Have standard operating procedures at the facility, such as how patients are scheduled for visits or the internal allocation of space or other resources, changed over the time period of observation?

12. What is the contribution of external (donor/NGO) resources to the operation of DSD models?

(v) Domain 5: Patient experiences with differentiated HIV testing

In the era of universal HIV treatment and same-day ART initiation, HIV testing is closely integrated with HIV treatment. Providers have begun to offer differentiated models of HIV testing to complement DSD treatment models. For the second and later rounds of this study, we will add a domain to capture patients' experiences with HIV testing and linkage to treatment. We will ask the following questions:

13. What differentiated models for HIV testing are in use?

14. What is the role of differentiated testing for those re-engaging in HIV care?

15. To what extent are those who test HIV-negative offered HIV prevention options?

Responses to all of these 15 questions will be estimated for individual models of care and compared among the models in use at each study site. Questions may be revised, refined, removed, or added based on data availability, Department of Health priorities, and other factors. The protocol will be amended whenever additional data, not described below, are needed.

### 4. STUDY DESIGN AND PROCEDURES

Note: In May 2022, the Sentinel-South Africa protocol is being amended to allow for up to five annual rounds of data collection from patients, providers, and facilities. Although a number of the questions in the data collection instruments have been revised, some aggregate data fields added or removed, and a second cohort of patients added to explore HIV testing, the procedures for and risks of the study are almost identical.

*a. Overview*

To answer the broad range of research questions listed above, the study will include surveys of patients and providers; a time and motion study of providers and observation of patient time use; and collection of non-human subjects indicators such as aggregate patient numbers, unit costs, and staff allocation records. With written informed consent of participants, data provided directly by patients (patient survey responses) will also be linked to their medical record data.

For purposes of this study, we will define each separate approach for interacting with ART patients that we observe at the study sites as a model of care. This will include conventional care (similar to the pre-differentiation model of care) for patients eligible for other models but not enrolled in them; conventional care for patients not eligible for other models, such as those with detectable viral loads; and each additional model of care offered by each study site. Common additional models of care currently in use in South Africa include facility pick up points, external pick up point and adherence clubs. Several less common models have also been implemented at the study sites, as shown in Table 1 below. In the second and later rounds of data collection, differentiated models of care for HIV testing will also be included.

*b. Study sites*

Round 1 of Sentinel-South Africa was conducted at 24 sites (healthcare facilities with their associated DSD models) in 4 districts of South Africa. Rounds 2 and later will be conducted at 18 of the same sites in 3 districts but one district will be exchanged for another in order to provide more geographic diversity to the study. All rounds will collect data from up to 24 sites. For Round 1, within each district, a preliminary set of 8 potential study sites was purposively selected based on availability of TIER.Net systems, ART patient volume, facility ownership (only public facilities were selected), and the variety of DSD models implemented. After brief site evaluations and in consultation with the DOH, the final 24 sites (6 per District) were selected, as shown in Table 1. For rounds 2 and later, we will exchange the sites in Ekurhuleni district for another district which will be determined in collaboration with the DOH. We note that these sites are intended to capture the variation among South African ART sites in terms of DSD model implementation, uptake, outcomes, costs, etc. They are not intended as a nationally representative group of facilities.

**Table 1. Study sites for Sentinel-South Africa**

| Facility | Setting | ART patients | Current DSD models (preliminary) |
|---|---|---|---|
| **West Rand District** | | | |
| Simunye Clinic (Westonaria) | Urban | 1,783 | Ex-PuP, AC, Fac-PuP |
| Tarlton Clinic | Rural | 1,803 | Ex-PuP, Fac-PuP |
| Fanyana Nhlapo Clinic | Urban | 1,897 | Ex-PuP, AC, Fac-PuP |
| Bekkersdal East Clinic | Urban | 2,116 | Ex-PuP, AC, Fac-PuP |
| Zuurbekom Clinic | Rural | 2,301 | Ex-PuP, Fac-PuP |

| Facility | Setting | ART patients | Current DSD models (preliminary) |
|---|---|---|---|
| Krugersdorp Central Clinic | Urban | 2,959 | Ex-PuP, Fac-PuP |
| **Ehlanzeni District** | | | |
| Msogwaba Clinic | Urban | 6,622 | Ex-PuP, Fac-PuP, Pele Box |
| Manzini Clinic | Rural | 3,553 | Ex-PuP, Fac-PuP |
| Legogote Clinic | Rural | 1,943 | Ex-PuP, Fac-PuP |
| White River Municipal Clinic | Rural | 3,001 | Ex-PuP, Fac-PuP |
| Nelspruit CHC | Urban | 5,234 | Ex-PuP, Fac-PuP, Pele Box |
| Kanyamazane Health Centre | Urban | 5,515 | Ex-PuP, Fac-PuP |
| **King Cetshwayo District** | | | |
| Mandlazini Clinic | Rural | 1,182 | Ex-PuP, AC, Fac-PuP |
| Ntuze Clinic | Rural | 1,509 | Ex-PuP, AC, Fac-PuP |
| Umkhontokayise Clinic | Rural | 2,231 | Ex-PuP, AC, Fac-PuP |
| Khandisa Clinic | Rural | 3,361 | Ex-PuP, AC |
| Phaphamani Clinic | Rural | 5,190 | Ex-PuP, AC, Fac-PuP |
| Richards Bay Clinic | Urban | 7,934 | Ex-PuP, AC, Fac-PuP, Youth model |
| **Ekurhuleni District (Round 1 only)*** | | | |
| Bonaero Park Clinic | Urban | 1,482 | Ex-Pup, Fac-PuP |
| Zonkizizwe 1 Clinic | Urban | 2,386 | Ex-PuP, Fac-PuP |
| Tamaho Clinic | Urban | 2,658 | Ex-PuP, AC, Fac-PuP |
| Kempton Park Civic Centre Clinic | Urban | 3,502 | Ex-PuP |
| Dresser Clinic | Urban | 4,560 | Ex-PuP, Fac-PuP |
| Goba Clinic | Urban | 7,213 | Ex-PuP, Fac-PuP |
| **Additional District – to be determined (Rounds 2 and later)*** | | | |
| 6 additional sites to be determined | TBD | TBD | TBD |

Ex-PuP=External pickup points; Fac-PuP=Facility pickup points; AC=Adherence clubs.

*For rounds 2 and later, six sites from a district other than Ekurhuleni will be chosen with input from the Department of Health for rounds 2 and later. We will obtain approval from NHRD for these sites as we have for the other districts.

*c. Domain 1 (Provider time)*

> *What is the allocation of clinicians' and other staff time at the facility level, by staff cadre and type of patient, and how is it affected by the number and uptake of models at the site?*
>
> *How has the introduction of DSD models affected quality of care for HIV and non-HIV patients, defined as provider (clinical and non-clinical) time spent per patient?*

Questions in Domain 1 pertain to the allocation and efficiency of provider time use within the study clinics, which are expected to change as result of the scaling up of DSD models. These questions will be answered with a time-and-motion study. Written informed consent will be sought from providers for the time-and-motion observations. The data collection instrument and consent information sheet and form are included as appendices to this protocol.

*(i) Sample selection*

A sample of up to five providers will be selected at each study site, for a total sample size of up to 120 for up to 24 sentinel sites in each round. The providers involved will include nurses, lay counsellors and/or community health workers, doctors/medical officers, and/or DSD model-specific staff, based on their role in providing HIV treatment. The participants will be purposively selected from within the site's staff cadre and invited to participate, in agreement with the individual serving as facility in-charge or facility manager.

Different calendar days will be observed during the study period, with days selected based on when each site schedules ART care and to represent typical patient care days for each cadre of provider. To minimize the burden on participants, no provider will be observed for more than two days.

*(i) Inclusion and exclusion criteria*

Inclusion criteria for the time and motion study are:

- Patient-facing or patient-supporting service provider at the study site (patient-supporting providers include data clerks, pharmacists, etc.)
- Directly or indirectly involved in the site's implementation of ART and DSD models
- Employed in current role at the study site for at least six months
- Provides written informed consent to participate

Exclusion criteria for the time and motion study are:

- None.

*(ii) Data collection*

We will collect the data fields listed below. We will also record non-patient-facing time, such as non-patient-facing duties (e.g. record-keeping, outreach activities, or administration) and other time (breaks, idle time, and transit).

For each participant, we will record the following demographic information:

- Gender
- Age
- Years of experience and years working at the site
- Provider cadre
- Current role
- Participation in earlier rounds of the Sentinel-South Africa time and motion study
- Participation in earlier or this round of the Sentinel-South Africa provider interview

For each interaction with a patient over the course of the day, we will record:

- Date
- Start time of daily observation (beginning of working day)
- Start time of activity block (e.g. patient consultation, break, meeting, etc.)
- End time of activity block
- Patient type (e.g. HIV, ART, other)
- Reason for visit/services provided at visit (e.g. consultation, counseling, rescripting)
- End time of daily observation (end of working day)

Depending on patient volume and provider role, the data set may include up to several dozen interactions over the course of a day, along with start and end times for non-patient facing duties and other time. As mentioned, no individual identifiers will be recorded.

*(iii) Implementation of the study*

In each round, research assistant will first request written informed consent from each individual provider identified as a potential participant. For participants who provide consent, the research assistant will be assigned to the participant for the entire working day. The research assistant, equipped with a tablet, will observe the start and end times of each interaction and the other data fields listed above. For this purpose, we expect to use SurveyCTO software loaded onto tablets (https://www.surveycto.com). Research assistants will be stationed in inconspicuous locations in the study sites, where they can make the required observations without causing disruption to clinic operations or inconveniencing patients. The research assistants will not be in the same space as the patients and <u>will not directly observe any patient-provider interactions</u>. Instead, the health care provider participating in the study will be asked to complete a short form for each patient to document the patient type that was seen during each activity block. No identifiers will be collected with these observations.

*(iv) Data analysis*

We will analyze the time-and-motion data to generate mean time intervals, in minutes, for each combination of provider cadre/patient type/interaction type in the data set (and to the extent that data allow) by round of data collection. For example, it is likely that at each site, one combination observed will be a nurse/stable ART patient/annual consultation and prescription refill. We will estimate the mean time required for this combination by pooling observations across all the Sentinel sites. We will repeat this exercise for all the common combinations observed, including for non-patient facing time. If data are not available for all three of the variables (provider cadre/patient type/interaction type) we will define types of interactions based on data we do have. Results will be used to estimate and compare staff time allocations per patient per year for each DSD model and determine whether DSD models are associated with an increase or decrease in the potential number of patients who can be managed with the existing staff complement. We will also look for associations between staff time use and the overall proportion of ART patients enrolled in DSD models and consider how this changes over time. Finally, results will be used to estimate the staff component of treatment costs, with provider fully loaded salaries (total cost to company) multiplied by the time spent per patient. We will also link the domain 1 data with the provider survey in order to compare provider satisfaction with actual time and motion observations.

*(v) Informed consent*

Written informed consent will be sought from each provider participating in the time-and-motion study in each round. Potential participants will be referred to the research assistant by the facility in-charge. The research assistant will explain that we are conducting a study to understand time use and would like to record the timing of the potential participant's interactions with ART and non-ART patients throughout the day. Potential participants will be assured that we will not observe actual interactions with patients, only record staff provider cadre (e.g. "nurse"), patient type (e.g. ART, non-ART HIV, non-HIV), interaction type (or reason for consultation) (e.g. "scheduled medical visit and medication prescription extension"), and start and end times for each interaction. Participants will be asked to complete a short form about the patient (no identifiers) for every patient interaction to avoid having study staff interact with patients.

Potential participants will also be informed that participation is completely voluntary and that observation can be halted at any time, if they do not feel comfortable. Participants will remain completely anonymous; no individual identifiers of any kind will be recorded for the participants or the patients they interact with, and facility names, while recorded, will not be reported in a way that allows respondents to be identified by their provider cadre. Participants who participated in the first round of data collection will also be allowed to participate in the second and later rounds of data collection and will be asked in each round if they have previously participated.

*d. Domain 2 (Provider experiences)*

Questions in Domain 2 pertain to providers' experiences with DSD models. These questions will be answered through a survey of providers at the study sites. Questionnaires will include both closed- and open-ended questions about the strengths and weaknesses of the models; how the advent of DSD models has changed provider responsibilities, work burden, and time allocation; and the effect of DSD models on job satisfaction. Written informed consent will be sought for provider interviews at each round of data collection.

> *How has the introduction of DSD models affected individual providers' perceived workloads?*
>
> *How has the introduction of DSD models affected individual providers' job satisfaction?*

*(ii) Questionnaire*

We will administer a questionnaire to a sample of up to 10 providers per study site per data collection round, with questions addressing providers' views on how the advent of DSD models has changed 1) individual job responsibilities and challenges; 2) facility operations in general; and 3) the respondent's job satisfaction. We will also ask participants to comment on challenges faced in implementing DSD models, the impact of DSD models on provider time allocation and efficiency (average time spent treating each patient) while caring for ART or non-ART patients, the implementation of differentiated HIV testing models, and other related topics. The questionnaire and consent information sheet and form are included as appendices to this protocol. No identifiers will be collected for any participant in the provider survey.

*(iii) Sample selection*

For each round of the survey, we will enroll up to 10 providers per facility, preferentially including all staff who manage ART patients and the site's operations manager, one or more lay counselors or community health workers, a pharmacist or pharmacy assistant, and any other cadre that is relevant to the DSD model program, as identified by the site. To select participants, we will ask the facility in-charge (facility manager) to choose the individuals in each cadre who are most involved in DSD model implementation. At some sites, each cadre will have only one representative on staff, and in some cases the cadre may be missing entirely. For facilities that have fewer than 10 providers in total, we will enrol as many as are involved in DSD model implementation and meet other enrollment criteria. At each site, we will work with the operations manager to select the most relevant individuals. Participants who participated in the first round of data collection will also be allowed to participate in the second and third rounds of data collection and will be asked in each round if they have previously participated. Responses will be linked across rounds for those who participate in >1 round.

*(iv) Inclusion and exclusion criteria*

Inclusion criteria for provider interviews are:

- Direct or indirect service provider at the study site (indirect providers include supervisors, technical advisors, etc.)
- Directly or indirectly involved in the site's implementation of ART and DSD models
- Employed in current role at the study site for at least six months
- Provides written informed consent to participate.

Exclusion criteria for provider interviews are:

- None.

*(v) Implementation of survey*

A Sentinel-South Africa study research assistant will administer the surveys in person in a private location at the clinic at a convenient time for the clinic staff member. We anticipate that each survey will take 45-60 minutes to complete.

*(vi) Data analysis*

We will first estimate and report simple frequencies of responses to each closed question on the provider survey, by provider cadre by data collection round. We will then summarize responses to open-ended questions. If data allow, we will stratify results by the models of care in use at the site and/or by the proportion of patients enrolled in non-conventional models of care. We will also link the Domain 1 data with the provider survey in order to compare provider satisfaction with actual time and motion observations.

*(vii) Informed consent*

Providers identified for potential survey participation will be asked for written informed consent at each round. The consent form will explain the purpose of the survey and assure participants that 1) they are not required to answer any questions they do not wish to and can stop the survey at any time; 2) they may decline to participate in the study entirely without any harmful consequences; and 3) no individual identifiers will be collected—we will not record names or any other identifying information. Those who provide consent will then be administered the survey.

*(viii) Distress protocol*

If a provider exhibits distress reflective of what would be expected in an interview about a sensitive topic, study staff will offer support and extend the opportunity to: (a) stop the interview; (b) regroup; (c) continue. If a participant's distress reflects acute emotional distress beyond what would be expected in an interview about a sensitive topic, study staff will take the following actions: (a) stop the interview; (b) give the provider a quiet private space and time interval to regroup if desired; (c) encourage the participant to contact his/her mental health provider, if they have one; (d) if the participant does not have a mental health provider, refer the participant to the clinic operations manager for more information on resources available to health care providers experiencing burnout or challenges related to workplace stress.

*e. Domain 3 (Patient experiences with service delivery)*

A quantitative, structured questionnaire will be administered to ART patients at each annual round of data collection to understand patients' satisfaction with their current model of care, motivation for enrolling in that model, and direct and indirect costs of accessing care. The focus will be on treatment service delivery, not on clinical aspects or outcomes of HIV treatment itself. Written informed consent will be sought for patient surveys.

*How much time do patients spend at facilities and at other DSD model venues for each model of care?*

*What are the direct and indirect costs to patients of participation in each model?*

*What do patients like and dislike about each model, how satisfied are they with their healthcare, and what would their preferences be for different model characteristics in the future?*

*(i) Questionnaire*

A structured questionnaire, designed for quantitative analysis, will be administered to a sample of patients at each site at each round of data collection.
Questions will address:

(i) Costs to patients of seeking care (transport, time, lost wages, child care, etc.)
(ii) Time required for seeking care (travel, time at healthcare facility, time participating in DSD interactions)
(iii) Patient satisfaction with their current model of care
(iv) Patient's preferences as to best and worst aspects of seeking care

Identifiers will be collected to allow questionnaire responses to be linked to respondents' clinical records. Written informed consent will be sought from all participants at each round of data collection, including consent to link questionnaire responses to clinical records and to collect data from clinical records. The questionnaire and consent information sheet and signature form are included as appendices to this protocol.

*(ii) Sample selection*

We will recruit adult ART patients who are enrolled in a defined model of care and present at the study sites or other DSD model venues or enrolled in home ART delivery model during the recruitment period. At the study sites, clinic staff will inform potentially eligible patients that they may be eligible to participate in a research study in a confidential space during routine visits. The information that the patient may be eligible to participate in a research study will not be provided in a common area or in the waiting area. In order to mitigate the potential sense of obligation to participants, patients who are interested in hearing more about the study will be referred to a research assistant for additional information.

For the home ART delivery model, starting in round 2 community health workers (CHW) will share the study information leaflet (flyer) and inform potentially eligible patients that they may be eligible to participate in a research study during routine home visits. In addition, there will be an invitation to the patient to participate in the study. The invitation will provide brief information regarding the study and provide the patient with research assistant contact details. If they are interested in participating or hearing more about the study, they could either send a call-back (please-call-me) to the phone number provided on the invitation. Alternatively, they can complete their contact details on the invitation and sign the form indicating that they give consent for the study team to contact them. The CHWs will return completed invitation forms to study staff at the clinic. Research assistants will then contact potentially eligible subjects who agreed to be contacted, to screen them. If patients are interested and eligible, the research assistant will set up an appointment for them to come to the clinic or meet at alternative secure locations to complete the consent process at a time convenient to the patient and where possible to coincide with their scheduled clinic visits. The study information leaflet and invitation to participate in the study are included as appendices to this protocol.

Prior to proceeding with screening and consent for all potentially eligible subjects, the research assistant will request verbal agreement to screening. The research assistant will explain that a study is underway and that patients who voluntarily enroll in the study will be asked questions about their feelings about the care at the clinic, costs incurred for care, and feelings about being enrolled in their DSD model. They will be told that the study will have no effect on the care they receive; that they do not have to participate in the study in order to continue to receive care at the site as they usually would; and that they do not have to answer any questions they do not wish to. The research assistant will enter the patient in the screening register and administer written informed consent; those who consent will then be administered the survey.

We will aim to enroll up to 10 patients per site per model of care. We will include conventional care defined as two models:  1) patients eligible for, but not enrolled, in a differentiated model other than conventional care and 2) patients not eligible for DSD models. Patients will be recruited consecutively as they complete their visits to the facility or other DSD model venue, based on availability of study interviewers. Study interviewers will await potential participants at locations convenient for each model of care (for example, for adherence clubs held at facilities, the interviewer will be available near the club's meeting place as soon as club activities have ended for a group of potential participants). We anticipate that each of the 24 sites for each round will include an average of 7 models of care, including the two conventional care models described above.

*(iii) Inclusion and exclusion criteria*

Inclusion criteria for the patient survey are:

- Living with HIV and on ART for at least six months at the study site
- ≥ 18 years old (18 and older considered adult for research purposes in South Africa)
- Enrolled in a specified model of care (including conventional care) up to the target number of participants for that model and have received at least one medication refill under this model
- Provide written informed consent to participate.

Exclusion criteria for the patient survey are:

- Unable to communicate in any of the languages into which the questionnaire has been translated or that is known to the research assistant

- Not physically, mentally, or emotionally able to participate in the study, in the opinion of the investigators or study staff.
- Unwilling to take the time required to complete the questionnaire on the day of consent.

Eligibility based on these criteria will be determined through completion of a survey screening form. The screening form will also allow us to compare the gender and age distribution of the population enrolled in the survey with those of the full potentially eligible population. The screening form is included as an appendix to this protocol.

*(iv) Implementation of survey*

For each round of data collection, a trained Sentinel-South Africa research assistant will administer the survey in person in a private location at the clinic or other DSD model venues either while the patient is waiting for services or after the patient has completed the visit. We anticipate that each interview will last 60 minutes, including the consent process. The questionnaire will document the subject's basic demographic and socioeconomic characteristics, details about their HIV care, their understanding of and satisfaction with aspects of the DSD or conventional care, and costs incurred while obtaining care, such as transport and lost wages.

*(v) Data analysis*

For each round of data collection, we will first estimate and report simple frequencies of responses to each question on the patient survey, by site, model, and patient type. For patient costs of seeking care, we will estimate total cost/healthcare system interaction and then multiply by the number of interactions per patient year to estimate a cost/patient/year. Monetary and time costs for patients will be estimated separately; time costs will also be converted to a monetary value using the local minimum wage or another appropriate metric. Questions on patient satisfaction, barriers, preferences, etc. will be reported as frequencies, stratified by model of care, patient type, age group, and gender as data allow.

*(vi) Informed consent*

In each round, upon referral to the research assistant, patients will receive a more complete description of the study, including the details of why it is being done, the types of questions that will be asked, and the need for written informed consent. Patients will be assured that participation is voluntary and that they can withdraw from the study at any time, without affecting the quality of care provided by the site. They will also be offered the opportunity to ask questions. If all inclusion/exclusion criteria have been met, the patient will be asked to provide written informed consent to participate.

After receiving verbal agreement from the potential participant, the research assistant will complete a screening form to record study eligibility for each patient screened. The screening form will not collect any identifiable information pertaining to individual patients prior to receipt of written informed consent. For patients who decline to participate in the study (consent refused), the study interviewer will indicate the refusal and, if offered, the reason for refusal in the screening form. Participants who participated in the first round of data collection will also be eligible to participate in the second and third rounds of data collection and will be asked in each round if they have previously participated.

The patient survey information sheet and consent form will be translated into the languages most commonly spoken by patients in the study districts, including Zulu and Sesotho, and possibly other languages as identified during site preparation visits. Translated consent documents will be submitted to required ethics committees for review prior to use with any study subjects.

*(vii) Distress protocol*

If a patient exhibits distress reflective of what would be expected in an interview about a sensitive topic, study staff will offer support and extend the opportunity to: (a) stop the interview; (b) regroup; (c) continue. If a patient's distress reflects acute emotional distress beyond what would be expected in an interview about a sensitive topic, study staff will offer support and take the following actions: (a) stop the survey; (b) give the participant a quiet private space to regroup; (c) encourage the participant to contact his mental health provider, if they have one; if the participant does not have a mental health provider, provide the participant with a list of resources available including appropriate hotlines and referral to the appropriate person at the local clinic.

*f. Domain 4 (Resources)*

In addition to the human subjects' data described above, we will collect facility-level, aggregate data on patient volumes, reasons for visits for HIV and non-HIV patients, staff complements, and other operational indicators during each round of data collection.

*Has the total number or mix of patients seen at the healthcare facility overall, including HIV and non-HIV patients and HIV patients in different stages of treatment, changed over the time period of observation?*

*Has the total number or mix of providers at the healthcare facility overall, including for HIV and non-HIV services, changed over the time period of observation?*

*Have standard operating procedures at the facility, such as how patients are scheduled for visits or the internal allocation of space or other resources, changed over the time period of observation?*

*What is the contribution of external (donor/NGO) resources to the operation of DSD models?*

The data to be collected for Domain 4 include routine reports and records generated by the facilities, District Health Offices, and nongovernmental partners. We will aim to collect the number of HIV, ART, and non-HIV patients presenting at the site in each time period and the number of full-time equivalent staff at the site in each time period, by cadre, including lay counselors, community health workers, and staff paid by external partners. We will also describe in detail the site's operating procedures with relevance to models of HIV treatment.

*g. Domain 5 (Patient experience with differentiated HIV testing)*

Beginning with Sentinel 2.0 (round 2), a quantitative questionnaire will also be administered to patients undergoing HIV testing to understand the presence of differentiated HIV testing and its role engagement or re-engagement in HIV care process and what services are being provided on the day of HIV testing for patients receiving positive and negative test results. Written informed consent will be sought for Domain 5 participants.

*(i) Questionnaire*

A structured questionnaire, designed for quantitative analysis, will be administered to a sample of patients at each site. Questions will address the testing modality, reason for testing, location in the clinic tested, department referred from, other services provided, and HIV testing history. For those testing HIV positive, questions will address prior ART exposure and readiness to initiate ART. For those who have previously been on ART, the questionnaire will ask about timing and reasons for disengagement, timing and reasons for re-engagement, and what services were provided at reengagement. For those testing HIV negative, questions will address the offer and uptake of PrEP and other preventive strategies after the negative test result.

Identifiers will be collected to allow questionnaire responses to be linked to respondents' clinical records. Written informed consent will be sought from all participants, including consent to link questionnaire responses to clinical records and to collect data from clinical records. The questionnaire and consent information sheet and form are included as appendices to this protocol.

*(ii) Sample selection*

We will recruit individuals who are participating in facility-run HIV testing during the recruitment period. Recruitment will occur either before the HIV test has been conducted, while the patient is in the queue, or after the test and subsequent procedures have been conducted.

At the study sites, clinic staff will inform potentially eligible patients that they may be eligible to participate in a research study in a confidential space during routine visits. The information that the patient may be eligible to participate in a research study will not be provided in a common area or in the waiting area. In order to mitigate the potential sense of obligation to participants, patients who agree will be referred to a research assistant, who will explain to each potentially eligible subject that a study is underway and that patients who voluntarily enroll in the study will be asked questions about their HIV testing history and ART history, and for re-engagers, the timing and reasons for disengagement and re-engagement. They will be told that the study will have no effect on the care they receive; that they do not have to participate to continue to receive care at the site as they usually would; and that they do not have to answer any questions they do not wish to. The research assistant will enter the patient in the screening register and administer written informed consent; those who consent will then be administered the survey.

We will aim to enroll up to 50 patients per site. Patients will be recruited consecutively either before they test for HIV or as they complete their testing visit to the facility or other HIV testing venue, based on availability of study interviewers. Study interviewers will await potential participants at locations convenient for each testing modality.

*(iii) Inclusion and exclusion criteria*

Inclusion criteria for the patient survey are:

- Undergoing HIV testing at the study site or other testing site within the catchment area
- ≥ 18 years old (18 and older considered adult for research purposes in South Africa)
- Provide written informed consent to participate.

Exclusion criteria for the patient survey are:

- Unable to communicate in any of the languages into which the questionnaire has been translated or that is known to the research assistant
- Not physically, mentally, or emotionally able to participate in the study, in the opinion of the investigators or study staff.
- Unwilling to take the time required to complete the questionnaire on the day of consent.

Eligibility based on these criteria will be determined through completion of a survey screening form. The screening form will also allow us to compare the gender and age distribution of the population enrolled in the survey with those of the full potentially eligible population. The screening form is included as an appendix to this protocol. Research assistants will request verbal agreement for screening using the script included in the screening form.

*(iv) Implementation of survey*

A trained Sentinel-South Africa research assistant will administer the survey in person in a private location at the clinic either while the patient is waiting for services or after the patient has completed the clinical visit. We anticipate that each interview will last 45 minutes, including the consent process. The questionnaire will document the subject's basic demographic and socioeconomic characteristics, information about HIV testing history and ART history, and for re-engagers, the timing and reasons for disengagement and re-engagement. Patients who initiate the study procedures before their HIV test will conduct informed consent and the first portion of the questionnaire, which collects information on demographics and HIV testing history. They will then be asked to return to the interviewer after their HIV test and subsequent procedures are completed to complete the questionnaire domains that include readiness to initiate, ART history, and questions about re-engagement. The patient will also be given the option of a telephonic follow up one week after the informed consent date if they would prefer that to completing the questionnaire on the same day. For those who initiate the informed consent after their HIV test and subsequent procedures, all components will be done at once.

*(v) Data analysis*

We will first estimate and report simple frequencies of responses to each question on the patient survey, by site, model, and patient type. Questions on patient satisfaction, barriers, preferences, etc. will be reported as frequencies, stratified by model of care, patient type, age group, and gender as data allow.

*(vi) Informed consent*

Upon referral to the research assistant, patients will be read a statement asking if they agree to be screened for the study. Those who agree will receive a more complete description of the study, including the details of why it is being done, the types of questions that will be asked, and the need for written informed consent. Patients will be assured that participation is voluntary and that they can withdraw from the study at any time, without affecting the quality of care provided by the site. They will also be offered the opportunity to ask questions. If all inclusion/exclusion criteria have been met, the patient will be asked to provide written informed consent to participate. The research assistant will complete a screening form to record study eligibility for each patient screened. The screening form will not collect any identifiable information pertaining to individual patients prior to receipt of written informed consent. For patients who decline to participate in the study (consent refused), the study interviewer will indicate the refusal and, if offered, reason for refusal in the screening form.

The patient survey information sheet and consent form will be translated into the languages most commonly spoken by patients in the study districts, including Zulu and Sesotho, and possibly other languages determined during site preparation visits. Translated consent documents will be submitted to required ethics committees for review prior to use with any study subjects.

*(vii) Distress protocol*

If a patient exhibits distress reflective of what would be expected in an interview about a sensitive topic, study staff will offer support and extend the opportunity to: (a) stop the interview; (b) regroup; (c) continue. If a patient's distress reflects acute emotional distress beyond what would be expected in an interview about a sensitive topic, study staff will offer support and take the following actions: (a) stop the survey; (b) give the participant a quiet private space to regroup; (c) encourage the participant to contact his mental health provider, if they have one; if the participant does not have a mental health provider, provide the participant with a list of resources available including appropriate hotlines and referral to the appropriate person at the local clinic.

### 5. SAMPLE SIZE

The total sample size for up to five rounds of the study is 12280. Table 2 presents the sample size for each round and data collection activity described above. All sample sizes are based on pooled analysis across all 24 study sites. We do not have preliminary data for any of the outcomes we intend to estimate, the study does not test a hypothesis, and all study procedures are minimal risk. We have therefore chosen to enroll a number of each type of participant that is feasible based on-site characteristics and study resources and will generate a large enough data set to analyze using standard methods.

**Table 2. Sample size**

| **Round 1 (Completed Sep 2021-Jan 2022)** | |
|---|---|
| **Study population** | **Sample size** |
| Domain 1: Time and motion study | Up to 120 providers (Up to 5 providers/site x 24 sites) |
| Domain 2: Providers surveyed | 240 providers (Up to 10 providers/site x 24 sites) |
| Domain 3: Patients surveyed | 1200 patients (Up to 10 patients/model x 5 models/site x 24 sites) |
| Total study subjects | =120 + 240 + 1200 +200 (potential for > 5 models)= 1760 study subjects maximum |

| Round 2 and later | |
|---|---|
| Study population | Sample size |
| Domain 1: Time and motion study | Up to 120 providers (Up to 5 providers/site x 24 sites) |

| | |
|---|---|
| Domain 2: Providers surveyed | 240 providers (Up to 10 providers/site x 24 sites) |
| Domain 3: DSD patients surveyed | 1680 patients (Up to 10 patients/model x 7 models/site x 24 sites) |
| Domain 5: HIV testing patients surveyed | Up to 1200 patients (Up to 50 patients/site x 24 sites) |
| Total study subjects | =120 + 240 + 1680 + 1200 + 200 (potential for > 7 models)= 3440 study subjects maximum |

| | |
|---|---|
| Total study sample size for Sentinel-South Africa Rounds 1 through 5 | 15520 study subjects maximum (1760 for Round 1 + 3440 for Round 2 + 3440 for Round 3 + 3440 for Round 4 + 3440 for Round 5) |

The target total sample size for the Round 1 of the study is 1560 subjects. To account for withdrawal after consent and/or the possibility of a site having more than 5 models, we will increase this to a maximum of 1760 study subjects for all of Round 1 of Sentinel-South Africa.

The target total sample size for the Round 2 and later is 3240 subjects per round. To account for withdrawal after consent and/or the possibility of a site having more than 7 models, we will increase this to a maximum of 3440 study subjects per round for Round 2 and later of Sentinel-South Africa.

Combining the total number of participants from Round 1, Round 2, Round 3, Round 4 and Round 5 of Sentinel-South Africa, the total study sample size for this protocol is 15520.

### 6. DATA ENTRY AND STORAGE

Data entry and storage for each data set are described in Table 3.

**Table 3. Data entry and storage**

| Data set | Data entry and storage |
|---|---|
| Domain 1: Time and motion study | Data will be collected electronically using Survey CTO or a similar software program. In cases of power failures or difficulties with the tablets, data will be entered onto paper study forms and then transcribed into a database at a local study office. These forms will be stored in a locked cabinet at the study sites, with access limited to the study team. Electronic data files will be stored on secure, protected drives at the Health Economics and Epidemiology Research Office (HE$^2$RO) in Johannesburg and at Boston University in Boston, with access limited to relevant study staff.<br><br>All subjects will be assigned a seven-digit, sequential identification number. The study ID number will not be linked to any identifiers, which will not be collected for this domain of the study. Survey CTO requires secure log-in and access and the final dataset will be accessible only to the study team. |
| Domain 2: Provider survey | Provider survey responses will be entered into electronic databases at the time of interview, using tablets. If there are power failures, data will be |

| | |
|---|---|
| | entered onto paper study forms and then transcribed into a database at the local study office. Forms will be stored in a locked cabinet at the study sites, with access limited to the study team. Electronic data files will be stored on secure, protected drives at the Health Economics and Epidemiology Research Office (HE$^2$RO) in Johannesburg and at Boston University in Boston, with access limited to relevant study staff.<br><br>Provider survey records will not contain any individual identifiers. Survey CTO or a similar software program requiring secure log-in and access by invitation will be used to create an electronic database to manage quantitative study data. On a regular basis, the data will be converted to SAS, STATA or SPSS for final cleaning and data analysis. All analytic databases will be password protected with access restricted to the members of the study team. |
| Domain 3: Patient survey - DSD of ART | A screening register will be kept by the study interviewers to record the consent process and keep track of those who do not consent, to allow us to determine if our sample is biased by patient characteristics due to differential consent. The screening register will not contain any individual identifiers. It will request age category, gender, and information required to determine survey eligibility only. The register will be kept as a form on the screeners' tablets.<br><br>Patient survey responses will be entered into electronic databases at the time of interview, using tablets. If there are power failures, data will be entered onto paper study forms and then transcribed into a database at the local study office. Forms will be stored in a locked cabinet at the study sites, with access limited to the study team. Electronic data files will be stored on secure, protected drives at the Health Economics and Epidemiology Research Office (HE$^2$RO) in Johannesburg and at Boston University in Boston, with access limited to relevant study staff.<br><br>All subjects will be assigned a seven-digit, sequential identification number. The study ID number will be used to identify individual subjects in the study databases and for all data analysis. Survey CTO or a similar software program requiring secure log-in and access by invitation will be used to create an electronic database to manage quantitative study data. On a regular basis, the data will be converted to SAS, STATA or SPSS for final cleaning and data analysis. All analytic databases will be password protected with access restricted to the members of the study team. |
| Domain 4: Resources | Data for Domain 4 will consist of aggregate indicators which will be collected using a standard template at each site. This will not be human subjects-related data and confidentiality will not be required. |
| Domain 5: Patient survey - Differentiated HIV testing | A screening register will be kept by the study interviewers to record the consent process and keep track of those who do not consent, to allow us to determine if our sample is biased by patient characteristics due to differential consent. The screening register will not contain any individual identifiers. It will request age category, gender, and information required |

to determine survey eligibility only. The register will be kept as a form on the screeners' tablets.

Patient survey responses will be entered into electronic databases at the time of interview, using tablets. If there are power failures, data will be entered onto paper study forms and then transcribed into a database at the local study office. Forms will be stored in a locked cabinet at the study sites, with access limited to the study team. Electronic data files will be stored on secure, protected drives at the Health Economics and Epidemiology Research Office (HE2RO) in Johannesburg and at Boston University in Boston, with access limited to relevant study staff.

All subjects will be assigned a seven-digit, sequential identification number. The study ID number will be used to identify individual subjects in the study databases and for all data analysis. Survey CTO or a similar software program requiring secure log-in and access by invitation will be used to create an electronic database to manage quantitative study data. On a regular basis, the data will be converted to SAS, STATA or SPSS for final cleaning and data analysis. All analytic databases will be password protected with access restricted to the members of the study team.

### 7. DISSEMINATION OF FINDINGS

The primary audience for this evaluation is the South African Department of Health and its partners, which will use the results to improve, target, and budget for the national implementation of DSD models. Many of the findings, however, will likely be of broader interest in South Africa and other countries, where more information about DSD model costs and impacts are eagerly sought. Results of the evaluation will be made as widely available as possible, through journals, websites, and conferences. Only aggregated, stratified data will be presented; it will not be possible to identify any individual patients from any of the data that is presented.

### 8. ETHICAL CONSIDERATIONS

The evaluation will require ethical approval from the Institutional Review Board of Boston University and the University of the Witwatersrand's Human Ethics Research Committee.

#### *a. Potential risks and protections*

The study team will not collect any biomedical samples specifically for this study. Data for the study will be drawn from provider questionnaires, patient questionnaires, and direct observation. We therefore believe that our study poses no physical risks to subjects. We will request written informed consent for the four study populations with whom study staff will interact.

*(i) Population 1: Providers to be observed for time-and-motion study*

We have identified one potential risk for providers participating in the time-and-motion study.

Risk 1: Loss of confidentiality

Study staff will observe providers starting and ending each interaction with a patient in order to record the amount of time required for each type of interaction. Providers participating in the time-and-motion study will be asked for written informed consent. No identifiers of any kind will be collected about the provider. We will, however, collect the provider's staff cadre (e.g. enrolled nurse, pharmacy assistant, etc.), age and gender. At some study sites, only one individual will be in the cadre for which we collect data, making it potentially possible for someone with access to the data to identify that individual. If the person gaining access were a supervisor, knowledge of time spent on different types of interactions could be used to judge the individual's performance.

Protection against Risk 1:

Data from the time and motion study will be recorded on password protected tablets held by the study research assistants. The tablets will be kept in locked cabinets when not in use, with data transferred to the central database and removed from the tablets regularly. Data pertaining to individual staff members will not be provided to site managers or supervisors, who will receive only aggregate results. During analysis, data from all 24 study sites will be pooled, preventing any single individual from being identified.

*(ii) Population 2: Providers who participate in the provider survey*

We have identified two potential risks for providers participating in the provider survey.

Risk 1: Emotional distress

The provider survey will ask questions about job satisfaction and challenges that could cause emotional distress among participants.

Protection against Risk 1:

Study staff will be trained to identify distress among respondents. The distress protocol described above will be followed should any occur. We repeat it here for convenience.

> If a provider exhibits distress reflective of what would be expected in an interview about a sensitive topic, study staff will offer support and extend the opportunity to: (a) stop the interview; (b) regroup; (c) continue. If a participant's distress reflects acute emotional distress beyond what would be expected in an interview about a sensitive topic, study staff will offer support and take the following actions: (a) stop the interview; (b) give the provider a quiet private space to regroup but observed by a member of the study team who has received counselling training; (c) encourage the participant to contact his/her mental health provider, if they have one; (d) if the participant does not have a mental health provider, refer the participant to the clinic operations manager for more information on resources available to health care providers experiencing burnout or challenges related to workplace stress.

Risk 2: Loss of confidentiality

Providers participating in the provider survey will be asked for written informed consent. No identifiers of any kind will be collected about the provider. We will, however, collect the provider's staff cadre (e.g. enrolled nurse, pharmacy assistant, etc.), age and gender. At some study sites, only one individual will be in the cadre for which we collect data, making it potentially possible for someone with access to the data to identify that individual. If the person gaining access were a supervisor, responses could be used to judge the individual's performance or affect his or her employment in other ways.

Protection against Risk 2:

Data from the provider survey will be recorded on password protected tablets held by the study research assistants. The tablets will be kept in locked cabinets when not in use, with data transferred to the central database and removed from the tablets regularly. Data pertaining to individual staff members will not be provided to site managers or supervisors, who will receive only aggregate results. During analysis, data from all 24 study sites will be pooled, preventing any single individual from being identified.

*(iii) Populations 3 and 4: Patients who participate in the DSD of ART patient survey and patients who participate in the HIV testing survey*

We have identified two potential risks for patients participating in the DSD and HIV testing patient surveys.

Risk 1: Emotional distress

The patient survey will ask questions about health and other topics that could cause emotional distress among participants, some of whom will have HIV and may have encountered obstacles in navigating the treatment process. Interacting with them in order to explain the study and confirm eligibility before requesting written informed consent may cause some emotional distress for some potential subjects.

Protection against Risk 1:

Study staff will be trained to identify distress among respondents. Site staff who introduce the study to potential subjects will be trained to assure potential subjects that referral to study staff and enrolling in the study are completely voluntary and that those who do not wish to enroll will receive exactly the same care as the study site would otherwise have provided. Potential subjects will also be told that they can discontinue participation even after consenting without any effect on their care. The distress protocol described above will be followed should any occur. We repeat it here for convenience.

> If a patient exhibits distress reflective of what would be expected in an interview about a sensitive topic, study staff will offer support and extend the opportunity to: (a) stop the interview; (b) regroup; (c) continue. If a patient's distress reflects acute emotional distress beyond what would be expected in an interview about a sensitive topic, study staff will offer support and take the following actions: (a) stop the survey; (b) give the participant a quiet private space to regroup; (c) encourage the participant to contact his mental health provider, if they have one; if the participant does not have a mental health provider, provide the participant with a list of resources available including appropriate hotlines and referral to the appropriate person at the local clinic.

Risk 2: Loss of confidentiality

Patients participating in the patient survey will be asked for written informed consent. We will collect data indicating individuals' HIV status, their opinions on the quality of the services received, and some sensitive health information. A breach of confidentiality, for example through inadvertent loss of a storage device or paper files, would thus pose a risk to subjects.

Protection against Risk 2:

To protect patients against this risk, patient identifiers will be collected and stored separately from all other individual data. Identifiers will be entered on site and stored in encrypted, password protected files, so that no paper records containing identifying information are removed from the sites. Names, national identification numbers, and other identifying information will be used only for the purposes of linking disparate sources of data for the same patient (e.g. electronic medical record information to paper clinical records). As soon as a specific source document has been linked to the patient of interest, data from it will be entered in a record containing the Study ID number only. Analytic data sets will not contain any identifiers, and the linking files containing the identifiers will be destroyed after linking is complete.

All study data, whether in electronic or paper format, will be stored in secure locations. Password-protected laptops or tablets used on site will be kept in locked and secure cabinets and rooms when not in use. Files will be transferred to the study office on a regular basis and stored on secure servers and in locked cabinets. Study staff will not be permitted to download de-identified data sets for cleaning or analysis except with the explicit permission of a co-investigator, and datasets will not be stored on individual hard drives when not in use. Upon completion of the study, computer files and any data collection forms containing study data will be retained for seven years and then destroyed.

All study staff will be trained in Good Clinical Practice, Research Ethics, and study procedures to ensure that they understand both research confidentiality requirements and study confidentiality procedures. Study investigators will monitor data collection on an ongoing basis. They will report to the BU IRB and the Wits HREC any breaches in confidentiality identified. In the event that a breach in confidentiality does occur, staff will be retrained on human subjects' protection and confidentiality if possible or removed from the study if either the breach is too serious or if the PIs feel the staff member cannot be sufficiently retrained. Staff will be made aware of this condition on employment.

*b. Direct benefits*

There are no direct benefits to study subjects enrolled in this study.

*c. Indirect (societal) benefits*

The indirect benefits of this study are expected to be large. The Government of South Africa is embarking on the scale up of some DSD models for testing and treatment that will ultimately affect hundreds of thousands of patients. Generating early evidence of the expected patient and provider impact and patient cost of these interventions has the potential to make DSD models more effective and less expensive nationwide. The evaluation will also assist the government to prioritize models that are more preferable for providers and patients, as well as those that are most efficient.

Because the indirect benefits of the study are large and the risks to human subjects are minimal, we are confident that the benefits justify the risks.

*d. Informed consent*

Written, informed consent will be sought from all participants in the time-and-motion study, provider survey, DSD patient survey, and HIV testing patient survey. The informed consent information sheet will describe the nature and goals of the study and assure subjects their information will be kept confidential. It will explain to subjects what will occur in the study and the procedures to be followed and/or questions to be asked. It will indicate that after the study has been completed, fully deidentified data may be posted to a public research repository, as is typically required for journal publication.

The consent form will be administered by a trained study research assistant. Participants will be assured that data collected for our study will be kept strictly confidential and will never be reported to clinic staff or anyone outside the study team. For the patient survey, the full informed consent information sheet and form will be translated into the languages most commonly used by patients at the study sites. For providers all interviews will be conducted in English.

Participants in all four components will be offered a copy of the information sheet to keep if they wish.

A brief screening questionnaire will be used to determine the eligibility of patients for participation in domain 3 and 5. Potential participants will be read a scripted statement by the research assistant to confirm their agreement to be screened. This statement is included at the start of each screening form.

*e. Subject confidentiality*

As explained above, we will take multiple steps to protect subject confidentiality. These are detailed in the paragraph entitled "Potential Risks and Protections."

*f. Costs and payments*

For Round 1 of Sentinel-South Africa, there will be no costs or payments to subjects for participating in this study. We intend to provide light refreshments to participants in the patient and provider surveys in appreciation of their cooperation.

For Round 2 and later, patients participating in the DSD patient survey and the patient tester survey will receive compensation valued at 150 ZAR in the form of a voucher to thank them for their time and willingness to participate. Participants in the time and motion and provider survey components will not receive compensation for their participation. We intend to provide light refreshments to participants in both the patient and provider surveys in appreciation of their cooperation.

*g. Access to data*

As is often required by journals for publication of research manuscripts, we will post completely anonymized data sets to a public repository after all analysis and publication has been completed. The informed consent forms will alert participants to this and indicate that in providing consent to participate in the study, they are also consenting to the posting of completely de-identified data to a research repository.

*h. COVID-19 considerations*

In light of COVID-19 and the need to adhere to safety protocols to protect study staff and participants, we will implement the following precautions:

- Fieldwork will only commence when permitted according to the country's lockdown regulations.
- Training plans for the study team will be evaluated at the time and if COVID risk is too high, training will be done remotely via Zoom.
- We will supply personal protective equipment (PPE) material including masks, hand sanitizers, and/or other applicable PPE to the study team
- Regular screening of study team members for COVID-19 symptoms will be conducted
- If any team members show COVID-19 symptoms, he/she will immediately report it to the study PI and action will be taken as specified in the COVI-19 SOP
- Study interviewers will capture responses directly in Survey CTO during the interview – therefore alleviating the need for printed questionnaires.
- Study interviewers will practice physical distancing (minimum of 1.5 meters apart) at all times while conducting fieldwork
- A separate COVID-19 SOP will provide detailed guidance on safety protocols that need to be followed while conducting fieldwork
- We will provide training on the COVID-19 SOP and procedures that need to be followed at the clinics/facilities, including infection prevention control measures and PPE use.

### 10. APPENDICES

*Data collection instruments*

1. Time and motion study data collection form (Attachment 1a)
2. Provider survey instrument (Attachment 2a)
3. DSD patient survey instrument (Attachment 3b)
4. DSD patient survey eligibility screening form (Attachment 3a)
5. DSD patient survey linking form (Attachment 3g)
6. Tester survey instrument (Attachment 5b)
7. Tester survey eligibility screening form (Attachment 5a)
8. Tester survey linking form (Attachment 5e)

*Consent forms and information sheets*

1. Time and motion study information sheet (Attachment 1b)
2. Time and motion study consent form (Attachment 1c)
3. Provider survey information sheet (Attachment 2b)
4. Provider survey consent form (Attachment 2c)
5. DSD patient survey information sheet (Attachment 3c)
6. DSD patient survey consent form (Attachment 3d)
7. Tester survey information sheet (Attachment 5c)
8. Tester survey consent form (Attachment 5d)

*Study flyers and invitations*

1. DSD patient survey study flyer (Attachment 3e)
2. DSD patient survey invitation (Attachment 3f)
