## Supplementary file 2 for "The SENTINEL study of differentiated service delivery models for HIV treatment in Malawi, South Africa, and Zambia: research protocol for a prospective cohort study"

### D3a. SENTINEL2.0-South Africa Patient Survey - DSD Outcomes

| Field | Question | Answer |
|---|---|---|
| Screening form | | |
| Screening form > Form control | | |
| screening_no *(required)* | Screening number<br>*screening_no* | |
| surveyor_id *(required)* | Surveyor ID | |
| specify_surveyor *(required)* | Specify the surveyor | |
| district *(required)* | District name | |
| wr_facilities *(required)* | Facility name | |
| mp_facilities *(required)* | Facility name | |
| kzn_facilities *(required)* | Facility name | |
| screening_date *(required)* | Date | |
| intro | I. Introduction<br>*Ask the patient for a few minutes of their time. Introduce yourself and the study. Provide information on the study as per the training and give the patient the information sheet for the study. If the patient expresses interest in being in the study, proceed to section II below and then complete the eligibility screening in section III. If the participant is not interested in being in the study, thank them for their time, end the interaction, and answer the questions in section II below.* | |
| Screening form > II. Demographic description | | |
| gender *(required)* | 1. Gender | 0 Male<br>1 Female<br>2 Other |
| age *(required)* | 2. How old were you at your last birthday?<br>*Age (years)* | |
| Screening form > III. Eligibility screening | | |
| Eligibility_note | If all answers to the following screening questions 3-5 and 7 are "YES", proceed to administer the informed consent process. If any answers are "NO", thank the patient for their time and end the interaction. | |
| HIV_care_at_current_facility *(required)* | 3. Are you currently enrolled in HIV care at this facility?<br>*STOP if the response is NO!* | 1 Yes<br>0 No |
| is_patient_18 *(required)* | 4. Are you currently age 18 or older?<br>*STOP if the response is NO!* | 1 Yes<br>0 No |
| onART_for_6months *(required)* | 5. Have you been taking ART for at least 6 months?<br>*STOP if the response is NO!* | 1 Yes<br>0 No |

| Field | Question | Answer |
|---|---|---|
| HIV_care_model *(required)* | 6. Which HIV care model are you currently enrolled in? (Multiple responses possible for integrated models)<br>*(Note: this question provides the basis for question 5.)* | 1 Standard or conventional care, not in an alternative model AND NOT eligible for an alternative model for stable patients<br>2 Standard or conventional care, not in an alternative model AND eligible for an alternative model but not currently enrolled<br>3 Adherence club<br>4 Facility Pick Up Point<br>5 External Pick Up Point<br>6 Youth club<br>7 Pele Box<br>9 Home ART delivery<br>10 Bicycle model<br>8 Other (specify) |
| other_HIV_care_model *(required)* | Specify the other HIV care model you are currently enrolled in | |
| collected_medication_once *(required)* | 7. Have you had at least one medication collection visit under the model of care you said you are enrolled in?<br>*STOP if the response is NO!* | 1 Yes<br>0 No |
| Screening form > IV. Eligibility decision | | |
| eligible | This patient is ELIGIBLE for the study. Please proceed with the informed consent process | |
| consenting *(required)* | This patient | 1 CONSENTED to participate (Fill in survey instrument ID number below and proceed with survey)<br>2 DID NOT CONSENT to participate (Thank the participant for their time and end the interaction)<br>3 N/A (Target for this model has been reached) |
| V. Patient survey | | |
| sid *(required)* | Survey ID options | 1 Barcode<br>2 Enter manually |
| barcode_scan *(required)* | Scan survey ID | |
| survey_id *(required)* | SURVEY ID | |
| V. Patient survey > Notes | | |
| facility_loc *(required)* | Location within facility | |
| intro_statement | “Thank you for agreeing to participate in this survey. My name is ______. I will be asking you the questions. Most of the questions require that you select one of the options as your answer, although some questions you can select all the answers that apply. I will specify the options and instructions for you as I ask each question. If your answer is not one of the specified options please tell me and I will write your answer down. Please feel free to tell me whatever you are comfortable sharing. You should also remember that you do not have to share anything that you are not comfortable sharing and that you can stop this interview at any time without any risk to your rights or treatment and care. There are no right or wrong answers, so please be honest and help us to understand what is true for you and your community. Are you ready to begin?<br>*Read the following statement. Please repeat the statement translated into the local language based on primary languages.* | |
| V. Patient survey > Respondent demographics and socio-economic status | | |
| demographics_statement | “I’m going to start by asking you some basic questions about who you are, where you live, and your education and employment.” | |
| nationality *(required)* | 1. What is your nationality/country of origin? | 1 South Africa<br>2 Botswana<br>3 Lesotho<br>4 Mozambique<br>5 Malawi<br>6 Namibia<br>7 Swaziland<br>8 Zambia<br>9 Zimbabwe<br>10 Tanzania |

| Field | Question | Answer |
|---|---|---|
| | | 11 Burundi<br>12 Other African country (specify)<br>13 Other (specify) |
| other_nationality *(required)* | Please specify nationality/country of origin | |
| years_in_sa *(required)* | 2. If you are not South African, how long have you lived in South Africa? | 1 < 1 yr<br>2 1-2 years<br>3 2-5 years<br>4 >5 years<br>5 Seasonal work (Only work in South Africa during certain times of the year) |
| marital_status *(required)* | 3. What is your marital status? | 1 Never married<br>2 Married (customary/traditional or legal/civil)<br>3 Divorced<br>4 Separated<br>5 Widowed |
| have_a_partner *(required)* | 4. Is there someone who you have a relationship with and who you call your partner? | 1 Yes<br>0 No |
| living_with_spouse *(required)* | 5. Do you currently live with your husband/wife or your partner? | 1 No<br>2 Yes, married or living together |
| current_household *(required)* | 6. Do you think of the house you currently live in as your main house? | 1 Yes<br>2 No, my main house is somewhere else in South Africa<br>3 No, my main house is in another country |
| V. Patient survey > Respondent demographics and socio-economic status > Education | | |
| reading_ability *(required)* | 7. Do you know how to read and write? | 1 No<br>2 Yes – read and write<br>3 Yes – read only |
| edu_level *(required)* | 8. What was the highest level of school that you completed? | 1 No schooling<br>2 Primary<br>3 Secondary<br>4 Certificate/Diploma/ Post-secondary<br>5 Graduate degree |
| occupation *(required)* | 9. What is your occupation? | 1 Farming (my own or my family's farm)<br>2 Farm worker (someone else's farm)<br>3 Domestic worker or carer (paid)<br>4 Informal sector job (not farming or domestic) (e.g. trader, day service provider)<br>5 Formal sector job (salaried)<br>6 Household work and/or childcare (my own house, not paid)<br>7 Unemployed but looking for work<br>8 Student or trainee<br>9 Retired<br>11 Self-employed/own business<br>12 Unemployed but not looking for work<br>10 Other (specify) |
| other_occupation *(required)* | Please specify your occupation | |

| Field | Question | Answer |
|---|---|---|
| mostmoney *(required)* | 10. Where do you get MOST of your money from? | 1 Salary, business or job (formal or informal sector)<br>2 Government social grant<br>3 Spouse/partner<br>4 Parents/relatives<br>5 Friends<br>6 Other (specify) |
| specifymoney *(required)* | Please specify | |
| lack_of_food *(required)* | 11. Do you or the people in your household go without food often, sometimes, seldom, never? | 1 Never<br>2 Seldom<br>3 Sometimes<br>4 Often |
| government_grant *(required)* | 12. Do you or does anybody in your household, currently receive any support or grant from the government?<br>*Tick all that apply* | 0 No<br>1 Child grant<br>2 Partial disability / illness grant / temporary grant<br>3 Pension grant<br>4 Disability grant<br>5 Unemployment grant/UIF<br>7 COVID social relief grant<br>6 Other (specify) |
| other_support_grant *(required)* | Please specify the support or grant from the government | |
| healthcare_money *(required)* | 13. If a person in your household became ill and 100 Rands was needed for treatment or medicines, would you say it would be very easy, easy, difficult, or very difficult to find the money? | 1 Very difficult<br>2 Difficult<br>3 Easy<br>4 Very easy |
| V. Patient survey > Healthcare access and cost | | |
| V. Patient survey > Healthcare access and cost > ART questions | | |
| HIVtreatment_duration *(required)* | 14. How long have you been taking ART?<br>*Enter years* | |
| HIVtreatment_duration_months *(required)* | Surveyor: Now enter the number of months<br>*Months* | |
| months_for_med *(required)* | 15. On average, how many months of ART medication do you receive at a time?<br>*Months* | |
| additionaldiseases *(required)* | 16. Do you have other conditions/illnesses in addition to HIV? | 1 Yes<br>0 No |
| additionalservices *(required)* | 17. Which health care services are you routinely receiving at this facility, in addition to HIV care?<br>*(Tick all that apply)* | 1 TB<br>2 Diabetes<br>3 Hypertension<br>4 Asthma<br>5 Mental health<br>6 Malaria<br>7 Family planning<br>8 Antenatal care<br>9 Child health care<br>10 TB preventative therapy (TPT)<br>11 Other (specify) |
| specifyaddservices *(required)* | Please specify | |
| V. Patient survey > Healthcare access and cost > Follow-up questions ([additionalservices_name]) (1) | | (Repeated group) |
| receivettreatment *(required)* | 18. Do you receive treatment for [additionalservices_name] or as part of the health care services you are routinely receiving at this facility? | 1 Yes<br>0 No |
| combinevisits *(required)* | 19. Are you able to combine your HIV visits with the visits for [additionalservices_name]? | 0 Always<br>1 Very often<br>2 Sometimes<br>3 Rarely<br>4 Never |
| collectforall *(required)* | 20. Are you able to collect your medication for [additionalservices_name] at the same time as you collect your HIV medication? | 0 Always<br>1 Very often<br>2 Sometimes<br>3 Rarely<br>4 Never |

| Field | Question | Answer |
|---|---|---|
| healthcare_professional_visited *(required)* | 21. In the past 12 months have you sought health care from any other health care provider outside this facility? *Tick all that apply* | 0 No<br>1 Hospital<br>2 Private doctor<br>3 Traditional healer<br>4 Community health worker<br>5 Local NGO/FBO<br>6 Other (specify) |
| other_health_providers_visited *(required)* | Please specify other health care providers you sought health care from any outside this facility | |
| payforservice *(required)* | 22. Did you have to pay for the service? | 1 Yes<br>0 No |
| medicalaid *(required)* | 23. Are you covered by Medical Aid, Private health insurance, Medical Benefit Scheme, Provident Scheme, or Hospital Plan that helps you pay for health care or medication? | 1 Yes<br>0 No |
| transport_to_clinic *(required)* | 24. How do you usually get to the clinic? *Tick all that apply* | 1 Walk<br>2 Mini-bus/common taxi<br>3 Own car<br>4 Meter taxi/Uber/Taxify/Hired taxi<br>5 Brought by family/friends in their vehicles<br>6 Other (specify) |
| other_transport_to_clinic *(required)* | Please specify other means of getting to the clinic | |
| V. Patient survey > Healthcare access and cost > Get to clinic | | |
| travel_time *(required)* | 25. How long does it take you to get to the clinic? (One way – from home to the clinic) *Enter hours* | |
| travel_time_minutes *(required)* | Surveyor: Now enter minutes *Minutes* | |
| otherwisedoing *(required)* | 26. What would you otherwise have been doing if you had not come to the clinic today? | 1 Housework<br>2 Childcare (own children)<br>3 Caring for a relative or friend<br>4 Voluntary work<br>5 Leisure activities<br>6 Attending school or university<br>7 On sick leave<br>8 Seeking work<br>9 Paid work<br>10 Other (specify) |
| specifyotherwise *(required)* | Please specify | |
| expenses_incurred *(required)* | 27. What expenses/costs do you incur for each clinic visit? | 0 No costs<br>1 Transport<br>2 Loss of income due to missing work<br>3 Child care<br>4 Food/drinks<br>5 Other (specify) |
| other_expenses *(required)* | Please specify other expenses/costs you incur for each clinic visit | |
| transport_costs *(required)* | 28. Please estimate how much does public transport cost you in Rands each time you visit the clinic (Return trip – to the clinic and back home) *Amount in Rands* | |
| unpaid_time_costs *(required)* | 29. Please estimate how much does unpaid time off work cost you in Rands each time you visit the clinic *Amount in Rands* | |
| childcarecosts *(required)* | 30. Please estimate how much does child care cost you in Rands each time you visit the clinic *Amount in Rands* | |
| foodcosts *(required)* | 31. Please estimate how much food/drinks cost you in Rands each time you visit the clinic *Amount in Rands* | |
| othercosts *(required)* | 32. Please estimate how much does the 'other specified' cost you in Rands each time you visit the clinic *Amount in Rands* | |
| V. Patient survey > Healthcare access and cost > How many visits | | |
| nurse_and_collection_visit *(required)* | 33. For your HIV treatment, how many visits to this clinic where you both see a nurse and collect your medication do you attend per year? *Number of visits per year* | |
| nurse_and_collection_hours *(required)* | 34. How long does it take total, on average for each HIV clinic visit where you see a nurse and pick up medications (counting from when you arrive at the clinic to when you leave)? *Enter hours* | |
| nurse_and_collection_minutes *(required)* | Surveyor: Now enter minutes | |

| Field | Question | Answer |
|---|---|---|
| | *Minutes* | |
| med_collection_visit_only *(required)* | 35. For your HIV treatment, how many visits to this clinic where you do NOT see a nurse but do collect your medication do you attend per year?<br>*Number of visits per year* | |
| med_collection_hours *(required)* | 36. How long does it take total, on average for each ART medication pick-up (visits where you only pick up medications, do not see a nurse)?<br>*Enter hours* | |
| med_collection_minutes *(required)* | Surveyor: Now enter minutes<br>*Minutes* | |
| missed_visits *(required)* | 37. Have there been occasions where you missed your facility visits in the past year by more than 7 days? | 1 Yes<br>0 No |
| missed_visits_no *(required)* | 38. How many facility visits have you missed by more than 7 days?<br>*Visits missed in the past year* | |
| missed_visit_reason *(required)* | 39. Thinking of the most recent facility visit you missed, what was the MAIN reason you missed the visit? | 1 Forgot pick up date<br>2 Ill health<br>3 Nobody else to go for me<br>4 Buddy forgot<br>5 Buddy unwell<br>6 No money for transport<br>7 Could not leave work<br>8 Afraid HIV status will get known<br>10 Travelling/away from home<br>11 My appointment date/time was no longer convenient<br>9 Other (specify) |
| other_missed_visit_reasons *(required)* | Please specify other reasons for missing the visit | |
| stillhaveart *(required)* | 40. Did you still have ART medication in hand even though you missed your facility visit? | 0 No<br>1 No, but I was able to collect ART medication elsewhere<br>2 Yes, I still had ART medication |
| specifylocation *(required)* | Specify where you collected the medication | |
| facilitycall *(required)* | 41. Did someone from the facility call/message/visit you after you missed the facility visit? | 1 Yes<br>0 No |
| feelsick *(required)* | 42. What do you do if you feel sick or have questions related to your healthcare when you do not have a scheduled visit? | 1 Make a special visit to this clinic<br>2 Wait until my scheduled visit to this clinic<br>3 Contact a healthcare provider via phone<br>4 Other (specify) |
| specifyfeelsick *(required)* | Please specify | |
| treatmentquestions *(required)* | 43. If you have a question about your HIV treatment, what would you most likely do? | 1 Make a special visit to this clinic<br>2 Wait and ask during a regular visit to this clinic<br>3 Contact a healthcare provider via phone<br>4 Other (specify) |
| specifytreatmentquestions *(required)* | Please specify | |
| covidchanges *(required)* | 44. Did you experience any changes in how you received care for your HIV during the COVID pandemic? | 1 Yes<br>0 No |
| whatchanged *(required)* | 45. What changed?<br>*(Tick all that apply)* | 1 Staff shortages<br>2 Longer waiting times<br>3 Medication stockout<br>4 Clinic was closed for a period of time<br>5 Turned away without receiving services because of the reduced number of patients the facility could attend to because of social distancing |

| Field | Question | Answer |
| --- | --- | --- |
| | | 6 I received more months of medication<br>7 I received a 12-month script<br>8 Offered a different, more convenient medication collection location<br>9 I was offered home ART delivery<br>10 Other (specify) |
| specifychange *(required)* | Please specify | |
| V. Patient survey > Questions for patients in standard care | | |
| ever_enrolled_in_DSD *(required)* | 46. Have you ever been enrolled in a DSD model such as Adherence Club, Ex-PuP, Fac-PuP and etc.? | 0 No<br>1 Yes<br>2 Don't know |
| which_model *(required)* | 47. Which DSD model were you enrolled in? | 1 Adherence club<br>2 Facility Pick Up Point<br>3 External Pick Up Point<br>4 Youth club<br>5 Pele Box<br>7 Home ART delivery<br>8 Bicycle model<br>9 Dablap<br>10 Sha'p left<br>6 Other |
| specify_model *(required)* | Please specify the model you were enrolled in. | |
| back_referral_reasons *(required)* | 48. Can you recall the reasons you were referred back to regular (standard of care) HIV services by the model's staff?<br>*(Tick all that apply)* | 0 No<br>1 Ill health<br>2 Missed a visit<br>3 Missed ARV doses<br>4 Blood draw<br>5 Screened positive for TB<br>6 Diabetes complication<br>7 Hypertension complication<br>8 Don't know why<br>9 Other |
| specifybackreferral *(required)* | Please specify | |
| notenrolled *(required)* | 49. What is the reason you are currently not enrolled in a DSD model? | 1 I prefer standard of care (attending normal clinic visits)<br>2 The clinic is more convenient for me<br>3 I was never offered/ given a choice<br>4 I have another condition for which I need to come to the clinic (specify)<br>5 I don't know<br>6 Other (specify) |
| specifynotenrolled *(required)* | Please specify | |
| V. Patient survey > Questions for patients in standard care > 50 a. Accessibility /convenience | | |
| labelconvenience | a. Accessibility /convenience (How convenient to you are the following?): | 1 very inconvenient<br>2 inconvenient<br>3 neither convenient nor inconvenient<br>4 convenient<br>5 very convenient |
| refillplace *(required)* | i. Place you go for your drug refill | 1 very inconvenient<br>2 inconvenient<br>3 neither convenient nor inconvenient<br>4 convenient<br>5 very convenient |

| Field | Question | Answer |
| --- | --- | --- |
| dayoftheweek *(required)* | ii. Day of the week you are scheduled to pick up your drugs | 1 very inconvenient<br>2 inconvenient<br>3 neither convenient nor inconvenient<br>4 convenient<br>5 very convenient |
| timeoftheday *(required)* | iii. Time of the day you pick up your drugs | 1 very inconvenient<br>2 inconvenient<br>3 neither convenient nor inconvenient<br>4 convenient<br>5 very convenient |
| placeforreview *(required)* | iv. Place for the clinical consultation | 1 very inconvenient<br>2 inconvenient<br>3 neither convenient nor inconvenient<br>4 convenient<br>5 very convenient |
| dayforreview *(required)* | v. Day of the week you are scheduled for your clinical consultation | 1 very inconvenient<br>2 inconvenient<br>3 neither convenient nor inconvenient<br>4 convenient<br>5 very convenient |
| V. Patient survey > Questions for patients in standard care > 50 b. Environment | | |
| howgood | b. Environment (Tell me how good the following at this facility) are: | 1 very good<br>2 good<br>3 fair<br>4 poor<br>5 very poor |
| neatness *(required)* | vi. Neatness and cleanliness | 1 very good<br>2 good<br>3 fair<br>4 poor<br>5 very poor |
| ease *(required)* | vii. Ease of finding where to go (clinic flow) | 1 very good<br>2 good<br>3 fair<br>4 poor<br>5 very poor |
| privacy *(required)* | viii. Privacy | 1 very good<br>2 good<br>3 fair<br>4 poor<br>5 very poor |
| V. Patient survey > Questions for patients in standard care > 50 c. Efficiency | | |
| howsatisfied | c. Efficiency (how satisfied are the following aspects of your HIV care?): | 1 very inefficient<br>2 inefficient<br>3 neither efficient nor inefficient<br>4 efficient<br>5 very efficient |
| waitingtime *(required)* | ix. Waiting time to see a clinician | 1 very inefficient<br>2 inefficient<br>3 neither efficient nor inefficient<br>4 efficient<br>5 very efficient |
| collectiontime *(required)* | x. Waiting time to collect medication | 1 very inefficient<br>2 inefficient<br>3 neither efficient nor inefficient<br>4 efficient<br>5 very efficient |

| Field | Question | Answer |
|---|---|---|
| timewithprovider *(required)* | xi. Time spent with health provider | 1 very inefficient<br>2 inefficient<br>3 neither efficient nor inefficient<br>4 efficient<br>5 very efficient |
| costincurred *(required)* | xii. Cost incurred to attend visit | 1 very inefficient<br>2 inefficient<br>3 neither efficient nor inefficient<br>4 efficient<br>5 very efficient |
| V. Patient survey > Questions for patients in standard care > 50 d. Comprehensiveness | | |
| comprehensiveness | d. Comprehensiveness (how comprehensive were the following aspects of your HIV care?): | 1 very non-comprehensive<br>2 non-comprehensive<br>3 neither non-comprehensive nor comprehensive<br>4 comprehensive<br>5 very comprehensive |
| healthedureceipt *(required)* | xiii. Receipt of health education | 1 very non-comprehensive<br>2 non-comprehensive<br>3 neither non-comprehensive nor comprehensive<br>4 comprehensive<br>5 very comprehensive |
| counsellingreceipt *(required)* | xiv. Receipt of counselling | 1 very non-comprehensive<br>2 non-comprehensive<br>3 neither non-comprehensive nor comprehensive<br>4 comprehensive<br>5 very comprehensive |
| visitsrequired *(required)* | xv. The number of visits you are required to attend | 1 very non-comprehensive<br>2 non-comprehensive<br>3 neither non-comprehensive nor comprehensive<br>4 comprehensive<br>5 very comprehensive |
| V. Patient survey > Questions for patients in standard care > 50 e. Humaneness | | |
| humaneness | e. Humaneness (how satisfied are you with the following aspects relate to your interactions with your HIV care health provider?): | 1 very dissatisfied<br>2 dissatisfied<br>3 neither satisfied nor dissatisfied<br>4 satisfied<br>5 very satisfied |
| stafffriendliness *(required)* | xvi. Staff friendliness towards you | 1 very dissatisfied<br>2 dissatisfied<br>3 neither satisfied nor dissatisfied<br>4 satisfied<br>5 very satisfied |
| confidentiality *(required)* | xvii. Confidentiality | 1 very dissatisfied<br>2 dissatisfied<br>3 neither satisfied nor dissatisfied<br>4 satisfied<br>5 very satisfied |
| staffrespectfulness *(required)* | xviii. Respectfulness of staff | 1 very dissatisfied<br>2 dissatisfied<br>3 neither satisfied nor dissatisfied<br>4 satisfied<br>5 very satisfied |

| Field | Question | Answer |
| --- | --- | --- |
| staffattentiveness *(required)* | xix. Staff's attentiveness during interaction | 1 very dissatisfied<br>2 dissatisfied<br>3 neither satisfied nor dissatisfied<br>4 satisfied<br>5 very satisfied |
| opportunitytoask *(required)* | xx. Extent to which staff give client opportunity to ask questions | 1 very dissatisfied<br>2 dissatisfied<br>3 neither satisfied nor dissatisfied<br>4 satisfied<br>5 very satisfied |
| decisionmaking *(required)* | xxi. Extent to which staff involved client in decision-making about your healthcare | 1 very dissatisfied<br>2 dissatisfied<br>3 neither satisfied nor dissatisfied<br>4 satisfied<br>5 very satisfied |
| V. Patient survey > Questions for patients in standard care > Satisfaction rating | | |
| overall_satisfaction *(required)* | 51. How would you rate your overall satisfaction with the care that you receive at this facility? | 1 very dissatisfied<br>2 dissatisfied<br>3 neither satisfied nor dissatisfied<br>4 satisfied<br>5 very satisfied |
| overall_satisfaction_explained *(required)* | 52. Please explain your satisfaction rating above | |
| reasonabletime *(required)* | 53. What do you think would be a reasonable length of time to wait from arriving at the facility before receiving care from a health provider?<br>*Enter hours* | |
| reasonableminutes *(required)* | Surveyor: Now enter the minutes | |
| waittoreceivecare *(required)* | 54. How long do you usually wait from arriving at the facility to receive care from a health provider?<br>*Enter hours* | |
| waittoreceivecaremin *(required)* | Surveyor: Now enter the minutes | |
| disappointed *(required)* | 55. Were you disappointed with any aspect of your care at this facility? | |
| service_improvement *(required)* | 56. How could HIV services in this facility be improved?<br>*(select all that apply)* | 1 More staff<br>2 More information provided by staff<br>3 Better, more polite, or friendlier nurse and counselor attitude/manner<br>4 Better, more polite, or friendlier reception and admin attitude/manner<br>5 Better location<br>6 Open different days<br>7 Open different times of day<br>8 Open outside of work hours<br>9 Shorter waiting time<br>10 More counselling when there are problems<br>11 More counselling overall<br>12 Less counselling<br>13 Being able to pick up ARVs at different and more convenient sites<br>14 Being able to pick up ARVs at more convenient times<br>15 Being able to have someone else pick up your ARVs<br>16 Having somebody to support you take your ARVs |

| Field | Question | Answer |
|---|---|---|
| | | 17 Tracing when missed an appointment<br>18 Reminders via phone<br>19 More months of ARVS given at each visit<br>20 Fewer months of ARVS given at each visit<br>21 Better access to a nurse or clinic staff<br>22 Treatment/support for other illnesses (specify)<br>23 Other (specify and elaborate ) |
| other_service_improvements *(required)* | Please specify other ways HIV services can be improved | |
| V. Patient survey > Questions for patients in DSD models | | |
| first_enrollment *(required)* | 57. Can you recall the first time you were enrolled in this model? | 1 Yes<br>0 No |
| when *(required)* | 58. When were you first enrolled? | |
| V. Patient survey > Questions for patients in DSD models > How long on ART | | |
| howlongonart *(required)* | 59. How long had you been on ART when you enrolled in this model?<br>*Enter years* | |
| onartmonths *(required)* | Surveyor: Now enter the number of months | |
| ask_to_enroll *(required)* | 60. Did you ask to be enrolled in this model? | 1 Yes<br>0 No |
| consent_to_enroll *(required)* | 61. Did you have to provide consent to be enrolled in this model? | 0 No<br>1 Yes – written consent<br>2 Yes – verbal consent<br>99 Not sure/don't know/can't remember |
| choice_to_enroll *(required)* | 62. Were you given a choice about joining this model? | 0 No<br>1 Yes<br>99 Not sure/don't know/can't remember |
| whatwasthechoice *(required)* | 63. What was the choice you were given | |
| whychoosetojoin *(required)* | 64. Why did you choose to join this model? | |
| counsellingwhenjoining *(required)* | 65. Do you receive any counselling, education, and/or adherence support before or at the time you joined this model? | 1 Yes<br>0 No |
| describecounselling *(required)* | 66. Please describe | |
| understandthemodel *(required)* | 67. When you joined this model, did you understand the differences between your new model and the previous model or the care you received before you joined the model? | 1 Yes<br>0 No |
| whatyouunderstood *(required)* | 68. Please explain what you understood about the care you would receive under this model | |
| happy_to_enroll *(required)* | 69. Were you happy to be enrolled in a DSD model? | 0 No<br>1 Somewhat<br>2 Yes<br>3 Neither happy nor unhappy (did not care) |
| discussmodel *(required)* | 70. When last did you discuss your model choice with your provider? | 1 Never<br>2 When I first enrolled in my current model<br>3 During my last clinical visit<br>4 At my last drug pick-up<br>5 Other (specify) |
| specifydiscussmodel *(required)* | Please specify | |
| awareofmodels *(required)* | 71. Are you aware of other DSD models currently available at this facility, and what would be your preferred model? | 1 Adherence club<br>2 Facility Pick Up Point<br>3 External Pick Up Point<br>4 Youth club<br>5 Pele Box<br>7 Home ART delivery<br>8 Bicycle model<br>9 Dablap<br>10 Sha'p left |

| Field | Question | Answer |
|---|---|---|
| | | 6 Other |
| specifyawareofmodels *(required)* | Please specify | |
| preferredmodelreason *(required)* | 72. Please elaborate on the reasons for your preferred model of care | |
| V. Patient survey > Questions for patients in DSD models > 73 a. Accessibility /convenience | | |
| labelconvenience2 | a. Accessibility /convenience (How convenient to you are the following?): | 1 very inconvenient<br>2 inconvenient<br>3 neither convenient nor inconvenient<br>4 convenient<br>5 very convenient |
| refillplace2 *(required)* | i. Place you go for your drug refill | 1 very inconvenient<br>2 inconvenient<br>3 neither convenient nor inconvenient<br>4 convenient<br>5 very convenient |
| dayoftheweek2 *(required)* | ii. Day of the week you are scheduled to pick up your drugs | 1 very inconvenient<br>2 inconvenient<br>3 neither convenient nor inconvenient<br>4 convenient<br>5 very convenient |
| timeoftheday2 *(required)* | iii. Time of the day you pick up your drugs | 1 very inconvenient<br>2 inconvenient<br>3 neither convenient nor inconvenient<br>4 convenient<br>5 very convenient |
| placeforreview2 *(required)* | iv. Place for the clinical consultation | 1 very inconvenient<br>2 inconvenient<br>3 neither convenient nor inconvenient<br>4 convenient<br>5 very convenient |
| dayforreview2 *(required)* | v. Day of the week you are scheduled for your clinical consultation | 1 very inconvenient<br>2 inconvenient<br>3 neither convenient nor inconvenient<br>4 convenient<br>5 very convenient |
| V. Patient survey > Questions for patients in DSD models > 73 b. Environment | | |
| howgood2 | b. Environment (Tell me how good the following at this facility) are: | 1 very good<br>2 good<br>3 fair<br>4 poor<br>5 very poor |
| neatness2 *(required)* | vi. Neatness and cleanliness | 1 very good<br>2 good<br>3 fair<br>4 poor<br>5 very poor |
| ease2 *(required)* | vii. Ease of finding where to go (clinic flow) | 1 very good<br>2 good<br>3 fair<br>4 poor<br>5 very poor |
| privacy2 *(required)* | viii. Privacy | 1 very good<br>2 good<br>3 fair<br>4 poor<br>5 very poor |

5 very poor

| Field | Question | Answer |
|---|---|---|
| V. Patient survey > Questions for patients in DSD models > 73 c. Efficiency | | |
| howsatisfied2 | c. Efficiency (how efficient are the following aspects of your HIV care?): | 1 very inefficient<br>2 inefficient<br>3 neither efficient nor inefficient<br>4 efficient<br>5 very efficient |
| waitingtime2 *(required)* | ix. Waiting time to see a clinician | 1 very inefficient<br>2 inefficient<br>3 neither efficient nor inefficient<br>4 efficient<br>5 very efficient |
| collectiontime2 *(required)* | x. Waiting time to collect medication | 1 very inefficient<br>2 inefficient<br>3 neither efficient nor inefficient<br>4 efficient<br>5 very efficient |
| timewithprovider2 *(required)* | xi. Time spent with health provider | 1 very inefficient<br>2 inefficient<br>3 neither efficient nor inefficient<br>4 efficient<br>5 very efficient |
| costincurred2 *(required)* | xii. Cost incurred to attend visit | 1 very inefficient<br>2 inefficient<br>3 neither efficient nor inefficient<br>4 efficient<br>5 very efficient |
| V. Patient survey > Questions for patients in DSD models > 73 d. Comprehensiveness | | |
| comprehensiveness2 | d. Comprehensiveness (how comprehensive were the following aspects related to your HIV care?): | 1 very non-comprehensive<br>2 non-comprehensive<br>3 neither non-comprehensive nor comprehensive<br>4 comprehensive<br>5 very comprehensive |
| healthedureceipt2 *(required)* | xiii. Receipt of health education | 1 very non-comprehensive<br>2 non-comprehensive<br>3 neither non-comprehensive nor comprehensive<br>4 comprehensive<br>5 very comprehensive |
| counsellingreceipt2 *(required)* | xiv. Receipt of counselling | 1 very non-comprehensive<br>2 non-comprehensive<br>3 neither non-comprehensive nor comprehensive<br>4 comprehensive<br>5 very comprehensive |
| visitsrequired2 *(required)* | xv. The number of visits you are required to attend | 1 very non-comprehensive<br>2 non-comprehensive<br>3 neither non-comprehensive nor comprehensive<br>4 comprehensive<br>5 very comprehensive |
| V. Patient survey > Questions for patients in DSD models > 73 e. Humaneness | | |
| humaneness2 | e. Humaneness (how satisfied are you with the following aspects related to your interactions with your HIV care health provider?): | 1 very dissatisfied<br>2 dissatisfied<br>3 neither satisfied nor dissatisfied<br>4 satisfied<br>5 very satisfied |
| stafffriendliness2 *(required)* | xv.i Staff friendliness towards you | 1 very dissatisfied<br>2 dissatisfied |

| Field | Question | Answer |
|---|---|---|
| | | 3 neither satisfied nor dissatisfied<br>4 satisfied<br>5 very satisfied |
| confidentiality2 *(required)* | xvii. Confidentiality | 1 very dissatisfied<br>2 dissatisfied<br>3 neither satisfied nor dissatisfied<br>4 satisfied<br>5 very satisfied |
| staffrespectfulness2 *(required)* | xviii. Respectfulness of staff | 1 very dissatisfied<br>2 dissatisfied<br>3 neither satisfied nor dissatisfied<br>4 satisfied<br>5 very satisfied |
| staffattentiveness2 *(required)* | xix. Staff's attentiveness during interaction | 1 very dissatisfied<br>2 dissatisfied<br>3 neither satisfied nor dissatisfied<br>4 satisfied<br>5 very satisfied |
| opportunitytoask2 *(required)* | xx. Extent to which staff give client opportunity to ask questions | 1 very dissatisfied<br>2 dissatisfied<br>3 neither satisfied nor dissatisfied<br>4 satisfied<br>5 very satisfied |
| decisionmaking2 *(required)* | xxi. Extent to which staff involved client in decision-making about your healthcare | 1 very dissatisfied<br>2 dissatisfied<br>3 neither satisfied nor dissatisfied<br>4 satisfied<br>5 very satisfied |
| V. Patient survey > Questions for patients in DSD models > Satisfaction rating | | |
| satisfaction_before_model *(required)* | 74. How would you rate your overall satisfaction with your ART care BEFORE you started this model? | 1 Extremely dissatisfied<br>2 Not satisfied<br>3 Neither satisfied nor dissatisfied<br>4 Satisfied<br>5 Very satisfied |
| satisfaction_during_model *(required)* | 75. How would you rate your overall satisfaction with your ART care now that you are in this model? | 1 Extremely dissatisfied<br>2 Not satisfied<br>3 Neither satisfied nor dissatisfied<br>4 Satisfied<br>5 Very satisfied |
| model_satisfaction_rating *(required)* | 76. Please explain your satisfaction rating for this model | |
| reasonabletime_dsd *(required)* | 77. What do you think would be a reasonable length of time to wait from arriving at the clinic before receiving care from a health provider?<br>*Enter hours* | |
| reasonableminutes_dsd *(required)* | Surveyor: Now enter the minutes | |
| waittoreceivecare_dsd *(required)* | 78. How long do you usually wait from arriving at the clinic to receive care from a health provider?<br>*Enter hours* | |
| waittoreceivecaremin_dsd *(required)* | Surveyor: Now enter the minutes | |
| q51_skip | 79. Do you attend any out-of-facility HIV events as part of your HIV care? By "event" we mean anything involving your treatment that is not at this facility, including club meetings, picking up medications outside the clinic, etc. | 1 Yes<br>0 No |
| out_of_facility_events *(required)* | 80. How many out-of-facility HIV events do you attend per year? By "event" we mean anything involving your treatment that is not at this facility, including club meetings, picking up medications outside the clinic, etc.<br>*Number/year* | |
| transport_to_model_event *(required)* | 81. How do you usually get to the out-of-facility model events?<br>*Tick all that apply* | 1 Walk<br>2 Mini-bus/common taxi |

| Field | Question | Answer |
|---|---|---|
| | | 3 Own car<br>4 Meter taxi/Uber/Taxify/Hired taxi<br>5 Brought by family/friends in their vehicles<br>6 Other (specify) |
| other_travel_to_model_event *(required)* | Please specify other ways you get to the out-of-facility model events | |
| V. Patient survey > Questions for patients in DSD models > Travel time | | |
| travel_dsd_hours *(required)* | 82. How long does it take you to travel to the out-of-facility model events? (One way – from home to the clinic)<br>*Enter hours* | |
| travel_dsd_minutes *(required)* | Surveyor: Now enter minutes<br>*Minutes* | |
| otherwisedoing_dsd *(required)* | 83. What would you otherwise have been doing if you had not attended out-of-facility model events? | 1 Housework<br>2 Childcare (own children)<br>3 Caring for a relative or friend<br>4 Voluntary work<br>5 Leisure activities<br>6 Attending school or university<br>7 On sick leave<br>8 Seeking work<br>9 Paid work<br>10 Other (specify) |
| specifyotherwisedoing_dsd *(required)* | Please specify | |
| expenses_incurred_dsd *(required)* | 84. What expenses/costs do you incur for each out-of-facility model event? | 0 No costs<br>1 Transport<br>2 Loss of income due to missing work<br>3 Child care<br>4 Food/drinks<br>5 Other (specify) |
| other_expenses_incurred_dsd *(required)* | Please specify other expenses/costs you incur for each out-of-facility model event | |
| transport_costs_dsd *(required)* | 85. Please estimate how much does transport cost you in Rands each time you make an out-of-facility model visit.<br>(Return trip – to the clinic and back home)<br>*Amount in Rands* | |
| unpaid_time_costs_dsd *(required)* | 86. Please estimate how much income (wages or salary) you lose in Rands each time you attend a model event.<br>*Amount in Rands* | |
| childcarecosts_dsd *(required)* | 87. Please estimate how much child care costs you in Rands each time you attend an out-of-facility event<br>*Amount in Rands* | |
| foodcosts_dsd *(required)* | 88. Please estimate how much food/drinks cost you in Rands each time you attend an out-of-facility event<br>*Amount in Rands* | |
| othercosts_dsd *(required)* | Please estimate how much does the 'other specified' cost you in Rands each time you attend an out-of-facility event<br>*Amount in Rands* | |
| V. Patient survey > Questions for patients in DSD models > Out of facility subgroup | | |
| dsd_event_hours *(required)* | 89. How long does it take total, on average for each out-of-facility model event (Hours and minutes, including travel time and time at event)<br>*Enter hours* | |
| dsd_event_minutes *(required)* | Surveyor: Now enter minutes<br>*Minutes* | |
| overall_satisfaction_dsd *(required)* | 90. How would you rate your overall satisfaction with your out-of-facility model events? | 1 Extremely dissatisfied<br>2 A little dissatisfied<br>3 Neither satisfied nor dissatisfied<br>4 Satisfied<br>5 Very satisfied |
| explainsatisfaction *(required)* | 91. Please explain your satisfaction rating for your out-of-facility model events | |
| reasonabletime_outoffacility *(required)* | 92. What do you think would be a reasonable length of time to wait from arriving at the out-of-facility model event before receiving care from a health provider?<br>*Enter hours* | |
| reasonableminutes_outoffacility *(required)* | Surveyor: Now enter the minutes | |
| waittoreceivecare_outoffacility *(required)* | 93. How long do you usually wait from arriving at the out-of-facility model event to receive care from a health provider?<br>*Enter hours* | |
| waittoreceivecaremin_outoffacility *(required)* | Surveyor: Now enter the minutes | |

| Field | Question | Answer |
|---|---|---|
| referred_back *(required)* | 94. Have you been referred back to regular (standard of care) HIV services by this model's staff for any reason? *(Tick all that apply)* | 0 No<br>1 Ill health<br>2 Missed a visit<br>3 Missed ARV doses<br>4 Blood draw<br>5 Screened positive for TB<br>6 Diabetes complication<br>7 Hypertension complication<br>8 Don't know why<br>9 Other (specify) |
| specify_referred_back *(required)* | Specify the reason you were referred back | |
| differentmodel *(required)* | 95. Since the first time you were enrolled in a DSD model, have you ever been enrolled in a different model than you are currently? | 1 Yes<br>0 No |
| previousmodels *(required)* | 96. Which model/s were you previously enrolled in? *(tick-all that apply)* | 1 Adherence club<br>2 Facility Pick Up Point<br>3 External Pick Up Point<br>4 Youth club<br>5 Pele Box<br>7 Home ART delivery<br>8 Bicycle model<br>9 Dablap<br>10 Sha'p left<br>6 Other |
| specifypreviousmodels *(required)* | Please specify | |
| whychangemodels *(required)* | 97. What was the reason you changed models? | 1 Pregnancy<br>2 End of pregnancy or postpartum period<br>3 My age meant I was eligible for a different model for older individuals<br>4 I moved houses, or my place of work changed<br>5 My VL was elevated (I was unwell)<br>6 My VL became suppressed<br>7 I had stopped treatment<br>8 The model is no longer offered at the facility<br>9 I preferred to be in a different model<br>10 Other (specify) |
| specifywhychangedmodels *(required)* | Please specify | |
| missed_visits_dsd *(required)* | 98. Have there been occasions in the past year where you missed your out-of-facility model event by more than 7 days? | 1 Yes<br>0 No |
| missed_events *(required)* | 99. How many out-of-facility model events have you missed by more than 7 days? *Specify number* | |
| missed_visit_reason_dsd *(required)* | 100. Why did you miss the out-of-facility model event? | 1 Forgot pick up date<br>2 Ill health<br>3 Nobody else to go for me<br>4 Buddy forgot<br>5 Buddy unwell<br>6 No money for transport<br>7 Could not leave work<br>8 Afraid HIV status will get known<br>10 Travelling/away from home<br>11 My appointment date/time was no longer convenient<br>9 Other (specify) |
| other_missed_visit_reasons_dsd *(required)* | Please specify | |
| stillhaveart_dsd *(required)* | 101. Did you still have ART medication in hand even though you missed your out-of-facility model event? | 0 No |

| Field | Question | Answer |
| --- | --- | --- |
| | | 1 No, but I was able to collect ART medication elsewhere<br>2 Yes, I still had ART medication |
| facilitycall_dsd *(required)* | 102. Did someone from the facility call/message/visit you after you missed the out-of-facility model event? | 1 Yes<br>0 No |
| feelsick_outoffacility *(required)* | 103. What do you do if you feel sick or have questions related to your healthcare when you do not have a scheduled visit? | 1 Make a special visit to this clinic<br>2 Wait until my scheduled visit to this clinic<br>3 Wait until my scheduled out-of-facility HIV event<br>4 Contact a healthcare provider via phone<br>5 Other (specify) |
| specifyfeelsick_outoffacility *(required)* | Please specify | |
| treatment_questions *(required)* | 104. If you have a question about your HIV treatment, what would you most likely do? | 1 Make a special visit to this clinic<br>2 Wait and ask during a regular visit to this clinic<br>3 Wait and ask during a regular out-of-facility HIV event<br>5 Contact a healthcare provider via phone<br>4 Other (specify) |
| other_treatment_questions *(required)* | Please specify what else you would do if you had a question about your treatment | |
| contactfacility *(required)* | 105. In the past 12 months, if you had questions regarding your visit or medication collection scheduling or location were you able to contact the facility or DSD model service provider by phone? | 1 Yes<br>0 No |
| unabletocontact *(required)* | 106. What was the reason? | 0 I did not have airtime or data<br>1 I did not have facility contact details<br>2 I did not have the DSD service provider contact details<br>3 The facility contact details were not working<br>4 The DSD service provider contact details were not functional<br>5 The phone call to the facility was not answered<br>6 The phone call to the DSD service provider was not answered<br>7 Other (specify) |
| specifyunabletocontact *(required)* | Please specify | |
| receiveinfo *(required)* | 107. Did you receive adequate information regarding your clinic visits or medication collection scheduling or location? | 0 No<br>1 Yes<br>2 Partly |
| specifynoinfo *(required)* | Specify | |
| V. Patient survey > Questions for patients in DSD models > Questions for likert_dsd2 | | |
| dsd2_likert | 108. Please state how much you agree or disagree with the following statements | 1 Strongly disagree<br>2 Mildly disagree<br>3 Neither agree or nor disagree<br>4 Mildly agree<br>5 Strongly agree<br>6 No response/don't know |
| transport_concerns *(required)* | a. I am concerned about transport cost to get to the clinic | 1 Strongly disagree<br>2 Mildly disagree<br>3 Neither agree or nor disagree<br>4 Mildly agree<br>5 Strongly agree<br>6 No response/don't know |

| Field | Question | Answer |
|---|---|---|
| transport_concerns_dsd *(required)* | b. I am concerned about transport cost to get to my model events (e.g. club meetings, medication pickups) | 1 Strongly disagree<br>2 Mildly disagree<br>3 Neither agree or nor disagree<br>4 Mildly agree<br>5 Strongly agree<br>6 No response/don't know |
| unsure_of_dsd *(required)* | c. I am unsure about how the DSD model works | 1 Strongly disagree<br>2 Mildly disagree<br>3 Neither agree or nor disagree<br>4 Mildly agree<br>5 Strongly agree<br>6 No response/don't know |
| hiv_info_insufficient *(required)* | d. I don't receive enough information about HIV and ART | 1 Strongly disagree<br>2 Mildly disagree<br>3 Neither agree or nor disagree<br>4 Mildly agree<br>5 Strongly agree<br>6 No response/don't know |
| long_waiting_time *(required)* | e. I have to wait for a long time to receive care | 1 Strongly disagree<br>2 Mildly disagree<br>3 Neither agree or nor disagree<br>4 Mildly agree<br>5 Strongly agree<br>6 No response/don't know |
| safe_arv_storing *(required)* | f. I am concerned about safely carrying and storing ARVs at home | 1 Strongly disagree<br>2 Mildly disagree<br>3 Neither agree or nor disagree<br>4 Mildly agree<br>5 Strongly agree<br>6 No response/don't know |
| status_disclosure_concerns *(required)* | g. I am concerned about other people finding out that I am HIV positive | 1 Strongly disagree<br>2 Mildly disagree<br>3 Neither agree or nor disagree<br>4 Mildly agree<br>5 Strongly agree<br>6 No response/don't know |
| other_patient_interactions *(required)* | h. I am concerned about having to interact with other patients in my treatment model | 1 Strongly disagree<br>2 Mildly disagree<br>3 Neither agree or nor disagree<br>4 Mildly agree<br>5 Strongly agree<br>6 No response/don't know |
| model_quality_care *(required)* | i. I am concerned about the quality of care in my treatment model | 1 Strongly disagree<br>2 Mildly disagree<br>3 Neither agree or nor disagree<br>4 Mildly agree<br>5 Strongly agree<br>6 No response/don't know |
| no_concerns *(required)* | j. I have no concerns about my HIV treatment or model of service delivery | 1 Strongly disagree<br>2 Mildly disagree<br>3 Neither agree or nor disagree<br>4 Mildly agree<br>5 Strongly agree<br>6 No response/don't know |
| work_troubles *(required)* | k. I have trouble taking time off work to get to clinic visits | 1 Strongly disagree<br>2 Mildly disagree<br>3 Neither agree or nor disagree<br>4 Mildly agree<br>5 Strongly agree<br>6 No response/don't know |

| Field | Question | Answer |
|---|---|---|
| work_troubles_dsd *(required)* | l. I have trouble taking time off work to get to model events | 1 Strongly disagree<br>2 Mildly disagree<br>3 Neither agree or nor disagree<br>4 Mildly agree<br>5 Strongly agree<br>6 No response/don't know |
| prefer_few_visits *(required)* | m. I prefer to come to the clinic as few times per year as possible | 1 Strongly disagree<br>2 Mildly disagree<br>3 Neither agree or nor disagree<br>4 Mildly agree<br>5 Strongly agree<br>6 No response/don't know |
| patient_connections *(required)* | n. I like being able to connect with other HIV-positive patients at the clinic | 1 Strongly disagree<br>2 Mildly disagree<br>3 Neither agree or nor disagree<br>4 Mildly agree<br>5 Strongly agree<br>6 No response/don't know |
| patient_connections_dsd *(required)* | o. I like being able to connect with other HIV-positive patients in my model of care | 1 Strongly disagree<br>2 Mildly disagree<br>3 Neither agree or nor disagree<br>4 Mildly agree<br>5 Strongly agree<br>6 No response/don't know |
| otherconcerns *(required)* | p. Do you have other concerns about your HIV treatment or model of treatment delivery | 1 Yes<br>0 No |
| specifyconcern *(required)* | Please specify | |
| V. Patient survey > Questions for patients in DSD models > Recommendation subgroup | | |
| dsd_recommendation *(required)* | 109. Would you recommend this model to another patient who is on ART treatment? | 1 Yes<br>0 No |
| explain_recommendation *(required)* | 110. Why or why not? | |
| disappointed_dsd *(required)* | 111. Were you disappointed with any aspect of your care in this DSD model? | |
| dsd_improvement *(required)* | 112. How could services on this model be improved?<br>*(select all that apply)* | 1 More staff<br>2 More information provided by staff<br>3 Better, more polite friendlier, nurse and counselor attitude<br>4 Better, more polite, or friendlier, reception and admin staff attitude<br>5 Better location for model events<br>6 Events on different days<br>7 Events at different times of day<br>8 Events outside of work hours<br>9 Shorter waiting time<br>10 More counselling when there are problems<br>11 More counselling overall<br>12 Less counselling<br>13 Being able to pick up ARVs at different and more convenient sites<br>14 Being able to pick up ARVs at more convenient times<br>15 Being able to have someone else pick up your ARVs<br>16 Having somebody to support you take your ARVs |

| Field | Question | Answer |
|---|---|---|
| | | 17 Contacting you when you miss an appointment<br>18 Reminders via phone or SMS<br>19 More months of ARVs given at each visit<br>20 Fewer months of ARVs given at each visit<br>21 Better access to a nurse or clinic staff<br>22 Treatment/support for other illnesses (specify)<br>23 Other (specify and elaborate ) |
| other_dsd_improvement *(required)* | Specify other ways services on this model could be improved | |
| V. Patient survey > Questions for patients in DSD models > Conclusion subgroup | | |
| best_dsd_thing *(required)* | 113. In conclusion, what is the best thing for you about this model? | |
| worst_dsd_thing *(required)* | 114. And what is the worst thing for you about model? | |
| closing | Please thank the participant for their time and ask if they have any additional questions about the study. | |
| notes *(required)* | Surveyor notes | |
| sid_2 *(required)* | Survey ID options | 1 Barcode<br>2 Enter manually |
| barcode_scan_2 *(required)* | Scan survey ID | |
| survey_id_repeat *(required)* | SURVEY ID | |
| not_eligible | This patient is NOT ELIGIBLE for the study. Thank the participant for their time but do not proceed with the survey | |
| surveyor_initials *(required)* | Surveyor initials | |
