## Supplementary file 3 for "The SENTINEL study of differentiated service delivery models for HIV treatment in Malawi, South Africa, and Zambia: research protocol for a prospective cohort study"

### RESEARCH INFORMATION FORM FOR PATIENT SURVEY

**Title of Project: Outcomes of Differentiated Models of Service Delivery for HIV Treatment at Sentinel Sites in South Africa (Sentinel-South Africa)**

**Principal Investigators: Sydney Rosen and Sophie Pascoe**

---

| | |
|---|---|
| Study Number: | |
| Study Title: | **Outcomes of Differentiated Models of Service Delivery for HIV Treatment at Sentinel Sites in South Africa (Sentinel-South Africa)** |
| Sponsor: | Bill & Melinda Gates Foundation |
| Investigators: | Prof Sydney Rosen, Research Professor (Boston University, United States) |
| | Dr. Sophie Pascoe, Principal Researcher ($HE^2RO$, South Africa) |

Good day. My name is _______________, and I am a research assistant at the Health Economics and Epidemiology Research Office ($HE^2RO$), part of Wits University in Johannesburg. I would like to invite you to consider participating in a research study entitled "Outcomes of Differentiated Models of Service Delivery for HIV Treatment at Sentinel Sites in South Africa (Sentinel-South Africa).

**What is this research study about?**

$HE^2RO$ and Boston University in the United States are conducting a research study about different ways that clinics are delivering treatment for HIV in South Africa and how these different ways of delivering treatment, which are called "models of care", affect both the clinics and their staff and the patients who are receiving treatment.

From patients like you, we would like to find out whether the model of care you are participating in has changed your access to treatment, the costs you incur for seeking treatment, and your satisfaction with the services you are receiving. We are inviting adult patients in this clinic who are receiving HIV treatment (antiretroviral therapy, or ART) in any of the models of care offered by this clinic to participate in the study. If you choose to participate, you will be asked to answer a set of questions today, and we will also look at your clinic records.

This study is being conducted by Professor Sydney Rosen from Boston University in the U.S. and Dr. Sophie Pascoe from $HE^2RO$ (Wits University) in South Africa. Other investigators involved in the study are Dr. Amy Huber and Ms. Idah Mokhele from Wits University. This study is being sponsored by the Bill & Melinda Gates Foundation in the United States.

I would like to provide you with some more information about the study, so that you can decide whether you would like to take part in it. Taking part in this study is voluntary. You can choose not to take part, and if you join, you may quit at any time. If you decide not to participate, you will receive the same care from this clinic that you would have received otherwise. This information sheet may have some words that you do not understand. Please ask the study staff to explain any words or information that you do not understand.

**What is the purpose of this research study?**

In many countries, including South Africa, the Department of Health and healthcare providers are trying to find easier ways to provide ART to the many thousands of patients who need it. This effort has led to the development of what are called “differentiated service delivery” models (DSD models), which adjust the timing, location, and other aspects of ART for different kinds of patients. The goal of DSD models is to make treatment more accessible to patients so that they can more easily adhere to treatment and be more satisfied with the services they received and to reduce the burden of the national ART program on clinics and healthcare workers.

In South Africa, the Department of Health has rolled out three main models of care, known as facility pick up points, external pick up point and adherence clubs. Some individual clinics and partner organizations have created other models as well. Now that clinics and patients have some experience with these different models, it is important to find out how many patients are participating in each model, how well the models are performing, how much they cost both the clinics and patients, and whether patients and healthcare providers are satisfied with them. This study is being conducted in collaboration with the Department of Health to try to find answers to these questions, so that models can be improved in the future.

**What happens in this research study?**

This study is taking place in 24 clinics in South Africa. You will be one of approximately 1200 patients to be asked to participate in this study. Patients in this study must be at least 18 years old and receiving ART in any of the models of care offered by your clinic, including conventional care. If you participate in the study, it will take about 60 minutes of your time today.

If you agree to participate in this study, I will ask you a set of question about you, your household, your health, and your experience of being treated at this clinic and the model of care you are participating in, and your satisfaction with the services at this clinic. The questions will take about 30-45 minutes to complete and will be conducted in English, Zulu or Sesotho.

We will also link your answers to the questions today to your clinic medical records. The reason that we need to do this is so we can see whether patients with different characteristics and experiences and in different models of care have different outcomes in their medical records. To make this link, we will ask you your name and information needed to find your individual medical record. After we link your clinic records to the question responses, all links between your answers and your clinic records will be made anonymous—they will not include your name or any other identifiers. We would like to ask your permission to link the answers you give today with your clinic medical records starting when you initiated ART and continuing for the next twenty-four (24) months, including if you transfer to another clinic.

Any reports that are written using information from this study will combine information for many patients and it will not be possible to identify any individual patient from the information that is presented. If you do not wish to give us permission to link your answers with your clinic records, unfortunately you will not be able to participate in this study, but this will in no way influence the health care, treatment or services that you receive at this clinic.

**Are there risks or discomforts from participating?**

This study will not affect the care and treatment you will receive. The main risk to participating in this study is the small risk of loss of confidentiality, i.e., that your name and questionnaire results may become known. The study team will take every measure to ensure that this does not happen, however, and to keep your answers confidential and will not tell or show anyone what you have said. Your name and other identifying information will be used only to locate your clinic records, and not for any other purpose.

The other risk of this study is that some of the questions you are asked may make you feel uncomfortable or emotionally distressed. The study staff and clinic staff will make every effort to reduce your distress. You do not have to respond to any question unless you feel comfortable doing so. We can stop at any time if you prefer not to finish the questionnaire. We can also arrange for you to spend time in a private room or space if you feel that you need to time to recover from your distress. Please inform any member of the staff if you feel that you do not want to remain in the clinic to participate in the study or would like to speak individually with someone on the study or clinic staff.

**Are there potential benefits from participating?**

You will receive no direct benefit from taking part in this study. However, the information that you provide may help us to understand ways to improve healthcare services in facilities in South Africa and how to appropriately support patients who are taking long-term treatment.

**What other choices do I have?**

Your alternative is not to participate in this study.

**Are there any costs or payments to me?**

You will not be paid or incur any costs for your participation in the study. ART is free at this clinic.

**How will my information be protected?**

The information that we collect from this research project will be kept private. Once you agree to join the study, we will assign you a study number in order to protect your privacy. Your real name will not be used in any report coming from this study. All consent forms, questionnaires, and notes from the study will be stored in a locked unit, and only study staff and designated officials will have access to them. We will not report whether you have participated in the study to any community members or health facilities or health care staff. However, complete confidentiality cannot be guaranteed.

Also, upon signing this consent you give designated officials from the Institutional Review Boards at the University of the Witwatersrand, Boston University, and the Office of Human Subject Protection in the U.S. Department of Health and Human Services consent to look at your study records. They are ensuring that everything happening in this study is ethical. They would only review the study records to ensure that your privacy and integrity is being maintained and protected. A description of this study will be available on http://www.ClinicalTrials.gov. This web site will not include information that can identify you. At most, the web site will include a summary of the results. You can search this web site at any time.

In addition, the final data set containing your responses may be posted in a public data repository as a part of the research publication process.  The posted data set will have all identifiers removed so that it is not possible to identify any individual respondent.

**Participant's rights**

Taking part in this study is voluntary. You have the right to refuse to take part. If you decide to be in the study and then change your mind, you can withdraw from the research. Your participation is completely up to you. If you choose to take part, you have the right to stop at any time without losing any of your rights as a patient here in any way. Your decision will not affect your being able to get health care at this clinic or any other health benefits to which you are entitled.

The researchers may decide to discontinue your participation without your permission because he/she may decide that staying in the study will be bad for you, or the sponsor may stop the study.

For questions about the study please contact:

Dr. Sophie Pascoe, Principal Researcher, Health Economics and Epidemiology Research Office, Johannesburg. Telephone 010 001 7930 or.

This study has been approved by the Human Research Ethics Committee (Medical) of the University of Witwatersrand, Johannesburg. HREC safeguards the rights and dignity of all human subjects who agree to participate in a research study and the integrity of the research being conducted.
If you have any concerns over the way the study is being conducted, please contact the chairperson of the HREC who is Dr. Clement Penny and may be contacted on 011 717 2301, or by email at.

#### RESEARCH CONSENT FORM FOR PATIENT SURVEY

Title of Project: Outcomes of Differentiated Models of Service Delivery for HIV Treatment at Sentinel Sites in South Africa (Sentinel-South Africa)

Principal Investigators: Sydney Rosen and Sophie Pascoe

---

INFORMED CONSENT SIGNATURE PAGE

Signing this consent form indicates that you have read the consent form information sheet (or have had it read to you), that your questions have been answered to your satisfaction, and that you voluntarily agree to participate in this research study. You will receive a copy of this consent form to keep.

| ______________________________ | ______________________________ | ____________ | ______ |
|---|---|---|---|
| Participant (Signature or Thumbprint) | (Printed Name and Surname) | Date | Time |

| ______________________________ | ______________________________ | ____________ | ______ |
|---|---|---|---|
| Person Obtaining Consent (Signature) | (Printed Name and Surname) | Date | Time |

| ______________________________ | ______________________________ | ____________ | ______ |
|---|---|---|---|
| Witness* (Signature) | (Printed Name and Surname) | Date | Time |

*Witness signature required if patient provides mark or thumbprint rather than signature

| **Participant Survey ID Number** |
|---|
