## Supplementary file 4 for "The SENTINEL study of differentiated service delivery models for HIV treatment in Malawi, South Africa, and Zambia: research protocol for a prospective cohort study"

### SENTINEL Provider Survey

#### RESEARCH CONSENT FORM FOR PROVIDER SURVEY

**Title of Project: Outcomes of Differentiated Models of Service Delivery for HIV Treatment at Sentinel Sites in South Africa (Sentinel-South Africa)**

**Principal Investigators: Sophie Pascoe and Amy Huber**

---

| | |
|---|---|
| Study Number: | M210241 |
| Study Title: | **Outcomes of Differentiated Models of Service Delivery for HIV Treatment at Sentinel Sites in South Africa (Sentinel-South Africa)** |
| Sponsor: | Bill & Melinda Gates Foundation |
| Investigators: | Dr. Sophie Pascoe, Principal Researcher (HE$^2$RO, South Africa) |
| | Dr. Amy Huber, Senior Researcher (HE$^2$RO, South Africa) |

Good day. My name is _______________, and I am a research assistant at the Health Economics and Epidemiology Research Office (HE$^2$RO), part of Wits University in Johannesburg. I would like to invite you to consider participating in a research study entitled "Outcomes of Differentiated Models of Service Delivery for HIV Treatment at Sentinel Sites in South Africa (Sentinel-South Africa)."

**What is this research study about?**

HE$^2$RO and Boston University in the United States are conducting a research study about different models of HIV testing and treatment delivery in South Africa and how these models of care affect both the clinics and their staff and the patients who are receiving treatment.

We are asking you to participate in this survey because you work at this clinic as a provider who has regular contact with the patients who access ART here. We would like to discuss the models of ART this clinic offers, your experiences with these models, and how the different models have affected the clinic's procedures and your workload and job satisfaction. We would also like to ask you a few questions about differentiated HIV testing. If you choose to participate, you will be asked to answer a set of questions today.

This study is being conducted by Drs. Sophie Pascoe and Amy Huber from HE$^2$RO (Wits University) in South Africa and they are supported by other investigators from HE$^2$RO and Boston University in the United States. This study is being sponsored by the Bill & Melinda Gates Foundation in the United States.

I would like to provide you with some more information about the study, so that you can decide whether you would like to take part in it. Taking part in this study is voluntary. You can choose not to take part and if you join, you may quit at any time. If you decide not to participate, it will have no effect on your position at the clinic. Please ask the study staff to explain anything you do not understand.

**What is the purpose of this research study?**

In many countries, including South Africa, the Department of Health and healthcare providers are trying to find easier ways to provide ART to the many thousands of patients who need it. This effort has led to the development of what are called "differentiated service delivery" models (DSD models), which adjust the timing, location, and other aspects of ART delivery for different kinds of patients. There has also been an introduction of DSD for HIV testing which adjusts those aspects for HIV testing. The goal of DSD models is to make treatment more accessible to patients so that they can more easily adhere to treatment and be more satisfied with the services they received and to reduce the burden of the national ART program on clinics and healthcare workers.

In South Africa, the Department of Health has rolled out three main differentiated models of care, known as facility pick up points, external pick up point and adherence clubs. Some individual clinics and partner organizations have created other models as well (e.g. such as Pele boxes, teen models, etc.). Now that clinics and patients have some experience with these different models, it is important to find out how many patients are participating in each model, how well the models are performing, how much they cost both the clinics and patients, and whether patients and healthcare providers are satisfied with them. This study is being conducted in collaboration with the Department of Health to try to find answers to these questions, so that models can be improved in the future.

**What happens in this research study?**

In this round of data collection, this study is taking place in 24 clinics in South Africa. You will be one of 240 providers to be asked to participate in this part of the study. If you participate in this survey, it will take about 45-60 minutes of your time today. We will ask you questions about clinic and DSD model procedures for testing and treatment, your professional responsibilities and experiences, changes as a result of DSD model implementation, and your job satisfaction. We will not ask you questions about yourself outside your job, your beliefs or behaviors, or any other personal information. We will not record your name or any other personal identifiers, only your position (job title). Any reports that are written using information from this study will combine information for many providers and it will not be possible to identify any individual participant from the information that is presented.

You may also be asked separately to participate in a time and motion observation. If you do participate in that survey we may also link the data that is collected in this provider survey with your time and motion observations. The reason that we do this is to try and understand how the time it takes to deliver differentiated models of care and interact with patients might impact your experience of implementation and your job satisfaction. To make this link we will use a unique participant ID that can only be accessed by the study team and we will not use any other personal identifiers.

**Are there risks or discomforts from participating?**

The only risk to participating in this study is the small risk of loss of confidentiality, i.e., that your name and questionnaire results may become known to others. The study team will take every measure to ensure that this does not happen, however, and to keep your answers confidential and will not tell or show anyone what you have said. None of your answers will be shared with others at this facility. Your participation and your answers will not affect your work at this health facility in any way. You may refuse to answer any questions or choose to stop the interview at any time. You do not have to respond to any question unless you feel comfortable doing so. We can stop at any time if you prefer not to finish the interview.

The other risk of this study is that some of the questions you are asked may make you feel uncomfortable or emotionally distressed. The study staff will make every effort to reduce your distress. You do not have to respond to any question unless you feel comfortable doing so. We can stop at any time if you prefer not to finish the questionnaire.

### SENTINEL Provider Survey

**Are there potential benefits from participating?**

You will receive no direct benefit from taking part in this study. However, the information that you provide may help us to understand ways to improve healthcare services in facilities in South Africa and how to appropriately support both patients and providers.

**What other choices do I have?**

Your alternative is not to participate in this study.

**Are there any costs or payments to me?**

You will not be paid or incur any costs for your participation in the study.

**How will my information be protected?**

The information that we collect from this research project will be kept private. Once you agree to join the study, we will assign you a study number in order to protect your privacy. We will not record your name or other personal identifiers. All consent forms, questionnaires, and notes from the study will be stored in a locked unit, and only study staff and designated officials will have access to them. We will not report whether you have participated in the study to any others at the clinic, community members, or health facilities or health care staff. However, complete confidentiality cannot be guaranteed.

Also, upon signing this consent you give designated officials from the Institutional Review Boards at the University of the Witwatersrand, Boston University, and the Office of Human Subject Protection in the U.S. Department of Health and Human Services consent to look at your study records. They are ensuring that everything happening in this study is ethical. They would only review the study records to ensure that your privacy and integrity is being maintained and protected. A description of this study will be available on http://www.ClinicalTrials.gov and on the South African Clinical Trials Registry (http://www.sanctr.gov.za/) and Pan-African Clinical Trials Registry (https://pactr.samrc.ac.za/). This web site will not include information that can identify you. At most, the web site will include a summary of the results. You can search these web sites at any time.

In addition, the final data set containing your responses may be posted in a public data repository as a part of the research publication process. The posted data set will not have any personal identifiers so that it is not possible to identify any individual respondent.

**Participant's rights**

Taking part in this study is voluntary. You have the right to refuse to take part. If you decide to be in the study and then change your mind, you can withdraw from the research, however because we do not collect any personal identifiers it would not be possible to remove your data once it has been submitted. Your participation is completely up to you. If you choose to take part, you have the right to stop at any time without losing any of your rights as a healthcare worker here in any way. The researchers may decide to discontinue your participation without your permission because he/she may decide that staying in the study will be bad for you, or the sponsor may stop the study.

For questions about the study please contact:

**Dr. Sophie Pascoe**, Principal Researcher, Health Economics and Epidemiology Research Office, Johannesburg. Telephone 010 001 7930 or

This study has been approved by the Human Research Ethics Committee (Medical) of the University of Witwatersrand, Johannesburg. HREC safeguards the rights and dignity of all human subjects who agree to participate in a research study and the integrity of the research being conducted.
If you have any concerns over the way the study is being conducted, please contact the chairperson of the HREC:

### SENTINEL Provider Survey

#### RESEARCH CONSENT FORM FOR PROVIDER SURVEY

Title of Project: Outcomes of Differentiated Models of Service Delivery for HIV Treatment at Sentinel Sites in South Africa **(Sentinel-South Africa)**

Principal Investigators: Sophie Pascoe and Amy Huber

---

INFORMED CONSENT SIGNATURE PAGE

Signing this consent form indicates that you have read the consent form information sheet (or have had it read to you), that your questions have been answered to your satisfaction, and that you voluntarily agree to participate in this research study. You will receive a copy of this consent form to keep.

| ____________________ | ____________________________ | _____________ | _______ |
|---|---|---|---|
| Participant (Signature) | (Printed Name and Surname) | Date | Time |

| ____________________ | ____________________________ | _____________ | _______ |
|---|---|---|---|
| Surveyor (Signature) | (Printed Name and Surname) | Date | Time |

| ____________________ | ____________________________ | _____________ | _______ |
|---|---|---|---|
| Witness* (Signature) | (Printed Name and Surname) | Date | Time |

*Witness signature required if patient provides mark or thumbprint rather than signature

| **Provider Survey ID Number** |
|---|
