## Supplementary file 5 for "The SENTINEL study of differentiated service delivery models for HIV treatment in Malawi, South Africa, and Zambia: research protocol for a prospective cohort study"

Attachment 2a: Provider survey instrument (updated)

**SENTINEL-South Africa Providers' Survey**

Survey ID

**Introduction**

*Ask the participant for a few minutes of their time. Introduce yourself and the study. Provide information on the study as per the training and give the participant the information sheet for the study. If the provider agrees to participate, allow the participant to sign and date the informed consent and then sign and date the consent form yourself. After the consent form has been signed, leave the information sheet with the participant.*

*If consent has been obtained, continue to Question 1. If consent has not been obtained, thank the provider for their time and end the interaction with the provider.*

Surveyor ID ______________________ Facility Name ______________________

Date (DD/MM/YYYY) ______________________

| Q# | Question | Responses |
|---|---|---|
| **1** | **Respondent description and position at facility** | |
| 1.1 | Did you participate in the previous round of SENTINEL interviews that was conducted during 2021? | (1) Yes<br>(2) No |
| 1.2 | What is your current role at this facility? | (1) Site Operations Manager<br>(2) Doctor/Medical Officer/Clinical Officer<br>(3) Lay Counselor (on site)<br>(4) Professional Nurse<br>(5) Staff Nurse<br>(6) Assistant Nurse<br>(7) Outreach Worker or Community Health Worker (off site)<br>(8) Pharmacist<br>(9) Pharmacy Assistant<br>(10) Admin Clerk<br>(11) Data Capturer<br>(12) Other (specify) |
| 1.3 | Are you employed by the DoH/MoH or a partner? | (1) DoH/MoH<br>(2) Partner |
| 1.4 | What is your employment status? | (1) Full time<br>(2) Full time – rotating<br>(3) Part-time<br>(4) Contract<br>(5) Other (specify) |
| 1.5 | How many years have you worked in your current role/capacity? | Years, months |
| 1.6 | How many years have you worked at this facility? | Years, months |
| 1.7 | In a typical five-day work week, how much of your work time do you spend on HIV-related service delivery? | Number of days, decimal allowed |
| 1.8 | In a typical five-day work week, how much of your work time do you spend providing HIV treatment? | Number of days, decimal allowed |
| 1.9 | What is your age in years? | Years |
| 1.10 | What is your gender? | Male/Female |
| **2** | **Involvement in DSD models** | |
| *Surveyor: Explain to respondent what you mean by "DSD models"* | | |
| 2.1 | In the last year has this clinic started offering or implemented any new DSD models or collection methods? | Yes/No |

| Q# | Question | Responses |
|---|---|---|
| 2.2 | Which models are currently offered at this facility? | List models |
| 2.3 | Do you have any responsibilities for DSD models for treatment, such as helping to organize clubs, provide services in the community, etc.? | Yes/no |
| 2.4 | If yes, which models are you involved in? (Check all that apply) | ☐ Adherence club<br>☐ Youth Club<br>☐ Facility Pick Up Point<br>☐ Ward Based Outreach Teams (WBOTs)<br>☐ External Pick Up Point<br>☐ Pele Box (ATM)<br>☐ Home ART delivery<br>☐ Other (specify)<br>☐ Bicycle model |
| 2.6 | In a typical five-day work week, how much of your work time do you spend supporting DSD models, where "supporting" means anything having to do with DSD models including patient care and administration? | Number of days, decimal allowed |
| **3** | **Effect of DSD models on job responsibilities** | |
| 3.1 | Has anything happened in the last year that has affected your workload? | No<br>Yes (Specify) |
| 3.2 | Has anything happened in the last year that has affected your job satisfaction? | No<br>Yes (Specify) |
| 3.3 | Did having some patients in DSD models "free up" some of your time in the last year? | Yes/no/don't know |
| 3.4 | If yes, what exactly do you do with the extra time? | Open ended |
| 3.5 | Do you think that implementing DSD models for treatment delivery made your job harder or easier in the last year? | Harder/easier/no change/don't know |
| 3.6 | In your view, did clinic procedures, policies, resources, attitudes or other aspects of working here change as a result of having DSD models implemented in the last year? | Yes/no |
| 3.7 | If yes, what are those changes? | Open ended |
| **5** | **Views of DSD models** | |
| 5.1 | Do you think that having DSD models available improves or worsens the care that this facility's ART patients receive? | Improve/worsen/no change/don't know |
| 5.2 | Please explain your answer. | Open ended |
| 5.3 | How confident are you that you understand the requirements for each DSD model? | Likert scale<br>Not confident/Slightly confident/Somewhat confident/Fairly confident/Completely confident |
| *5.4* | Have you ever enrolled a patient into a DSD model before they have been on ART for the amount of time specified in the guidelines (6 mos)? | ☐ Yes<br>☐ No |
| *5.5* | If yes, please think about the most recent time you enrolled a patient into a DSD model after less than 6 months on ART. How was the patient selected into the model? | ☐ Patient requested to be enrolled<br>☐ I recommended the model to them<br>☐ Other (specify) |

| Q# | Question | Responses |
|---|---|---|
| *5.6* | If yes, did you have any concerns about enrolling that patient into a DSD model? | Open ended |
| *5.7* | Have you ever had a patient who has moved from a youth or teen model (like a teen club) into an adult model? If yes, did the patient face any challenges? | ☐ Yes<br>☐ No |
| *5.8* | If yes, could you please explain any challenges that the patient(s) faced? | Open ended |
| *5.9* | How might these challenges be addressed? | Open ended |
| 5.10 | Are you aware of targets set by the Department of Health or others for getting more patients into DSD models? | ☐ Yes<br>☐ No |
| 5.11 | Which models have enrolment targets? | ☐ Adherence club<br>☐ Youth Club<br>☐ Facility Pick Up Point<br>☐ Ward Based Outreach Teams (WBOTs)<br>☐ External Pick Up Point<br>☐ Pele Box (ATM)<br>☐ Home ART delivery<br>☐ Other (specify)<br>☐ Bicycle model |
| 5.12 | Do you feel pressure to enroll patients in DSD models? | ☐ Yes<br>☐ No |
| 5.13 | How much time does it take to enroll a patient into a DSD model at this facility? | Minutes_______ |
| 5.14 | How do you think this clinic's offering of DSD models of ART can be improved? | Open ended |
| **6** | **DSD training** | |
| 6.1 | Were you trained on DSD model implementation in the last year (generally or for specific models)? | ☐ Yes<br>☐ No |
| | If yes, what model/models were covered during the training? | ☐ Adherence club<br>☐ Youth Club<br>☐ Facility Pick Up Point<br>☐ Ward Based Outreach Teams (WBOTs)<br>☐ External Pick Up Point<br>☐ Pele Box (ATM)<br>☐ Home ART delivery<br>☐ Other (specify)<br>Bicycle model |
| 6.2 | If yes, do you feel that the training effectively prepared you for DSD implementation? | ☐ Yes<br>☐ No |
| 6.3 | Please elaborate (on your previous response) | Open ended |
| | Did the training cover how to give patients a choice of which DSD model to enroll in? | ☐ Yes (specify)<br>☐ No<br>☐ Don't know |
| 6.4 | Who conducted the training? | ☐ DoH/MoH<br>☐ Partner<br>☐ Other (Specify) |
| 6.5 | Was the training in person or virtual? | ☐ In person |

| Q# | Question | Responses |
|---|---|---|
| | | ☐ Virtual |
| 6.6 | How long was the training? | Number of days ____<br>Number of hours ____ |
| **7** | **DSD Choice** | |
| 7.4 | Are there multiple DSD models that are offered at this facility to patients established on treatment? | ☐ Yes<br>☐ No |
| 7.1 | What information do you give to patients about DSD models offered at the facility? (select all that apply) | ☐ Eligibility criteria<br>☐ Location of medication collection<br>☐ Type of provider seen<br>☐ Frequency of visits<br>☐ Potential benefits of the models<br>☐ Potential drawbacks of the models<br>☐ Other (specify) |
| 7.2 | To which patients do you give information about DSD models? (Select all that apply) | ☐ Eligible patients<br>☐ Patients who are not yet eligible<br>☐ All patients regardless of eligibility<br>☐ Other (specify) |
| 7.3 | Do you give established (stable) patients a choice of whether they join a model or remain in the standard of care? | ☐ Yes<br>☐ No<br>☐ Sometimes (specify) |
| 7.5 | If yes to 7.4, how do eligible patients get to know of all DSD models offered at this facility? | Open ended |
| 7.6 | If yes to 7.4, do you give established patients a choice between available models? | ☐ Give them a choice<br>☐ Do not give them a choice<br>☐ Other (specify) |
| 7.7 | If you do not let the patient choose, what criteria do you use to decide which model would be best for the patient? | Open ended |
| 7.8 | For models for patients who are not eligible for CCMDD, either because they are new on ART or don't have a suppressed viral load, are patients given a choice as to whether to join the model or remain in standard of care? | ☐ Yes<br>☐ No<br>☐ Sometimes (specify) |
| 7.9 | What could be done to better facilitate patient choice for DSD models from the provider perspective? | |
| **8** | **DSD of HIV testing questions** | |
| 8.1 | Which models of DSD testing are currently offered at this facility? | ☐ Facility HIV testing<br>☐ HIV self-testing<br>☐ Community/Outreach HIV testing<br>☐ Other (specify) |
| 8.2 | Do you have any responsibilities for DSD models for testing? | Yes/no |
| 8.3 | If yes, which testing models are you involved in? (Tick all that apply) | ☐ Facility HIV testing<br>☐ HIV self-testing<br>☐ Community/Outreach HIV testing<br>☐ Other (specify) |
| 8.4 | From your perspective, are any patient needs not being met with regard to the models of testing services available at this facility? If yes, please describe these unmet needs. | Open ended |
| 8.5 | What services are offered to those who test negative after the test? | ☐ Counselling<br>☐ PREP<br>☐ Condoms |

| Q# | Question | Responses |
|---|---|---|
| | | ☐ Other (specify) |
| 8.6 | What services are offered to those who test positive after the test? | ☐ Counselling<br>☐ ART<br>☐ Condoms<br>☐ CD4 testing<br>☐ Other (specify) |
| **9. Questions about reengagement** | | |
| 9.1 | When you initiate a patient on ART, do you ask if the patient was ever on ART before? | ☐ Yes<br>☐ No |
| 9.2 | Are patients who are re-initiating ART managed any differently from those who are naïve (first-time) initiators? | ☐ Yes<br>☐ No |
| 9.3 | If yes to 9.3, How are these processes different from naïve (first time) initiators? Probe: easier, harder, takes more time, takes less time. | Open ended |
| 9.4 | With regard to patients who are re-engaging, what are the main reasons that patients offer at this facility as to why they disengaged from care? | ☐ Mobility/relocation<br>☐ Distance to the clinic<br>☐ Difficulty getting time off work<br>☐ Change in employment<br>☐ Tired of treatment<br>☐ Other (specify) |
| 9.5 | With regard to patients who are re-engaging, what are the main reasons that patients offer at this facility as to why they chose to come back to the facility to re-engage in care? | ☐ Worried about being off ART<br>☐ Accessing care is now easier<br>☐ They feel sick<br>☐ Successful tracing attempts by the facility<br>☐ Concern for their children<br>☐ Other (specify) |
| 9.63 | What is the process at this facility to re-initiate a patient on ART if they arrive at the facility without a transfer letter? | Open ended |
| 9.7 | Can you think of any modifications to HIV services that could support a quicker or easier re-engagement process? | Open ended |
| **10. Questions about the first 6 months on treatment** | | |
| 10.1 | In the past, patients who are initiating ART have been more likely to drop out of care (become lost to follow up) soon after they start, often within the first 6 months. Have you noticed that this is happening here at your facility? | ☐ Yes<br>☐ No |
| 10.2 | To what extent do you think this is a problem at this facility? Probe: Please tell me why you think this? | Open ended |
| 10.3 | Does the facility offer any special services or interventions during the early treatment period (first 6 months on ART)? Probe: if yes, please explain in detail | Open ended |
| 10.4 | Could you please provide information about how many months of ARVs you dispense and whether counselling and education is provided for each visit in the first six months? | |
| 10.4a | Visit 1 (ART initiation visit): | Amount of ARVs dispensed: ____ doses<br>Counselling session provided? Y/N<br>Education session provided? Y/N |
| 10.4b | Visit 2: | Amount of ARVs dispensed: ____ doses<br>Counselling session provided? Y/N<br>Education session provided? Y/N |

| Q# | Question | Responses |
|---|---|---|
| 10.4c | Visit 3: | Amount of ARVs dispensed: ____ doses<br>Counselling session provided? Y/N<br>Education session provided? Y/N |
| 10.4d | Visit 4 (only if occurs within 6 months): | Amount of ARVs dispensed: ____ doses<br>Counselling session provided? Y/N<br>Education session provided? Y/N |
| 10.4e | Visit 5 (only if occurs within 6 months): | Amount of ARVs dispensed: ____ doses<br>Counselling session provided? Y/N<br>Education session provided? Y/N |
| 10.4f | Visit 6 (only if occurs within 6 months): | Amount of ARVs dispensed: ____ doses<br>Counselling session provided? Y/N<br>Education session provided? Y/N |
| 10.5 | Are there any patient or staff support measures you can think of that would improve missed appointments in the first 6 months on ART? | |
| 11. | **Is there anything else you'd like to tell me before we finish?** | **Open ended** |

*Surveyor: "Thank you sincerely for your time. We have completed this interview and are grateful for your help in improving our understanding of how other models of HIV treatment delivery affects healthcare providers' jobs."*
