## Supplementary file 6 for "The SENTINEL study of differentiated service delivery models for HIV treatment in Malawi, South Africa, and Zambia: research protocol for a prospective cohort study"

### D1. SENTINEL2.0-South Africa Time-Motion Form

| Field | Question | Answer |
|---|---|---|
| Form control | | |
| observer_id *(required)* | Observer ID | |
| specify_observer *(required)* | Specify the observer | |
| district *(required)* | District name | |
| wr_facilities *(required)* | Facility name | |
| mp_facilities *(required)* | Facility name | |
| kzn_facilities *(required)* | Facility name | |
| Observation_day *(required)* | Observer: Please select observation day for this participant | 1 Day 1<br>2 Day 2 |
| sid *(required)* | Participant ID options | 1 Barcode<br>2 Enter manually |
| barcode_scan *(required)* | Scan participant ID | |
| participant_id *(required)* | Participant ID | |
| cadre *(required)* | Participant cadre | 1 Professional Nurse<br>2 Enrolled nurse<br>3 Doctor<br>4 Pharmacist<br>5 Pharmacist Assistant<br>6 Other |
| other_cadre | Specify other cadre | |
| observation_date *(required)* | Date | |
| observation_day *(required)* | Day of week<br>*NB: Please ensure that you select the same day as indicated in the calendar* | 1 Monday<br>2 Tuesday<br>3 Wednesday<br>4 Thursday<br>5 Friday<br>6 Saturday<br>7 Sunday |

| Field | Question | Answer |
|---|---|---|
| start_time *(required)* | Time observation started for day | |
| Participant demographic information | | |
| Participant demographic information > Age and gender | | |
| age *(required)* | 1. What is your age in years? | |
| gender *(required)* | 2. What is your gender? | 1 Male<br>2 Female |
| participated *(required)* | 3. Have you participated in the provider survey? | 1 Yes<br>2 No |
| providersurveyid *(required)* | Observer please fill in the provider Survey ID<br>*if the participant was enrolled in the provider survey (Domain 2), Please provide the survey ID below* | |
| Participant demographic information > roleandexperiences | | |
| earliertimeandmotion *(required)* | 4. Have you participated in earlier rounds of the Time and Motion study? | 1 Yes<br>2 No |
| currentrole *(required)* | 5. What is your current role at this facility? | |
| yearsinrole *(required)* | 6. How many years have you worked in your current role/capacity?<br>*Enter years* | |
| monthsinrole *(required)* | Observer: Now enter the number of months | |
| yearsatfacility *(required)* | 7. How many years have you worked at this facility? | |
| monthsatfacility *(required)* | Observer: Now enter the number of months | |
| Participant demographic information > summaryofworkdays | | |
| hivservicestime *(required)* | 8. In a typical five-day work week, how much of your work time do you spend on HIV-related service delivery?<br>*(Number of days, decimal allowed). Enter a zero if no time is spent on HIV-related service delivery* | |
| hivservicestreatment *(required)* | 9. In a typical five-day work week, how much of your work time do you spend providing HIV treatment?<br>*(Number of days, decimal allowed). Enter a zero if no time is spent on providing HIV treatment* | |
| Participant demographic information > facilityprocedures | | |
| target *(required)* | 10. Is there a daily/weekly/monthly target for the number of patients each nurse/counsellor/others should see in a day/week/month? | 1 Yes<br>2 No |
| target_by *(required)* | Observer please fill in the target type (duration)<br>*Please indicate if target is per day, week, or month* | |
| target_number *(required)* | Observer please fill in the target<br>*Please indicate the target (e.g number of patient)* | |
| practice_normalpatients *(required)* | 11. If there are fewer than normal patients at the facility what is normal practice for clinic staff? | 1 a) Staff assigned to other non-patient activities/duties (normal work hours apply)<br>2 b) Staff have more unallocated time but normal work hours apply<br>3 c) Staff can leave once all their patients have been seen and paperwork complete<br>4 d) Other specify |
| otherpractice_normalpatients *(required)* | Observer please specify other | |
| practice_fewpatients *(required)* | 12. If there are more patients than normally expected what is the normal practice? | 1 a) The clinic remains open until all patients are seen (clinic hours extended)<br>2 b) Some patients may be asked to return another day unless urgent (normal clinic hours apply)<br>3 c) Other specify |
| otherpractice_fewpatients *(required)* | Observer please specify other | |
| totpatientsseen *(required)* | Total number of unique patients seen by provider today (day of observation)<br>*Please look for this number in the register at the end of the day. Or manually count patients seen by the provider at the end of the observation day* | |
| source *(required)* | Source<br>*Please specify where you verified the number* | |
| intro | Introduction<br>*Confirm that the provider has consented to being observed and ask if there are any remaining questions or concerns before you start.* | |
| Observation of time block (1) | | (Repeated group) |
| start *(required)* | Time observation started | |
| type *(required)* | Patient time block or non-patient time block | 1 a. patient interaction<br>2 b. non-patient interaction |
| patient *(required)* | Patient's primary category | 1 1. Non-HIV (chronic or acute)<br>2 2. HIV not yet on ART |

| Field | Question | Answer |
|---|---|---|
| | | 3 3. Initiating ART today<br>4 4. First six months on ART<br>5 5. Established on ART > 6 months<br>6 6. Elevated Viral Load<br>7 7. Patient representative or buddy (not patient)<br>8 8. Other (specify) |
| other_patient_category | Specify other patient category | |
| visit *(required)* | Primary reason for visit | 1 a. Acute emergency or COVID related<br>2 b. Scheduled clinical consultation<br>3 c. Unscheduled clinical consultation<br>4 d. Medication refill or collection only<br>5 e. HIV testing<br>6 f. Other (specify) |
| other_visit | Specify other reason for visit | |
| visit_procedure *(required)* | Procedures conducted during visit | 1 1. Medication refill or collection<br>2 2. ART initiation<br>3 3. Counseling<br>4 4. Laboratory test or health screening<br>5 5. Re-scripting<br>6 6. Other (specify) |
| other_visit_procedure *(required)* | Specify other procedure conducted during visit | |
| model *(required)* | Model patient is enrolled in | 1 1. Standard care (not enrolled in DSD models)<br>2 2. Adherence club<br>3 3. Facility pick up point<br>4 4. External pickup point<br>5 5. Youth club<br>6 6. Pele Box or locker<br>7 7. Home ART delivery<br>8 8. Bicycle model<br>9 9. Not applicable (non- ART)<br>10 10. DSD model unknown<br>11 11. Unknown whether in DSD model<br>12 12. Other (specify) |
| other_model | Specify other model patient is enrolled in | |
| block *(required)* | Reason for non-patient time block | 1 a. Patient related task (completing record, referral, etc.)<br>2 b. DSD model related task (Specify model and activity e.g. Home delivery-preparing medications for delivery, CAG-leading a group)<br>3 c. General administration or meetings<br>4 d. External outreach (community activities)<br>5 e. Personal break (lunch, tea, other personal time)<br>6 f. Training<br>7 g. Free time/ no patients<br>9 h. Transit<br>8 i. Other (specify) |
| other_block | Specify main reason for other block | |

| Field | Question | Answer |
|---|---|---|
| specify_task *(required)* | Specify the DSD model and related task | |
| alreadyseen *(required)* | Was this patient already seen by the provider? | 1 Yes<br>2 No |
| observercomments *(required)* | Observer's comments (if any)<br>*Your comments for this time block* | |
| provider_comments *(required)* | Provider's comments (if any) | |
| observercomments2 *(required)* | Observer's comments (if any)<br>*Your comments for this time block* | |
| end *(required)* | Time observation ended | |
| observer_notes *(required)* | Observer's notes<br>*Your overall comments for the entire day's observation* | |
| end_time *(required)* | Time observations ended for day | |
| closing | Please thank the participant for their time and ask if they have any additional questions about the study. | |
| sid_2 *(required)* | Participant ID options | 1 Barcode<br>2 Enter manually |
| barcode_scan_2 *(required)* | Scan participant ID | |
| participant_id_repeat *(required)* | PARTICIPANT ID | |
