## Supplementary file 7 for "The SENTINEL study of differentiated service delivery models for HIV treatment in Malawi, South Africa, and Zambia: research protocol for a prospective cohort study"

### D5a. SENTINEL2.0-South Africa Patient Testing Component

| Field | Question | Answer |
|---|---|---|
| Screening form | | |
| Screening form > Form control | | |
| screening_no *(required)* | Screening number<br>*screeningno* | |
| surveyor *(required)* | Surveyor ID | |
| specifysurveyor *(required)* | Specify the surveyor | |
| district *(required)* | District name | |
| wr_facilities *(required)* | Facility name | |
| mp_facilities *(required)* | Facility name | |
| kzn_facilities *(required)* | Facility name | |
| screeningdate *(required)* | Date | |
| intro | I. Introduction<br>*Ask the patient for a few minutes of their time. Introduce yourself and the study. Provide information on the study as per the training and give the patient the information sheet for the study. If the patient expresses interest in being in the study, proceed to section II below and then complete the eligibility screening in section III. If the participant is not interested in being in the study, thank them for their time, end the interaction, and answer the questions in section II below.* | |
| agreement | II. Brief screening agreement<br>*You are being asked to voluntarily give us some information to see if you might qualify to be enrolled in a research study. We are asking you to be in this study because you are testing for HIV today, and we are doing research on the different models of service delivery for HIV testing. If you agree, we will ask you to tell us your age, gender, and whether you plan to or have already tested for HIV at this clinic. This information will tell us if you are qualified to enroll in the study. If you qualify for the study and decide to join, the information that we get from you will become part of your study record. If you do not qualify or decide not to join the study, the information that we get from you will only be used to keep track of how many patients we have invited into the study and if those who do qualify and decide to join are different in age, gender, or receipt of HIV testing services, from those who do not join. We will keep this information until we have finished enrollment for the study, then destroy it. Saying yes to this screening does not mean you have to agree to be in the study. If you have any questions, please ask them now or at any time you can contact Sophie Pascoe, Principal Researcher, Health Economics and Epidemiology Research Office, Johannesburg, telephone 010 001 7930 or.* | |
| Screening form > III. Demographic description | | |
| gender *(required)* | 1. Gender | 0 Male<br>1 Female<br>2 Other |
| age *(required)* | 2. How old were you at your last birthday?<br>*Age (years)* | |

| Field | Question | Answer |
|---|---|---|
| Screening form > IV. Eligibility screening | | |
| Eligibilitynote | If both answers to the following screening questions 3 and 4 are "YES", proceed to administer the informed consent process. If any answers are "NO", thank the patient for their time and end the interaction. | |
| planningtotest *(required)* | 3. Are you planning to test for HIV at this facility today or have you already tested for HIV at this facility today?<br>*STOP if the response is NO!* | 1 Yes<br>0 No |
| ispatient18 *(required)* | 4. Are you currently age 18 or older?<br>*STOP if the response is NO!* | 1 Yes<br>0 No |
| Screening form > IV. Eligibility decision | | |
| eligible | This patient is ELIGIBLE for the study. Please proceed with the informed consent process | |
| consenting *(required)* | This patient | 1 CONSENTED to participate (Fill in survey instrument ID number below and proceed with survey)<br>2 DID NOT CONSENT to participate (Thank the participant for their time and end the interaction)<br>3 N/A (Target for this model has been reached) |
| V. Patient survey | | |
| sid *(required)* | Survey ID options<br>*For patients eligible for and consenting to the study, please indicate study survey ID number here:* | 1 Barcode<br>2 Enter manually |
| barcode_scan *(required)* | Scan survey ID | |
| survey_id *(required)* | SURVEY ID | |
| facilityloc *(required)* | Location within facility | |
| structurenote | For patients who are recruited before their HIV test, Section 1 questions will be asked before their HIV test and Section 2 questions will be asked after their HIV test. For those who are recruited after their HIV test, the questionnaire will start with Section 1 and proceed directly to Section 2.<br>*Note on survey structure:* | |
| intro_statement | "Thank you for agreeing to participate in this survey. My name is ______. I will be asking you the questions. Most of the questions require that you select one of the options as your answer, although some questions you can select all the answers that apply. I will specify the options and instructions for you as I ask each question. If your answer is not one of the specified options please tell me and I will write your answer down. Please feel free to tell me whatever you are comfortable sharing. You should also remember that you do not have to share anything that you are not comfortable sharing and that you can stop this interview at any time without any risk to your rights or treatment and care. There are no right or wrong answers, so please be honest and help us to understand what is true for you and your community. Are you ready to begin?<br>*Read the following statement. Please repeat the statement translated into the local language based on primary languages.* | |
| respondenttested *(required)* | Has this respondent tested? | 1 a. Not yet tested (will complete section 1 now and section 2 after completing test)<br>2 b. Not yet tested (will complete section 1 now and follow up by telephone for section 2)<br>3 c. Tested and has results (will complete section 1 and section 2 now) |
| section1note | SURVEYOR NOW YOU ARE ABOUT TO START SECTION 1 | |
| V. Patient survey > Section 1: Pre-test questions for those recruited before their HIV test, or starting point for those recruited after their HIV test. | | |
| note | Surveyor: "I'm going to start by asking you some basic questions about who you are, where you live, and your education and employment." | |
| V. Patient survey > Section 1: Pre-test questions for those recruited before their HIV test, or starting point for those recruited after their HIV test. > Respondent demographics and socio-economic status | | |
| nationality *(required)* | 1. What is your nationality/country of origin? | 1 South Africa<br>2 Botswana<br>3 Lesotho<br>4 Mozambique<br>5 Malawi<br>6 Namibia<br>7 Swaziland<br>8 Zambia<br>9 Zimbabwe<br>10 Tanzania |

| Field | Question | Answer |
|---|---|---|
| | | 11 Burundi<br>12 Other African country (specify)<br>13 Other (specify) |
| othernationality *(required)* | Please specify nationality/country of origin | |
| maritalstatus *(required)* | 2. What is your marital status? | 1 Never married<br>2 Married (customary/traditional or legal/civil)<br>3 Divorced<br>4 Separated<br>5 Widowed |
| partner *(required)* | 3. Is there someone who you have a relationship with and who you call your partner? | 1 Yes<br>0 No |
| livingwithspouse *(required)* | 4. Do you currently live with your husband/wife or your partner? | 1 No<br>2 Yes, married or living together |
| V. Patient survey > Section 1: Pre-test questions for those recruited before their HIV test, or starting point for those recruited after their HIV test. > Respondent demographics and socio-economic status > Demographic subgroup | | |
| currenthousehold *(required)* | 5. Do you think of the house you currently live in as your main house? | 1 Yes<br>2 No, my main house is somewhere else in South Africa<br>3 No, my main house is in another country |
| reading *(required)* | 6. Do you know how to read and write? | 1 No<br>2 Yes – read and write<br>3 Yes – read only |
| edulevel *(required)* | 7. What was the highest level of school that you completed? | 1 No schooling<br>2 Primary<br>3 Secondary<br>4 Certificate/Diploma/ Post-secondary<br>5 Graduate degree |
| occupation *(required)* | 8. What is your occupation? | 1 Farming (my own or my family's farm)<br>2 Farm worker (someone else's farm)<br>3 Domestic worker or carer (paid)<br>4 Informal sector job (not farming or domestic) (e.g. trader, day service provider)<br>5 Formal sector job (salaried)<br>6 Household work and/or childcare (my own house, not paid)<br>7 Unemployed but looking for work<br>8 Student or trainee<br>9 Retired<br>11 Self-employed/own business<br>12 Unemployed but not looking for work<br>10 Other (specify) |
| otheroccupation *(required)* | Please specify your occupation | |
| mostmoney *(required)* | 9. Where do you get MOST of your money from? | 1 Salary, business or job (formal or informal sector)<br>2 Government social grant<br>3 Spouse/partner<br>4 Parents/relatives<br>5 Friends<br>6 Other (specify) |

| Field | Question | Answer |
|---|---|---|
| specifymoney *(required)* | Please specify | |
| V. Patient survey > Section 1: Pre-test questions for those recruited before their HIV test, or starting point for those recruited after their HIV test. > Respondent demographics and socio-economic status > Demographic subgroup2 | | |
| foodsecurity *(required)* | 10. Do you or the people in your household go without food often, sometimes, seldom, never? | 1 Never<br>2 Seldom<br>3 Sometimes<br>4 Often |
| governmentgrant *(required)* | 11. Do you or does anybody in your household, currently receive any support or grant from the government?<br>*Tick all that apply* | 0 No<br>1 Child grant<br>2 Partial disability / illness grant / temporary grant<br>3 Pension grant<br>4 Disability grant<br>5 Unemployment grant/UIF<br>7 COVID social relief grant<br>6 Other (specify) |
| othersupportgrant *(required)* | Please specify the support or grant from the government | |
| healthcare_money *(required)* | 12. If a person in your household became ill and 100 Rands was needed for treatment or medicines, would you say it would be very easy, easy, difficult, or very difficult to find the money? | 1 Very difficult<br>2 Difficult<br>3 Easy<br>4 Very easy |
| V. Patient survey > Section 1: Pre-test questions for those recruited before their HIV test, or starting point for those recruited after their HIV test. > Healthcare access and cost | | |
| visitreason *(required)* | 13. What is the main reason for you coming to this facility today? | 1 HIV test<br>2 Pregnancy/antenatal<br>3 Chronic condition (specify)<br>4 Acute care (specify)<br>5 Other (specify) |
| specifyvisitreason *(required)* | Please specify | |
| additionalservices *(required)* | 14. Which health care services are you routinely receiving at this facility?<br>*(Tick all that apply)* | 0 None<br>1 TB treatment<br>2 Diabetes<br>3 Hypertension<br>4 Asthma<br>5 Mental health<br>6 Malaria<br>7 Family planning<br>8 Antenatal care<br>9 Child health care<br>10 TB preventative therapy (TPT)<br>11 Other (specify) |
| specifyservices *(required)* | Please specify | |
| transporttoclinic *(required)* | 15. How do you usually get to the clinic?<br>*Tick all that apply* | 1 Walk<br>2 Mini-bus/common taxi<br>3 Own car<br>4 Meter taxi/Uber/Taxify/Hired taxi<br>5 Brought by family/friends in their vehicles<br>6 Other (specify) |
| othertransport *(required)* | Please specify other means of getting to the clinic | |
| V. Patient survey > Section 1: Pre-test questions for those recruited before their HIV test, or starting point for those recruited after their HIV test. > Healthcare access and cost > Travel time subgroup | | |
| traveltime *(required)* | 16. How long does it take you to get to the clinic? (One way – from home to the clinic)<br>*Enter hours* | |
| travelminutes *(required)* | Surveyor: Now enter minutes<br>*Minutes* | |
| otherwisedoing *(required)* | 17. What would you otherwise have been doing if you had not come to the clinic today? | 1 Housework<br>2 Childcare (own children)<br>3 Caring for a relative or friend<br>4 Voluntary work<br>5 Leisure activities<br>6 Attending school or university |

| Field | Question | Answer |
|---|---|---|
| | | 7 On sick leave<br>8 Seeking work<br>9 Paid work<br>10 Other (specify) |
| specifyotherwisedoing *(required)* | Please specify other expenses/costs you incur for each clinic visit | |
| expenses *(required)* | 18. What expenses/costs do you incur for each clinic visit? | 0 No costs<br>1 Transport<br>2 Loss of income due to missing work<br>3 Child care<br>4 Food/drinks<br>5 Other (specify) |
| otherexpenses *(required)* | Please specify other expenses/costs you incur for each clinic visit | |
| V. Patient survey > Section 1: Pre-test questions for those recruited before their HIV test, or starting point for those recruited after their HIV test. > Questions related to HIV test experience | | |
| whytest *(required)* | 19. Why are you testing for HIV today? | 1 Pregnancy<br>2 Feeling ill<br>3 Partner or former partner was diagnosed with HIV<br>4 Workplace requirement<br>5 PrEP<br>6 Just checking my status/voluntary testing<br>7 Recommended by healthcare provider<br>8 Other (specify) |
| specifywhytest *(required)* | Please specify | |
| evertested *(required)* | 20. Have you ever taken an HIV test before?<br>*(If no, skip to post-test questions)* | 0 No<br>1 Yes<br>2 Don't know/can't remember |
| howmanytimes *(required)* | How many times have you tested for HIV? | |
| V. Patient survey > Section 1: Pre-test questions for those recruited before their HIV test, or starting point for those recruited after their HIV test. > Questions related to HIV test experience > Follow up questions (ever tested before) (1) | | (Repeated group) |
| recalltestdate *(required)* | Can you recall the test date? | 0 No<br>1 Yes, month and year<br>2 Yes, day month and year |
| testdatemy *(required)* | Test date (MM/YYYY) | |
| testdatedmy *(required)* | Test date (DD/MM/YYYY) | |
| testresult *(required)* | HIV test result | 0 Negative<br>1 Positive<br>2 Indeterminate |
| location *(required)* | Location of HIV test | 1 Public clinic or hospital<br>2 Private provider<br>3 Mobile HIV testing vehicle<br>4 At home<br>5 Workplace testing<br>6 Other (specify) |
| specifylocation *(required)* | Please specify | |
| reasonfortest *(required)* | Reason(s) for testing | 1 Pregnancy<br>2 Feeling ill<br>3 Partner or former partner was diagnosed with HIV<br>4 Workplace requirement<br>5 PrEP<br>6 Just checking my status/voluntary testing<br>7 Recommended by healthcare provider<br>8 Other (specify) |
| specifyreason *(required)* | Please specify | |
| V. Patient survey > Section 1: Pre-test questions for those recruited before their HIV test, or starting point for those recruited after their HIV test. > Questions related to HIV test experience > Follow up questions (ever tested before) (2) | | (Repeated group) |

| Field | Question | Answer |
|---|---|---|
| recalltestdate *(required)* | Can you recall the test date? | 0 No<br>1 Yes, month and year<br>2 Yes, day month and year |
| testdatemy *(required)* | Test date (MM/YYYY) | |
| testdatedmy *(required)* | Test date (DD/MM/YYYY) | |
| testresult *(required)* | HIV test result | 0 Negative<br>1 Positive<br>2 Indeterminate |
| location *(required)* | Location of HIV test | 1 Public clinic or hospital<br>2 Private provider<br>3 Mobile HIV testing vehicle<br>4 At home<br>5 Workplace testing<br>6 Other (specify) |
| specifylocation *(required)* | Please specify | |
| reasonfortest *(required)* | Reason(s) for testing | 1 Pregnancy<br>2 Feeling ill<br>3 Partner or former partner was diagnosed with HIV<br>4 Workplace requirement<br>5 PrEP<br>6 Just checking my status/voluntary testing<br>7 Recommended by healthcare provider<br>8 Other (specify) |
| specifyreason *(required)* | Please specify | |
| thankparticipant | Surveyor: Please thank the participant for their time so far and ask if they have any additional questions about the study. Explain that there is a second portion of the survey to be conducted after their HIV test is complete and they can either come to you after the test or you can follow up with them telephonically after one week.<br>*For participants who have not tested yet* | |
| V. Patient survey > Section 2: Post-test questions for those recruited before their HIV test. Questionnaire will continue directly from Section 1 to Section 2 for those recruited after their HIV test. | | |
| section2note | SURVEYOR NOW YOU ARE ABOUT TO START SECTION 2 | |
| sectionnote | Surveyor: “Thank you for returning to complete the survey after your test. As with before, I will be asking you the questions. Please feel free to tell me whatever you are comfortable sharing. You should also remember that you do not have to share anything that you are not comfortable sharing and that you can stop this interview at any time without any risk to your rights or treatment and care. There are no right or wrong answers, so please be honest and help us to understand what is true for you and your community. Are you ready to begin?" | |
| V. Patient survey > Section 2: Post-test questions for those recruited before their HIV test. Questionnaire will continue directly from Section 1 to Section 2 for those recruited after their HIV test. > Questions for all patients after testing for HIV | | |
| todayslocation *(required)* | 1. Location of today’s HIV test | 1 In-facility: VCT clinic<br>2 In-facility: ANC<br>3 In-facility: Other (specify)<br>4 Out-of-facility (specify) |
| specifytestloc *(required)* | Please specify | |
| testwhere *(required)* | 2. If given a choice, where would you have preferred to have today’s HIV test: | 1 Here at the clinic<br>2 At the pharmacy in your community<br>3 At home<br>4 Somewhere else (specify) |
| specifywhere *(required)* | Please specify | |
| referral *(required)* | 3. Were you referred from another department within the clinic before your test? | 1 Yes<br>0 No |
| specify *(required)* | 4. Specify | 1 Acute care<br>2 Chronic care<br>3 ANC<br>4 Other (specify) |
| specifydepartment *(required)* | Specify department | |
| who *(required)* | 5. Who conducted your HIV test? | 1 Nurse<br>2 Counsellor<br>3 Self<br>4 Other (specify) |

| Field | Question | Answer | |
|---|---|---|---|
| specifywho *(required)* | Specify | | |
| preferwho *(required)* | 6. Would you preferred to have someone else conduct your HIV test? | 0 | No |
| | | 1 | Yes, nurse |
| | | 2 | Yes, counsellor |
| | | 3 | Yes, self |
| | | 4 | Yes, other (specify) |
| specifyprefer *(required)* | Specify | | |
| confirmatory *(required)* | 7. Was this test a confirmatory (re)-testing after first indeterminate test? | 1 | Yes |
| | | 0 | No |
| firsttestloc *(required)* | 8. Where was first test conducted? | 1 | In-facility: VCT clinic |
| | | 2 | In-facility: ANC |
| | | 3 | In-facility: Other (specify) |
| | | 4 | Out-of-facility (specify) |
| specifyfirsttestloc *(required)* | Specify | | |
| when *(required)* | 9. When was first test conducted? | | |
| V. Patient survey > Section 2: Post-test questions for those recruited before their HIV test. Questionnaire will continue directly from Section 1 to Section 2 for those recruited after their HIV test. > Questions for all patients after testing for HIV > Satisfaction subgroup | | | |
| satisfaction *(required)* | 10. How satisfied were you with the care you received today during your HIV testing experience | 1 | Very satisfied |
| | | 2 | Satisfied |
| | | 3 | Moderately satisfied |
| | | 4 | Slightly satisfied |
| | | 5 | Not satisfied |
| elaborate *(required)* | 11. Could you please elaborate on the reasons for your level of satisfaction? | | |
| serviceimprovement *(required)* | 12. I would now like to ask a few questions specifically about your experience with the HIV testing component of your experience today. How could services on this HIV testing be improved? *(select all that apply)* | 1 | More staff |
| | | 2 | More information provided by staff |
| | | 3 | More information provided by staff |
| | | 4 | Better, more polite, or friendlier nurse attitude |
| | | 5 | Better, more polite, or friendlier reception and admin staff attitude |
| | | 6 | Better location for testing events |
| | | 7 | Testing offered on different days |
| | | 8 | Testing offered at different times of day |
| | | 9 | Testing offered outside of work hours |
| | | 10 | Shorter waiting time |
| | | 11 | More counselling when there are problems |
| | | 12 | More counselling overall |
| | | 13 | Less counselling |
| | | 14 | Being able to test at different and more convenient sites |
| | | 15 | Having somebody to support you take your HIV test |
| | | 16 | Better access to a nurse or clinic staff |
| | | 17 | Other (specify and elaborate ) |
| specifyservice *(required)* | Specify | | |
| V. Patient survey > Section 2: Post-test questions for those recruited before their HIV test. Questionnaire will continue directly from Section 1 to Section 2 for those recruited after their HIV test. > Questions for all patients after testing for HIV > Experience subgroup | | | |
| helpfulpart *(required)* | 13. In conclusion, what is the most helpful or supportive part for you about your experience testing for HIV today and why? | | |
| worstpart *(required)* | 14. And what is the worst part for you about your experience testing for HIV today and why? | | |
| hivtestresult *(required)* | T1. What was the result of your HIV test today? *HIV test result* | 1 | Positive |
| | | 2 | Negative |
| | | 3 | Indeterminate |

| Field | Question | Answer |
|---|---|---|
| | | 3 Indeterminate<br>4 Not willing to disclose (end survey) |
| V. Patient survey > Section 2: Post-test questions for those recruited before their HIV test. Questionnaire will continue directly from Section 1 to Section 2 for those recruited after their HIV test. > Questions for patients testing positive for HIV | | |
| V. Patient survey > Section 2: Post-test questions for those recruited before their HIV test. Questionnaire will continue directly from Section 1 to Section 2 for those recruited after their HIV test. > Questions for patients testing positive for HIV > Questions for patients with previous ART experience | | |
| previousmed *(required)* | 1. Have you previously taken any medication for treatment of HIV?<br>*(Interviewer be sure to specify this does not include medication for HIV prevention (PrEP or PEP)) (If no, skip to question 12)* | 1 Yes<br>0 No |
| table | Please complete the following table: | |
| V. Patient survey > Section 2: Post-test questions for those recruited before their HIV test. Questionnaire will continue directly from Section 1 to Section 2 for those recruited after their HIV test. > Questions for patients testing positive for HIV > Questions for patients with previous ART experience > Previous ART (1) | | (Repeated group) |
| recalldate *(required)* | Can you recall ART start date? | 0 No<br>1 Yes, month and year<br>2 Yes, day month and year |
| artstartdatemy *(required)* | ART start date (MM/YYYY) | |
| artstartdatedmy *(required)* | ART start date (DD/MM/YYYY) | |
| reasonforstarting *(required)* | Reason for starting/re-starting ART<br>*(see list)* | 1 Pregnancy<br>2 Feeling ill and wanted to feel better<br>3 Worried about my future or children's future<br>4 Health care provider recommended treatment<br>5 Other (specify) |
| otherreason *(required)* | Specify | |
| artenddate *(required)* | ART end date<br>*(DD/MM/YYYY)* | |
| reasonforstopping *(required)* | Reason for stopping medication | 1 Travel made it difficult to obtain medication<br>2 Work made it difficult to obtain medication<br>3 I was worried that someone would find out about my status<br>4 I had nowhere to discreetly store my medication<br>5 I started using alternative medication<br>6 I started feeling better<br>7 I started feeling worse or had bad side effects<br>8 The wait times were too long<br>9 I was no longer pregnant<br>10 I didn't believe my test result<br>11 I thought I was cured<br>12 Other |
| otherreason2 *(required)* | Specify | |
| V. Patient survey > Section 2: Post-test questions for those recruited before their HIV test. Questionnaire will continue directly from Section 1 to Section 2 for those recruited after their HIV test. > Questions for patients testing positive for HIV > Questions for patients with previous ART experience > Previous med group | | |
| disclosetostaff *(required)* | 3. Did you disclose the fact that you were previously on ART to any staff at the clinic today? | 1 Yes<br>0 No |
| where *(required)* | 4. Where in the process did you disclose your previous ART? | 1 Upon arrival<br>2 During the testing process<br>3 With the ART nurse<br>4 Other |
| specifywhere2 *(required)* | Specify | |
| transferletter *(required)* | 5. Did you bring a transfer letter to the clinic today? | 1 Yes<br>0 No |
| notransferletter *(required)* | 6. What was the reason that you did not bring a transfer letter? | 1 Did not know I would need one |

| Field | Question | Answer |
|---|---|---|
| | | 2 Did not have time to go to previous clinic<br>3 Did not have money to go to previous clinic<br>4 Was not treated nicely at previous clinic<br>5 Other |
| specifynoletter *(required)* | Specify | |
| V. Patient survey > Section 2: Post-test questions for those recruited before their HIV test. Questionnaire will continue directly from Section 1 to Section 2 for those recruited after their HIV test. > Questions for patients testing positive for HIV > Questions for patients with previous ART experience > Previous med group > Without ART subgroup | | |
| withoutart *(required)* | 7. How long have you been without ART medication?<br>*Days* | |
| withoutartmonths *(required)* | Now enter number of months | |
| withoutartyears *(required)* | Now enter number of years | |
| accessart *(required)* | 8. Were you able to access any ARV treatment in between your last clinic visit and today? | 1 Yes<br>0 No |
| howyouaccessed *(required)* | 9. How did you access ARV treatment? | 1 Borrowed from a spouse or family member<br>2 Accessed from a different clinic<br>3 Purchased from someone in the community<br>4 From a private pharmacy<br>5 Other |
| specifyaccess *(required)* | Specify | |
| calls *(required)* | 10. Did you receive any calls, home visits, or SMSes from your previous clinic after you did not return for a clinic visit or to collect your ART medication? | 0 No<br>1 Call<br>2 Home visit<br>3 SMSs<br>4 Other tracing method (specify) |
| specifycalls *(required)* | Specify | |
| influence *(required)* | 11. How did these influence your decision to come to the clinic to test today? | |
| V. Patient survey > Section 2: Post-test questions for those recruited before their HIV test. Questionnaire will continue directly from Section 1 to Section 2 for those recruited after their HIV test. > Questions for patients testing positive for HIV > Questions for patients with previous ART experience > Questions about ART initiation | | |
| readiness *(required)* | 12. How ready do you feel to initiate or re-initiate ART today? | 1 Definitely ready<br>2 Somewhat ready<br>3 Not ready at all |
| startart *(required)* | 13. Were you offered the chance to start or restart ART today? | 1 Yes<br>0 No |
| acceptoffer *(required)* | 14. Did you accept the offer to start ART today, meaning did the nurse or pharmacist give you ARVs to take home with you today? | 1 Yes<br>0 No |
| why *(required)* | Why or why not? | |
| expecttostart *(required)* | 15. When do expect to start? | 1 This week<br>2 This month<br>3 Later this year<br>4 Sometime in the future<br>5 Never<br>6 Don't know<br>7 Other (specify) |
| specifywhen *(required)* | Specify | |
| mainreason *(required)* | 16. What is your main reason for starting or restarting ART today? | 1 Pregnancy<br>2 Feeling ill and wanted to feel better<br>3 Diagnosed with TB<br>4 Health care provider recommended starting/restarting<br>5 Was worried about not being on treatment<br>6 Other (specify) |
| specifymainreason *(required)* | Specify | |
| servicesprovided *(required)* | 17. Could you please list the services that were provided to you as part of HIV testing today? | 1 Pre-test counselling<br>2 Post-test counselling |

| Field | Question | Answer |
|---|---|---|
| | | [illegible]<br>3 Confidential space<br>4 Condoms<br>5 Family planning<br>6 TB symptom screen<br>7 STI symptom screen<br>8 GBV screening<br>9 TB prevention medication (IPT)<br>10 TB treatment<br>11 ART initiation<br>12 Adherence counselling<br>13 CD4 testing<br>14 TB test<br>15 Other lab test (specify)<br>16 Other service (specify)<br>17 Referral to other services (specify)<br>18 Return date for next clinic visit |
| specifyservicesprovided *(required)* | Specify | |
| learnabouthiv *(required)* | 18. Did you learn as much about HIV and HIV prevention as you wanted to know? | 0 No<br>1 Yes |
| learnwhatelse *(required)* | Specify what else you would have liked to learn | |
| followupservices *(required)* | 19. Do you intend to follow up on anything else you might have been offered? (e.g. using condoms or family planning, referral to other services) | 1 Yes<br>0 No |
| specifywhynot1 *(required)* | Specify which service(s) and why not) | |
| specifywhy1 *(required)* | Specify which service(s) | |
| V. Patient survey > Section 2: Post-test questions for those recruited before their HIV test. Questionnaire will continue directly from Section 1 to Section 2 for those recruited after their HIV test. > Questions for patients testing positive for HIV > Questions for patients with previous ART experience > Questions about ART initiation > Services received subgroup | | |
| supportive *(required)* | 20. Do you consider the service that you received today to be welcoming and supportive? | 1 Very supportive<br>2 Supportive<br>3 Moderately supportive<br>4 Slightly supportive<br>5 Not supportive |
| providereason *(required)* | 21. Please provide reasons for your response to Q20 | |
| timespent2 *(required)* | 22. About how long did you spend at the clinic today, including for this survey?<br>*Interviewer note: we want to know how long it takes to get a test, start ART etc* | |
| timespentmin2 *(required)* | Surveyor: Now enter the minutes | |
| V. Patient survey > Section 2: Post-test questions for those recruited before their HIV test. Questionnaire will continue directly from Section 1 to Section 2 for those recruited after their HIV test. > Questions for patients testing negative for HIV | | |
| testingexp *(required)* | 1. What services did you receive as part of your testing experience today? | 1 Pre-test counselling<br>2 Post-test counselling<br>3 Confidential space<br>4 Condoms<br>5 Family planning<br>6 Offered PrEP<br>7 TB symptom screen<br>8 STI symptom screen<br>9 GBV screening<br>10 TB treatment<br>11 TB test<br>12 Other lab test (specify)<br>13 Other service (specify)<br>14 Referral to other services (specify)<br>15 Return date for next clinic HIV test |
| specifyexp *(required)* | Specify | |
| learn *(required)* | 2. Did you learn as much about HIV and HIV prevention as you wanted to know? | 0 No<br>1 Yes |
| whatelse *(required)* | Specify what else you would have liked to learn | |
| acceptprep *(required)* | 3. Did you accept PrEP if offered? | 0 No, did not accept<br>[illegible] |

| Field | Question | Answer |
|---|---|---|
| | | 1 No, was not offered<br>2 Yes |
| specifyprep *(required)* | Specify why not | |
| followuponservives *(required)* | 4. Do you intend to follow up on anything else you might have been offered? (e.g. using condoms or family planning, referral to other services) | 1 Yes<br>0 No |
| specifywhynot *(required)* | Specify which service(s) and why not) | |
| specifywhy *(required)* | Specify which service(s) | |
| timespent *(required)* | 5. About how long did you spend at the clinic today, including for this survey?<br>*Interviewer note: we want to know how long it takes to get a test, start ART etc* | |
| timespentmin *(required)* | Surveyor: Now enter the minutes | |
| closing | Please thank the participant for their time and ask if they have any additional questions about the study. | |
| notes *(required)* | Surveyor notes | |
| sid_2 *(required)* | Survey ID options | 1 Barcode<br>2 Enter manually |
| barcode_scan_2 *(required)* | Scan survey ID | |
| survey_id_repeat *(required)* | SURVEY ID | |
| not_eligible | This patient is NOT ELIGIBLE for the study. Thank the participant for their time but do not proceed with the survey | |
| surveyor_initials *(required)* | Surveyor initials | |
