## Supplementary file 8 for "The SENTINEL study of differentiated service delivery models for HIV treatment in Malawi, South Africa, and Zambia: research protocol for a prospective cohort study"

### SENTINEL Patient Survey Testing

#### RESEARCH INFORMATION FORM FOR PATIENT SURVEY (TESTING)

**Title of Project: Outcomes of Differentiated Models of Service Delivery for HIV Treatment at Sentinel Sites in South Africa (Sentinel-South Africa)**

**Principal Investigators: Sophie Pascoe and Amy Huber**

---

| | |
|---|---|
| Study Number: | M210241 |
| Study Title: | **Outcomes of Differentiated Models of Service Delivery for HIV Treatment at Sentinel Sites in South Africa (Sentinel-South Africa)** |
| Sponsor: | Bill & Melinda Gates Foundation |
| Investigators: | Dr. Sophie Pascoe, Principal Researcher (HE$^2$RO, South Africa) |
| | Dr. Amy Huber, Senior Researcher (HE$^2$RO, South Africa) |

Good day. My name is _______________, and I am a research assistant at the Health Economics and Epidemiology Research Office (HE$^2$RO), part of Wits University in Johannesburg. I would like to invite you to consider participating in a research study entitled "Outcomes of Differentiated Models of Service Delivery for HIV Treatment at Sentinel Sites in South Africa (Sentinel-South Africa).

**What is this research study about?**

HE$^2$RO and Boston University in the United States are conducting a research study about different ways that clinics are delivering HIV testing in South Africa and how these different ways of delivering testing affect both the clinics and their staff and the patients who are receiving testing.

From patients like you, we would like to find out more about your HIV testing experience, how this was done and your satisfaction with the services you are receiving. We are inviting adult patients in this clinic who are receiving HIV testing services offered by this clinic to participate in the study. If you choose to participate, you will be asked to answer a set of questions today, and we will also look at your clinic records.

This study is being conducted by Drs. Sophie Pascoe and Amy Huber from HE$^2$RO (Wits University) in South Africa and they are supported by other investigators from HE$^2$RO and from Boston University in the U.S. This study is being sponsored by the Bill & Melinda Gates Foundation in the United States.

I would like to provide you with some more information about the study, so that you can decide whether you would like to take part in it. Taking part in this study is voluntary. You can choose not to take part, and if you join, you may quit at any time. If you decide not to participate, you will receive the same care from this clinic that you would have received otherwise. This information sheet may have some words that you do not understand. Please ask the study staff to explain any words or information that you do not understand.

**What is the purpose of this research study?**

In many countries, including South Africa, the Department of Health and healthcare providers are trying to find easier ways to provide HIV testing services to the many thousands of patients who need it. This effort has led to the development of what are called “differentiated service delivery” (DSD) for HIV testing, which adjust the timing, location, and other aspects of HIV testing for different kinds of patients. The goal of DSD for HIV testing is to make testing more accessible to patients so that they can more easily test and link to treatment or prevention services and be more satisfied with the services they received.

In South Africa, the Department of Health offers multiple testing models including mobile testing, self-testing, and testing at the clinic. Some individual clinics and partner organizations have created other models as well (e.g. such as pop-up testing sites, testing campaigns, etc.). It is important to now try and find out how many patients are participating in each model, how well the models are performing and whether patients and healthcare providers are satisfied with them. This study is being conducted in collaboration with the Department of Health to try to find answers to these questions, so that different models of HIV testing and linkage to treatment and prevention services can be improved in the future.

**What happens in this research study?**

This study is taking place in 24 clinics in South Africa. You will be one of approximately 1200 patients to be asked to participate in this study. Patients in this study must be at least 18 years old and receiving HIV testing services offered by your clinic. If you participate in the study, it will take about 45 minutes of your time today.

If you agree to participate in this study, I will ask you a set of question about you, your household, your health, any previous HIV testing experience and your experience of receiving HIV testing services at this clinic today and your satisfaction with the services at this clinic. The questions will take about 30-45 minutes to complete and will be conducted in English, Zulu or Sesotho. You may have been asked to join the study before your HIV test. If you were, we will conduct informed consent and the first part of the questionnaire before your HIV test and we will then invite you to return after your visit is completed to do the second part of the questionnaire. If you were asked to join the study after your HIV test, we will complete the informed consent form and the full questionnaire at one time. You have a choice to complete the questionnaire on a later date if you would prefer. In order for us to complete the questionnaire on a later date, we will ask for your telephone number and when would be a convenient time to call you after one week to complete the questionnaire.

We will also link your answers to the questions today to your clinic medical records. The reason that we need to do this is so we can see whether patients with different characteristics and experiences and in different models of HIV testing link to HIV treatment or prevention services as recorded in their medical records. To make this link, we will ask you your name and information needed to find your individual medical record. After we link your clinic records to the question responses, all links between your answers and your clinic records will be made anonymous—they will not include your name or any other identifiers. We would like to ask your permission to link the answers you give today with your clinic medical records starting when you tested for HIV and continuing for the next twenty-four (24) months from when the survey was conducted, including if you transfer to or link to services at another clinic.

**What other choices do I have?**

Your alternative is not to participate in this study.

**Are there any costs or payments to me?**

You will not incur any costs for your participation in the study. HIV testing and ART are free at this clinic. You will be offered light refreshments during the study. In addition, after completing the study interview you will receive a voucher valued at R150, to thank you for your time and effort in participating in the study.

### SENTINEL Patient Survey Testing

#### RESEARCH CONSENT FORM FOR PATIENT SURVEY (TESTING)

Title of Project: Outcomes of Differentiated Models of Service Delivery for HIV Treatment at Sentinel Sites in South Africa (Sentinel-South Africa)

Principal Investigators: Sophie Pascoe and Amy Huber

---

INFORMED CONSENT SIGNATURE PAGE

Signing this consent form indicates that you have read the consent form information sheet (or have had it read to you), that your questions have been answered to your satisfaction, and that you voluntarily agree to participate in this research study. You will receive a copy of this consent form to keep.

| ___________________ | ___________________________ | _____________ | ______ |
|---|---|---|---|
| Participant (Signature) | (Printed Name and Surname) | Date | Time |

| ___________________ | ___________________________ | _____________ | ______ |
|---|---|---|---|
| Surveyor (Signature) | (Printed Name and Surname) | Date | Time |

| ___________________ | ___________________________ | _____________ | ______ |
|---|---|---|---|
| Witness* (Signature) | (Printed Name and Surname) | Date | Time |

*Witness signature required if patient provides mark or thumbprint rather than signature

| **Participant Survey ID Number** |
|---|
